# Relapse and delay-induced dynamics in human-to-human Nipah virus transmission: a mathematical modeling study

**DOI:** 10.64898/2026.03.02.26347485

**Authors:** Sarita Bugalia, Hao Wang, Swati Tyagi, Jai Prakash Tripathi, Liliana C.M. Salvador

## Abstract

Nipah virus (NiV) exhibits complex transmission dynamics shaped by infection-to-transmission delays, relapse, and delayed-onset neurological complications following recovery. The epidemiological consequences of these temporal processes, particularly how they jointly shape NiV dynamics, remain poorly characterized. We develop a delay differential equation model for human-to-human NiV transmission incorporating an effective population-level infection-to-transmission delay, immediate relapse, and delayed encephalitis relapse. We derive the transmission threshold ℛ_*h*_ and establish a unique feasible endemic equilibrium when ℛ_*h*_ > 1. The disease-free equilibrium is unstable whenever ℛ_*h*_ > 1, with a forward transcritical bifurcation at ℛ_*h*_ = 1. Delays can destabilize the endemic equilibrium, inducing sustained oscillations through Hopf bifurcation under hypothetical regimes. Calibration to Bangladesh incidence data (2001-2024) yielded ℛ_*h*_ ≈ 0.83 (95% bootstrap CI 0.74-1.03), with profile-likelihood regions spanning 0.67-0.95 and 96% of replicates subcritical. Relapse rates were not practically identifiable from annual incidence data; as the gradient of ℛ_*h*_ is nearly orthogonal to the weak parameter direction, subcriticality is unaffected and relapse contributions are interpreted as effective replenishment. The model generated recurrent incidence through relapse and delay feedback; projected resurgence applies only within the proposed framework. Sensitivity analyses show an oblique ℛ_*h*_ = 1 contour within the model: a 30% increase in immediate relapse lowers the required transmission increase from 21% to 10%. Immediate relapse most strongly shapes post-peak burden, while the infection-to-transmission delay controls epidemic timing and magnitude. Delays and relapse-mediated feedback can generate transient waves even when ℛ_*h*_ < 1, underscoring the importance of monitoring recovered individuals.

## 1 Introduction

Nipah virus (NiV) disease is a deadly emerging contagious disease that poses a growing public-health concern in Southeast Asia. It is caused by the zoonotic single-stranded RNA NiV, which is a member of the Henipavirus genus in the Paramyxoviridae family with subfamily Orthoparamyxovirinae [1]. Since its first recognized outbreak in Singapore and Malaysia in 1998-1999, NiV has caused recurrent outbreaks with substantial case fatality and significant socio-economic disruption [2–4]. Several hundred cases have been reported globally, and the reported case fatality has ranged from 40% to 75% in outbreaks since 2001. This underscores the necessity for a better understanding of the transmission dynamics and control of the disease [5].

Fruit bats are the primary natural reservoir of NiV, disseminating the virus in saliva, urine, semen, and excreta, and serving as the primary zoonotic source of human infection [6]. Evidence of NiV infection in fruit bats has been reported throughout Southeast Asia [5, 7, 8], with serological evidence also documented outside the region [9–12] as well. Outbreaks of NiV disease in human populations show a strong seasonal pattern during spring and winter, which is likely connected with the date palm sap harvesting season [5] and the breeding season of Pteropus (flying foxes) bats. In addition to zoonotic spillover, human-to-human transmission occurs through close contact with infected individuals and exposure to respiratory secretions [5, 13–16]. Human-to-human transmission was particularly evident during outbreaks in Bangladesh and India, which accounted for approximately 51% and 75% of cases, respectively [17]. Reported incubation periods of NiV in many humans are commonly 4-14 days [2, 5], although longer delays have been documented [18].

Clinically, NiV infection has a broad clinical spectrum ranging from asymptomatic or non-encephalitic illness to severe pneumonia and encephalitis [19]. Encephalitis, characterized by necrosis and vasculitis in the central nervous system (CNS), may present as either acute or late-onset. The latter can be difficult to diagnose since NiV exposure may have occurred several months earlier. Additionally, late-onset encephalitis has been reported several months after initial recovery [18, 20]. A study by Chattu et al. [17] confirmed that over 20 cases of relapsing NiV encephalitis have been observed, including one occurring 11 years after an asymptomatic infection. As of 2025, no licensed therapies or vaccines are available for NiV disease [5, 22]. Current treatment is limited to supportive care and symptom management; however, a vaccine is currently under development [23], and several antiviral drugs are under investigation, such as ribavirin, which has shown potential in reducing mortality in acute Nipah encephalitis [24]. To reduce transmission, avoidance of close contact with infected individuals, regular hand washing, and the use of protective equipment when caring for sick individuals are recommended [25].

In recent years, several mathematical studies have examined NiV transmission dynamics using compartmental models [26–37]. Biswas et al. [26] developed an SIR model with optimal control and showed that educational interventions discouraging consumption of raw date palm sap can reduce transmission. Mondal et al. [27] developed a model that includes quarantine and monitoring restrictions. They concluded that quarantine and improved hygiene can effectively control the spread of disease. Khan et al. [28] developed a model encompassing human-to-human and food-borne transmission, identifying improved treatment, diminished contact rates, and heightened vaccine efficacy as key drivers in disease control. Xie et al. [29] showed that time-varying interventions can work better than constant controls in lowering incidence, while Yu et al. [30] examined the global dynamics of NiV infection under vaccination and treatment. Tyagi et al. [31] introduced an asymptomatic class and examined equilibrium stability, determining that early-stage intervention can significantly diminish transmission. Barua and Dénes [32] developed a multi-host compartmental model incorporating transmission among bats, pigs, and humans and analyzed the global dynamics of the disease.

More recently, Dutta et al. investigated NiV transmission through food-borne, human-to-human, and deceased-body transmission routes and examined equilibrium behavior together with sensitivity and uncertainty analysis [33]. Another recent study developed a multi-host framework incorporating transmission among bats, pigs, and humans and investigated intervention strategies through control optimization [34]. Rathinasamy and Gnanaprakasam [35] considered NiV transmission using Atangana-Baleanu Caputo fractional derivatives together with physics-informed neural networks. Boulaaras et al. [36] developed a comprehensive multi-host framework and examined threshold behavior, stability properties, and numerical dynamics, while Haque et al. [37] investigated bifurcation behavior, sensitivity, and optimal control in NiV transmission. These studies provide important insights into multi-route transmission, multi-host interactions, intervention strategies, stability, bifurcation behavior, uncertainty, and alternative mathematical formulations of NiV dynamics.

However, despite clinical documentation of relapse and delayed-onset encephalitis, their mechanistic interaction with infection-to-transmission delays remains mathematically uncharacterized. NiV transmission involves multiple temporal processes, including the interval between exposure, disease progression, clinical recognition, and subsequent contribution to onward transmission. We represent these aggregated temporal processes through an effective infection-to-transmission delay. In the model, we consider this delay as the population-level lag between the generation of new infections and their delayed contribution to the infectious classes. When estimated from annual incidence data, this time delay is interpreted as an effective dynamical lag of the aggregated transmission process. Relapse and delayed-onset encephalitis can reintegrate recovered persons into infectious compartments, perpetuating transmission or producing recurring epidemic waves, while the effective infection-to-transmission delay may influence the timing and magnitude of outbreaks. To our knowledge, existing NiV modeling studies have not jointly incorporated immediate relapse, delayed-onset encephalitis, and an effective infection-to-transmission delay within a unified human-to-human transmission framework. Addressing this gap is important for understanding how these temporal and post-recovery mechanisms may shape NiV transmission dynamics and for informing future disease-control strategies.

We propose a delay differential equation (DDE) model for human-to-human NiV transmission, with the aim of capturing key temporal characteristics indicated by clinical and surveillance evidence. Nipah virus outbreaks are initiated through complex ecological and epidemiological processes, including zoonotic and foodborne spillover. In the present study, we do not explicitly model these spillover transmission routes because data on the timing, magnitude, and frequency of spillover introductions are limited. Instead, we focus on the human-to-human transmission phase after infection has been introduced into a human population. This restricted formulation allows us to isolate the effects of an effective population-level infection-to-transmission delay, relapse, and delayed post-recovery complications on human transmission dynamics. More specifically, the model includes an effective infection-to-transmission delay describing progression from exposure to infectiousness, a relapse pathway that returns recovered individuals to a less severe infectious stage, and a post-recovery delay reflecting development of delayed-onset encephalitis. We characterize the equilibria and their stability, examine Hopf bifurcations and delay-induced oscillatory dynamics, and assess whether relapse and post-recovery delays can produce recurring NiV transmission patterns in humans. Model parameters, including the transmission rate, relapse rates, time delays, and selected initial conditions, are estimated from annual NiV incidence data from Bangladesh. Finally, numerical simulations, bootstrap uncertainty analysis, and scenario-based sensitivity analyses are used to quantify how relapse and delay mechanisms shape outbreak timing, peak magnitude, and post-peak dynamics across epidemiologically plausible regimes.

Two findings emerge that extend beyond the present model. First, subcritical human-to-human transmission combined with relapse-mediated replenishment can sustain recurrent multi-year incidence waves, a behaviour that models without post-recovery feedback cannot produce and that is distinct from recurrence driven by repeated introductions. Second, annual incidence data do not constrain the individual relapse rates, yet the transmission threshold remains well determined because its gradient is nearly orthogonal to the weakly identified parameter direction. This separation between identifiable summary quantities and unidentifiable constituent rates is likely to arise for any relapsing infection observed at coarse temporal resolution, and it indicates which quantities can be reported from aggregated surveillance data and which require post-recovery follow-up.

The rest of the paper is organized as follows. Section 2 presents the model formulation and establishes positivity and boundedness of solutions. Section 3 quantifies the contribution of relapse to infection dynamics. Section 4 develops the dynamical analysis, including computation of the human-to-human transmission threshold (ℛ_*h*_), the stability analysis of equilibria, and Hopf bifurcation. Section 5 describes parameter estimation and model fitting. Section 6 presents numerical simulations, sensitivity analyses, and illustrative bifurcation results. Section 7 discusses implications, limitations, and future directions.

## 2 Model structure

In this study, we extend the framework of Tyagi et al. [31] to develop a compartmental model of humanto-human Nipah virus (NiV) transmission through direct contact. The model incorporates an effective infection-to-transmission delay, a short-timescale relapse pathway, and a delayed relapse pathway associated with delayed-onset encephalitis. The total human population at time *t*, denoted by *N* (*t*), is divided into seven epidemiological compartments: susceptible individuals *S*(*t*), exposed but noninfectious individuals *E*(*t*), early-stage infectious individuals with milder symptoms *I*_1_(*t*), later-stage infectious individuals with severe disease *I*_2_(*t*), hospitalized individuals with severe disease *H*(*t*), individuals receiving supportive care *T* (*t*), and recovered individuals *R*(*t*). Thus,

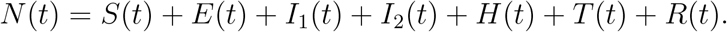

The proposed model with multiple discrete time delays is described by the following system of equations:

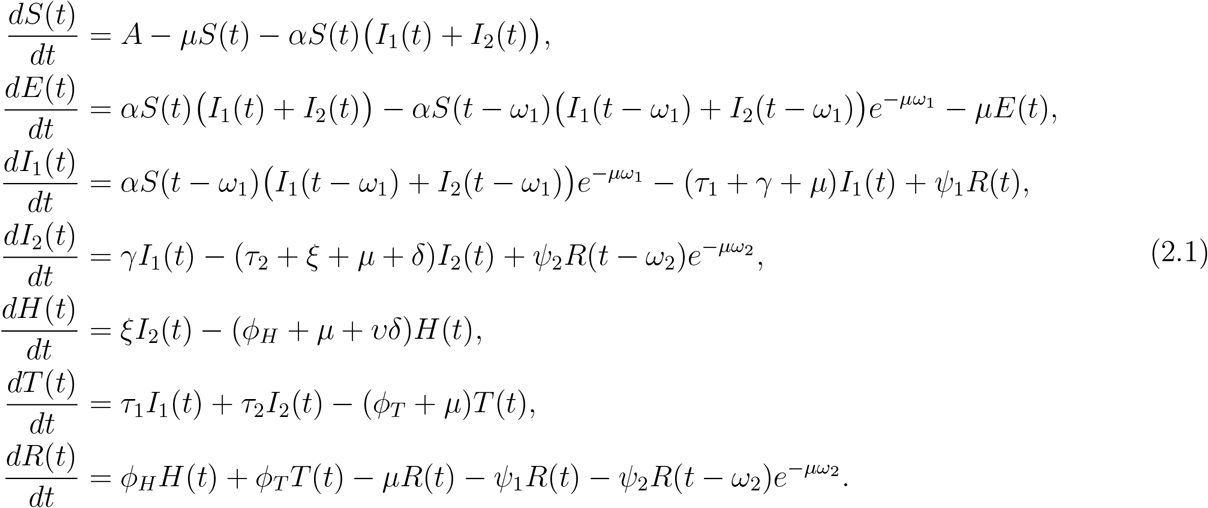

The schematic diagram of model (2.1) is shown in Figure 1. In model (2.1), individuals in *I*_1_(*t*) and *I*_2_(*t*) are assumed to transmit infection at the same effective coefficient *α*. Stage-specific transmissibility estimates are not available for NiV, and introducing separate coefficients would add a parameter that annual incidence data cannot identify. The two compartments nonetheless remain epidemiologically distinct, differing in progression, treatment, hospitalization, disease-induced mortality, and relapse pathway.

**Figure 1:**
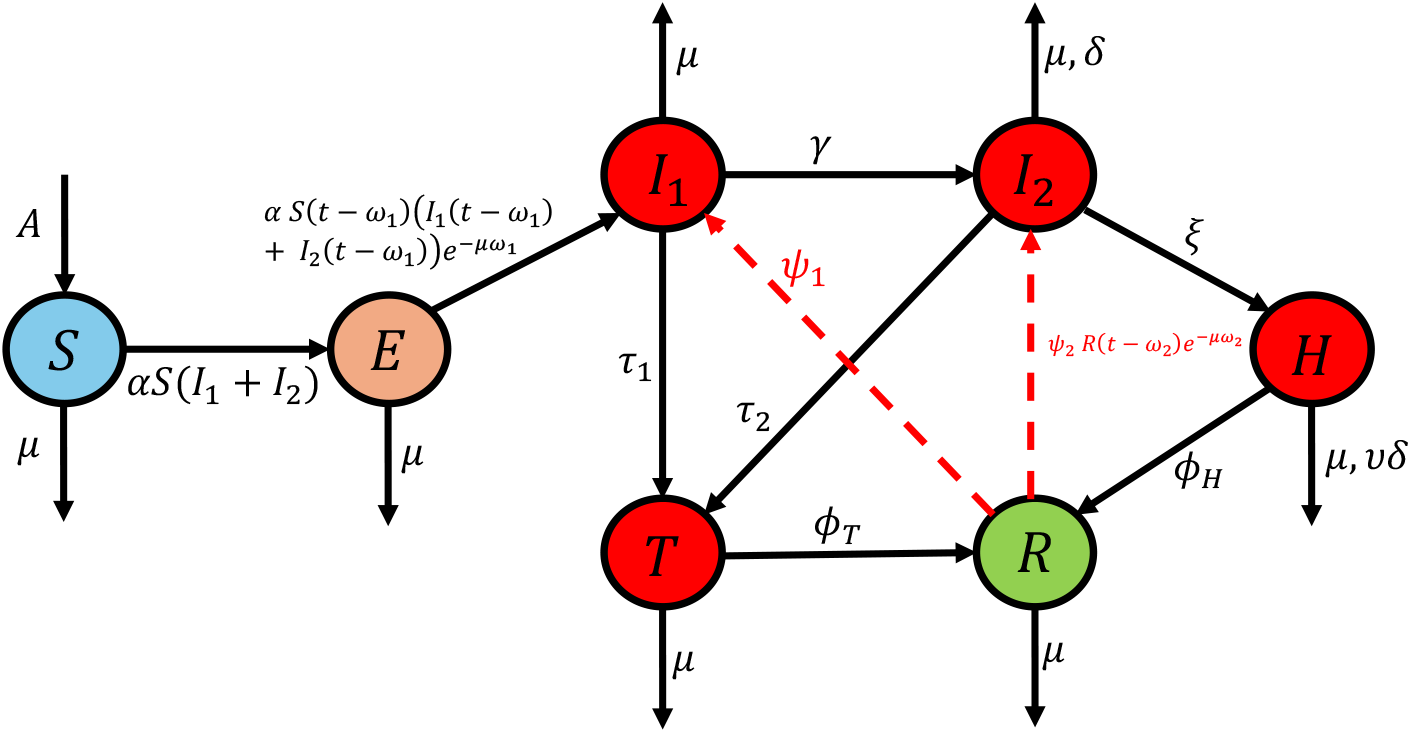
Schematic diagram illustrating the flow between the different compartments in the model (2.1).

Incidence is modeled in the mass-action form *αS*(*t*) (*I*_1_(*t*) + *I*_2_(*t*)). Because the total population remains approximately constant over the study period, mass-action and frequency-dependent incidence differ essentially by a rescaling of the transmission coefficient and yield equivalent fitted dynamics; we therefore interpret *α* as an effective, population-specific transmission coefficient rather than as a contact rate with absolute meaning. A frequency-dependent formulation, in which the per-capita contact rate remains bounded as population size increases, would be the more appropriate choice in settings where population size varies substantially.

The effective infection-to-transmission delay *ω*_1_ denotes the time between the acquisition of infection and progression to the early infectious class *I*_1_. Accordingly, the term 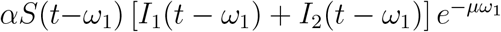 represents the rate at which individuals infected at time *t* − *ω*_1_ enter *I*_1_ at time *t*, where 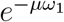 is the probability of surviving natural mortality over the delay period and *µ* is the natural mortality rate. The less symptomatic infectious individuals either progress to the severe disease stage or move to the supportive care compartment, depending on their disease status. Disease-induced mortality occurs in the severe infectious compartment *I*_2_ and the hospitalized compartment *H*. We assume that hospitalized individuals are no more likely to die than individuals in *I*_2_, and scale their disease-induced mortality rate by the parameter 0 *< v* ≤ 1. Thus, *v* = 1 means that hospitalized individuals experience the same disease-induced mortality rate as those in *I*_2_, while values closer to zero represent lower disease-induced mortality among hospitalized individuals.

We consider two relapse pathways to reflect the clinical heterogeneity of post-recovery Nipah virus outcomes. The term *ψ*_1_*R*(*t*) represents a short-timescale relapse or recrudescence process in which recovered individuals return to the less severe infectious class *I*_1_. This pathway is included phenomenologically to capture mild recurrence or rapid post-acute recrudescence after apparent recovery. The second pathway, 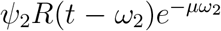, represents delayed relapse into the more severe symptomatic class *I*_2_. In the present formulation, this term is interpreted as a phenomenological delayed-state feedback, in which the rate of delayed severe or neurological relapse at time *t* depends on the size of the recovered population at the earlier time *t* − *ω*_2_. Delayed-state formulations of this general type have been used in epidemic delay differential equation models to represent lagged epidemiological effects [38–40]. Accordingly, *ω*_2_ represents an effective population-level post-recovery lag rather than an exact individual-level residence time in the recovered compartment. The factor 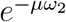 attenuates the delayed feedback by natural mortality over the lag, by analogy with the survival weighting that arises in cohort-based delay formulations; in the present phenomenological setting it scales the magnitude of the delayed contribution rather than tracking the survival of an identified cohort; a cohort-structured alternative is discussed in Section 7. This pathway is motivated by documented relapsed and late-onset Nipah encephalitis after apparent recovery, including relapsed encephalitis among survivors of acute Nipah encephalitis and late-onset encephalitis among individuals with initially non-encephalitic or asymptomatic infection [20]. The proposed model extends [31] by incorporating short-timescale and delayed relapse pathways through the parameters *ψ*_1_, *ψ*_2_, and *ω*_2_, and differs in the representation of progression from the exposed class to the infectious class. In [31], exposed individuals progress from the compartment *E* to the early infectious compartment *I*_1_ at a constant rate *λ*, corresponding to an exponentially distributed waiting time in the exposed class. In the present model, this mechanism is replaced by a fixed effective infection-to-transmission delay *ω*_1_. Thus, individuals infected at time *t*− *ω*_1_ who survive the delay period enter the infectious compartment *I*_1_ at time *t*. Unless otherwise stated, all model parameters are assumed to be positive, while the delay parameters are nonnegative. The biological interpretations of the model parameters are summarized in Table 1.

**Table 1:** Biological interpretation of the parameters in model (2.1).

| Parameters | Biological interpretation |
| --- | --- |
| $A$ | Recruitment rate of individuals joining the susceptible compartment |
| $\mu$ | Natural death rate |
| $\alpha$ | Effective disease transmission rate |
| $\gamma$ | Progression rate from less symptomatic stage to severe stage |
| $\tau_1$ | Treatment rate of less symptomatic individuals |
| $\tau_2$ | Treatment rate of severe diseased individuals |
| $\xi$ | Hospitalization rate of severe diseased individuals |
| $\delta$ | Disease-induced death rate |
| $\psi_1$ | Rate of short-timescale relapse or recrudescence from $R$ to $I_1$ |
| $\psi_2$ | Rate associated with delayed severe or neurological relapse from $R$ to $I_2$ |
| $\phi_H$ | Recovery rate of hospitalized individuals |
| $v$ | Coefficient accounting for reduced mortality in hospitalized ($H$) compared to infectious ($I_2$) individuals |
| $\phi_T$ | Recovery rate of treated individuals |
| $\omega_1$ | Effective population-level infection-to-transmission delay |
| $\omega_2$ | Effective population-level post-recovery lag associated with delayed relapse or recurrent neurological disease |

To determine the solutions of model (2.1), we consider the following associated initial conditions

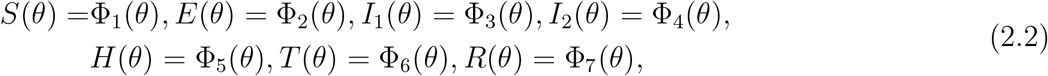

where *ω* = max {*ω*_1_, *ω*_2_}, *θ* ∈[ −*ω*, 0], (ϕ_1_(*θ*), ϕ_2_(*θ*), ϕ_3_(*θ*), ϕ_4_(*θ*), ϕ_5_(*θ*), ϕ_6_(*θ*), ϕ_7_(*θ*)) ∈ *C*([ −*ω*, 0], ℝ^7^), and *C*([ −*ω*, 0], ℝ^7^) is the Banach space of continuous functions from [−*ω*, 0] to ℝ^7^. Since each population has non-negative values, we consider non-negative initial histories, i.e.

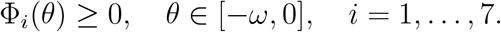

In addition, to ensure positivity of the solutions for the compartment *E* (and to preserve its biological cohort interpretation), the initial condition for *E* is defined in relation to the other variables as the following compatibility condition:

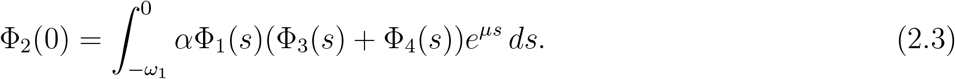

This compatibility condition is not required for local existence and uniqueness of solutions of the delay system, but it allows the exposed class to be represented as

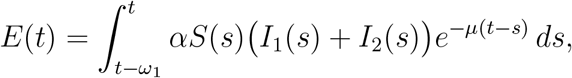

which implies *E*(*t*)≥0 whenever the remaining compartments are non-negative. Condition (2.3) is therefore a mathematical compatibility condition used to preserve the cohort interpretation and nonnegativity of the exposed compartment, rather than an additional biological assumption.

Because the delayed loss term 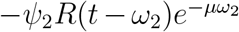, depends on the past recovered population, the standard quasipositivity argument does not apply directly to the *R*-equation. We therefore restrict attention to biologically admissible trajectories satisfying

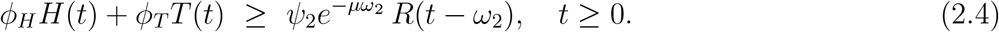

Under this assumption, the recovered equation satisfies

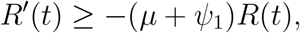

and hence *R*(*t*) ≥ 0 for *t* ≥ 0 whenever *R*(0) ≥ 0. For the calibrated parameter set used in the numerical analysis, condition (2.4) is verified numerically along the corresponding solution trajectory (see Section 5).

The right-hand side of system (2.1) defines a locally Lipschitz functional on the phase space *C*([−*ω*, 0], ℝ^7^), where *ω* = max {*ω*_1_, *ω*_2_} . Hence, for every continuous initial history satisfying the compatibility condition, the standard existence and uniqueness theorem for retarded functional differential equations guarantees a unique local solution [43]. Before examining the asymptotic properties of system (2.1), we establish positivity and boundedness of the solutions under the non-negativity and admissibility assumptions stated below. These a priori bounds prevent finite-time escape from bounded subsets of the phase space and, by the standard continuation theorem for retarded functional differential equations, extend the local solution uniquely to all *t* ≥ 0. The corresponding biologically feasible region is stated in Theorem 2.1, and the detailed proof is presented in Appendix A.

### Theorem 2.1.

*Assume that the initial history is continuous and non-negative, satisfies the compatibility condition* (2.3), *and that condition* (2.4) *holds along the solution trajectory. Then the solution of system* (2.1) *exists globally and remains non-negative. Moreover, the region*

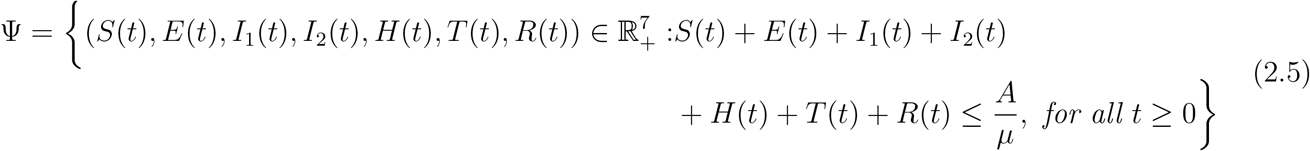

*is positively invariant. Furthermore, all non-negative solutions of system* (2.1) *are bounded and satisfy*

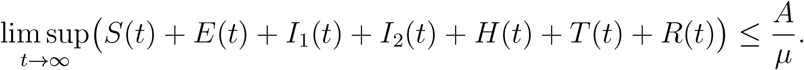

## 3 Contribution of relapse to infection dynamics

To assess when relapse becomes an epidemiologically important mechanism shaping epidemic dynamics, we introduce two model-derived diagnostic measures rather than established epidemiological indices. The first compares the size of the recovered population with the actively infectious population, reflecting the availability of individuals who may re-enter infectious states through relapse. The second quantifies the relative contribution of relapse-mediated returns to the infectious compartments compared with entries arising from human-to-human transmission. These measures are intended to summarize, along a model trajectory, the relative importance of relapse processes. The biological motivation for considering relapse after apparent recovery is supported by documented post-recovery recurrence in Nipah virus infection [20, 21].

We define the pool ratio at time *t* as the ratio of recovered individuals to the total infectious population,

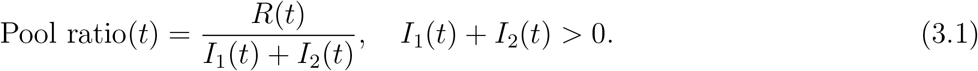

Large values of the pool ratio indicate that the recovered population substantially exceeds the number of currently infectious individuals. Within the present model, larger values of this ratio indicate a larger pool of recovered individuals available for relapse-mediated return to infectious states. The epidemiological importance of this mechanism therefore depends not only on the size of the recovered population but also on the relapse parameters and the competing transition rates.

In the model, incident infection arising from human-to-human transmission first enters the exposed class and subsequently enters the first infectious class after the effective population-level infection-to-transmission delay *ω*_1_. The corresponding rate of appearance of transmission-generated infectious individuals is given by

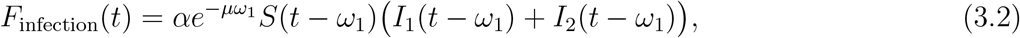

where the exponential term accounts for survival over the effective population-level infection-to-transmission period under natural mortality.

Relapse contributes to epidemic dynamics through two biologically distinct pathways. First, recovered individuals may return directly to the initial infectious class,

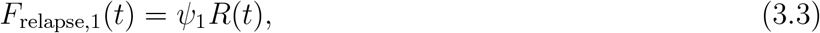

and second, relapse may occur after a delay *ω*_2_, contributing to the second infectious class,

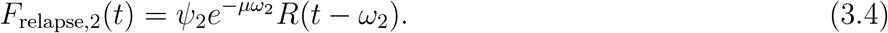

We then define the relapse contribution fraction as the proportion of total entries into infectious compartments attributable to relapse-related processes,

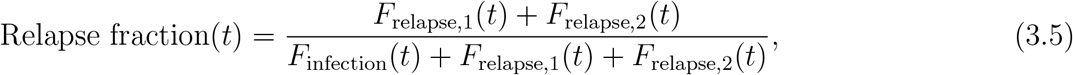

whenever the denominator is positive.

Values of the relapse fraction close to zero indicate that entries into infectious compartments are primarily shaped by human-to-human transmission, whereas larger values indicate that relapse-related returns to infectious states contribute substantially to the observed infectious burden. Although no sharp threshold separates primary-transmission-dominated dynamics from relapse-driven dynamics, the relapse contribution fraction provides insight into when relapse becomes epidemiologically relevant. In particular, relapse may play a significant role once the recovered population substantially exceeds the infectious population, especially during the declining or post-peak phase of the outbreak. This behavior is further illustrated via numerical simulations.

## 4 Dynamical analysis

To determine the long-term dynamics of the disease, we compute the disease-free and endemic equilibria of system (2.1), together with their feasibility conditions, representing states in which the infection either vanishes or persists. To characterize disease extinction or persistence, we first derive the human-to-human transmission threshold ℛ_*h*_. Next, we analyze the local stability of each equilibrium by linearizing the system about the corresponding equilibrium and examining the roots of the associated characteristic equation. This analysis is necessary because the inclusion of time delays renders the characteristic equation transcendental, with infinitely many characteristic roots that determine local stability. We identify parameter regimes in which all characteristic roots have negative real parts, implying local asymptotic stability, and examine conditions under which a pair of purely imaginary roots arises. In particular, for the endemic equilibrium, the time-delay parameters *ω*_1_ and *ω*_2_ are treated as bifurcation parameters, allowing us to determine critical values at which the qualitative dynamics of the system change.

### 4.1 Disease-free equilibrium and human-to-human transmission threshold

The disease-free equilibrium (DFE) is characterized by the absence of disease-related individuals. Thus, the exposed, infectious, hospitalized, treatment, and recovered compartments satisfy *E* = *I*_1_ = *I*_2_ = *T* = *H* = *R* = 0. Since the recovered class receives inflow only from the hospitalized and treatment classes, it also vanishes at the DFE. Substituting these conditions into the epidemiological subsystem of model (2.1), the disease-free equilibrium, denoted by *D*_0_, is

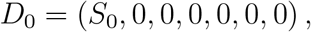

with 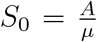 . We further compute the human-to-human transmission threshold, denoted by ℛ_*h*_, in the absence of additional zoonotic introductions. It measures the ability of human-to-human transmission to sustain disease spread after an initial introduction into the human population. Because model (2.1) contains delayed relapse feedback, we derive the transmission threshold directly from the cumulative infectious residence time generated by a newly infectious individual. New human-to-human infections are generated by infectious individuals in the classes *I*_1_ and *I*_2_. Thus, the transmission threshold is obtained by multiplying the transmission rate at the DFE by the total expected time spent in the infectious classes *I*_1_ and *I*_2_, accounting for progression, treatment, hospitalization, recovery, mortality, and relapse pathways.

For notational convenience, we define

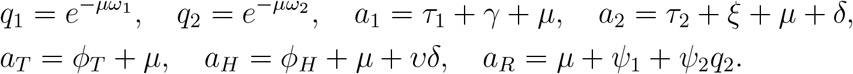

We consider the disease-transition subsystem obtained by suppressing the generation of new infections. We introduce one unit into the early infectious class *I*_1_ at *t* = 0 and assume zero disease history before the introduction, namely,

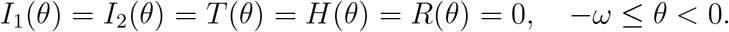

We further assume that condition (2.4) holds along the corresponding transition trajectory, so that all disease-related compartments remain non-negative. Let

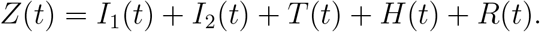

When the production of new infections is suppressed, summing the transition equations gives

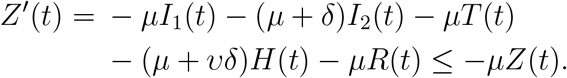

Therefore,

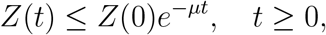

which ensures that the residence-time integrals are finite.

We construct the human-to-human transmission threshold directly from the cumulative infectious residence time generated by one newly infectious individual. This construction is used independently of the standard next-generation threshold theorem because the present delayed system is non-cooperative. The resulting matrix representation *FV* ^−1^ is analogous to the standard next-generation construction used for compartmental ODE models [44]; however, in the present DDE framework, the threshold property of ℛ_*h*_ is established independently through direct analysis of the characteristic equation.

Let *X*_1_ and *X*_2_ denote the total expected residence times in the infectious classes *I*_1_ and *I*_2_, respectively, following one unit entry into the infectious class *I*_1_. These residence times include possible return to infectiousness through relapse. Then *X*_1_ and *X*_2_ satisfy

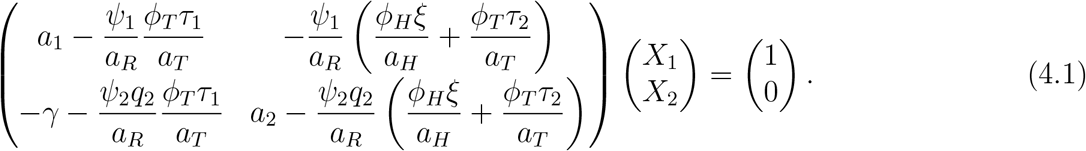

Therefore, the human-to-human transmission threshold is given by

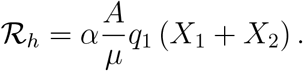

Solving the linear system (4.1) gives

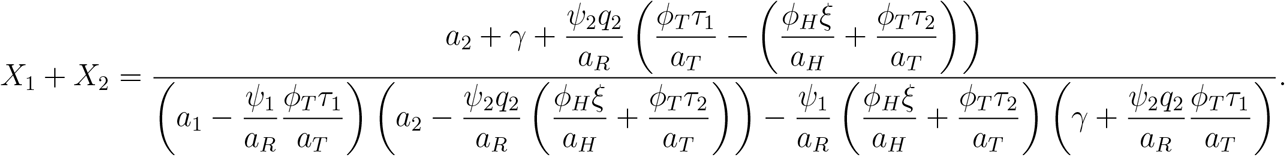

Equivalently, after clearing the common denominator *a*_*R*_, we obtain

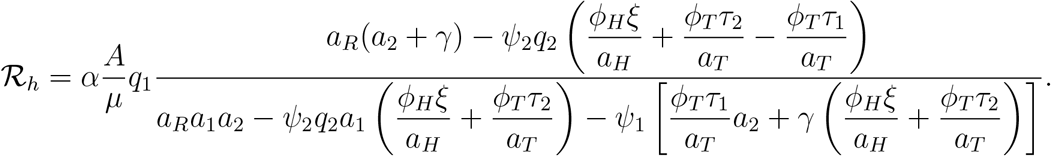

Substituting the definitions of *q*_1_, *q*_2_, *a*_1_, *a*_2_, *a*_*T*_, *a*_*H*_, and *a*_*R*_, the above expression is equivalent to

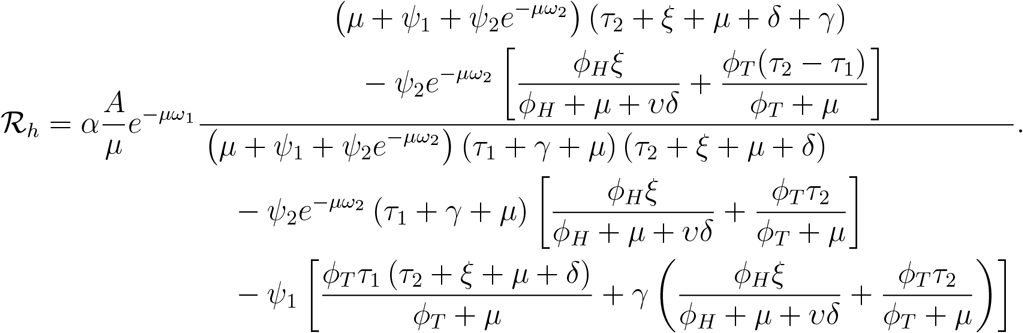

Under the biologically admissible parameter regime, the numerator and denominator of ℛ_*h*_ are positive. In particular, the denominator is bounded below by *µa*_1_*a*_2_ *>* 0, ensuring that the expected residence-time expression is well defined and finite. Consequently, the residence times *X*_1_ and *X*_2_ are non-negative, and for *α >* 0, the threshold ℛ_*h*_ is strictly positive. In ℛ_*h*_, the factor *αA/µ* represents the per-infective transmission rate in a fully susceptible population, where *A/µ* is the susceptible population size at the disease-free equilibrium. The factor 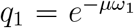 represents survival through the effective population-level infection-to-transmission delay. The quantity *X*_1_ + *X*_2_ represents the total expected time spent in the infectious classes *I*_1_ and *I*_2_, including possible return to infectiousness through relapse. Thus, ℛ_*h*_ represents the expected number of human infections generated by a typical infectious individual introduced into a wholly susceptible population under the model assumptions.

This residence-time interpretation of ℛ_*h*_ requires condition (2.4) to hold along the disease-transition trajectory, so that the residence-time integrals are finite and non-negative.

For the special case without relapse feedback, i.e., *ψ*_1_ = *ψ*_2_ = 0, the expression reduces to

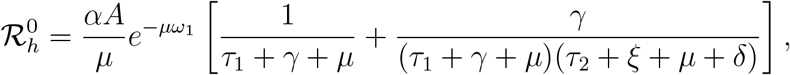

which corresponds to the expected infectious contribution from the *I*_1_ and *I*_2_ classes only.

### 4.2 Stability of disease-free equilibrium

Although the human-to-human transmission threshold ℛ_*h*_ characterizes the expected infectious contribution of a typical infected individual, it does not by itself provide a complete local stability criterion for the disease-free equilibrium in the present delay setting. The linearization contains the characteristic delay factors 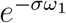 and 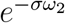, so the corresponding characteristic equation is transcendental and possesses infinitely many characteristic roots. For this reason, we determine the local stability of *D*_0_ by analyzing the characteristic roots of the full linearized delay system. The following theorem gives a spectral condition for local asymptotic stability of *D*_0_ for fixed *ω*_1_, *ω*_2_ ≥ 0.

#### Theorem 4.1.

*Let ω* = max{*ω*_1_, *ω*_2_}, *and assume µ >* 0. *Let*

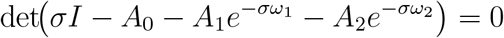

*denote the characteristic equation associated with the infection-feedback block of the linearization of system* (2.1) *at D*_0_. *If every root of this characteristic equation satisfies* ℜ(*σ*) *<* 0, *then D*_0_ *is locally asymptotically stable*.

*Proof*. Set

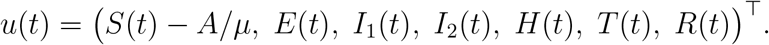

In a neighborhood of *D*_0_, system (2.1) defines a retarded functional differential equation on

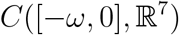

of the form

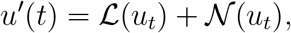

where ℒ is the Fréchet derivative at *D*_0_, and *N*(*φ*) = *o*(∥*φ*∥) as ∥*φ*∥ → 0.

To determine the local stability of *D*_0_, it suffices to consider the infection-feedback block

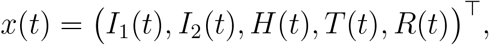

which satisfies the linear delay system

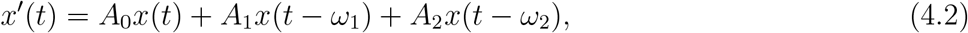

With

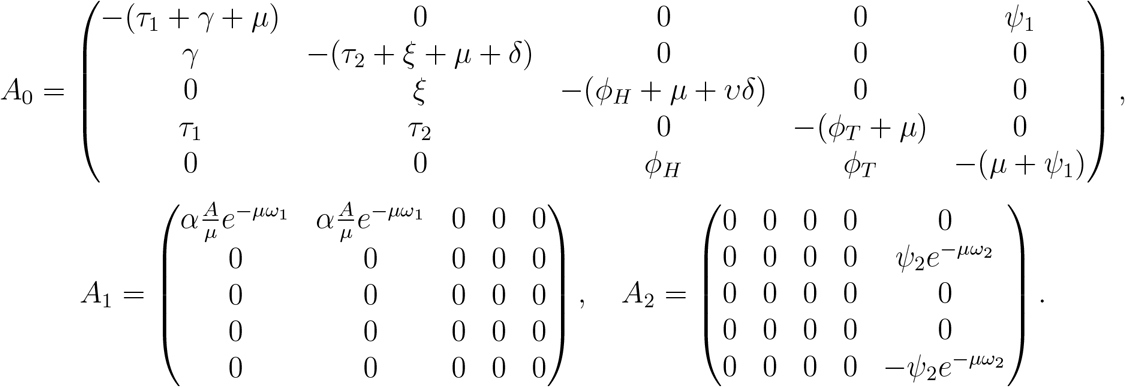

Seeking solutions of the form *x*(*t*) = *ve*^*σt*^ (*v*≠0) yields the characteristic matrix

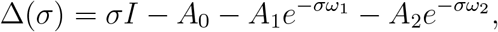

and hence the characteristic equations

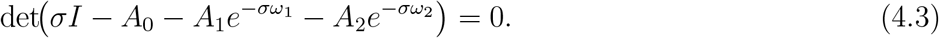

To relate the infection-feedback block to the full linearization, we write the linearized variables as

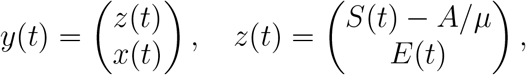

Where

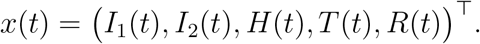

The full linearized system has block triangular form

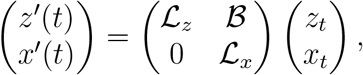

where ℒ_*x*_ is the infection-feedback operator associated with (4.2). The homogeneous *z*-subsystem is

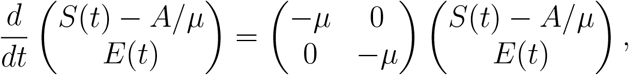

while the dependence on *I*_1_ and *I*_2_ enters only through the off-diagonal forcing operator . Consequently, the characteristic operator of the full linearization is block triangular, and its characteristic determinant factors as

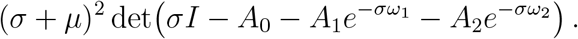

Hence the full spectrum consists of the stable root −*µ*, with multiplicity two, together with the characteristic roots of the infection-feedback block. Therefore, if every root of

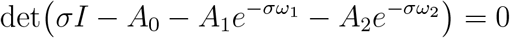

has negative real part, then every characteristic root of the full linearization lies in the open left halfplane. By the standard principle of linearized stability for retarded functional differential equations, if the spectrum of the linearized retarded system lies strictly in the open left half-plane, then the corresponding equilibrium of the nonlinear system is locally asymptotically stable [43].□

Theorem 4.1 provides the spectral criterion for local stability of the disease-free equilibrium. We next relate the human-to-human transmission threshold ℛ_*h*_ to the characteristic equation at *σ* = 0. The following proposition shows, without requiring cooperativity of the full delayed linearization, that ℛ_*h*_ *>* 1 guarantees the existence of a positive real characteristic root and hence instability of *D*_0_.

#### Proposition 4.2.

*Let F* = *A*_1_, *and V* = −(*A*_0_ + *A*_2_), *where A*_0_, *A*_1_, *A*_2_ *are the matrices appearing in the linearization at the disease-free equilibrium in Theorem 4*.*1. Under the assumptions used in Subsection 4*.*1 to ensure finite cumulative infectious residence times, the threshold* ℛ_*h*_ *admits the equivalent representation*

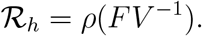

*Moreover*,

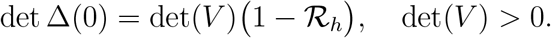

*Consequently, if* ℛ_*h*_ *>* 1, *the characteristic equation possesses at least one positive real root, and hence the disease-free equilibrium D*_0_ *is unstable*.

*Proof*. The matrix *A*_0_ + *A*_2_ is Metzler, since all its off-diagonal entries are non-negative. Its column sums are

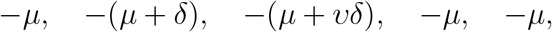

respectively. Hence *V* = −(*A*_0_ + *A*_2_) is a *Z*-matrix and **1**^⊤^*V >* 0, or equivalently, *V* ^⊤^**1** *>* 0. Therefore *V* ^⊤^, and hence *V*, is a nonsingular *M* -matrix. Consequently,

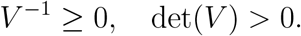

We next relate *V* ^−1^ to the cumulative infectious residence times used in the definition of ℛ_*h*_. Suppress the generation of new infections and consider the disease-transition subsystem

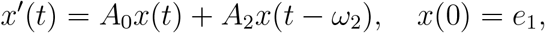

with zero prehistory, where

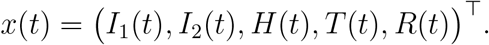

Under the finite-residence-time assumptions of Subsection 4.1, the solution satisfies *x*(*t*) → 0 as *t* → ∞, and its components are integrable on [0, ∞). Integrating the system over [0, ∞) gives

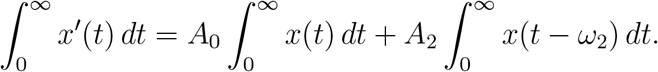

Since *x*(*t*) → 0 and *x*(0) = *e*_1_,

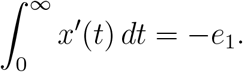

Moreover, because the prehistory is zero,

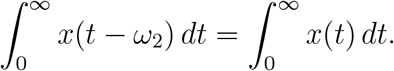

Thus,

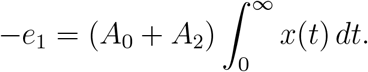

Since *V* = −(*A*_0_ + *A*_2_), it follows that

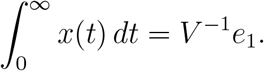

Therefore, the cumulative residence times in the two infectious compartments satisfy

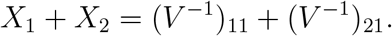

Now define

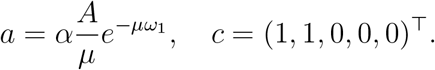

Then *F* = *a e*_1_*c*^⊤^. Using the residence-time definition of ℛ_*h*_ obtained in Subsection 4.1,

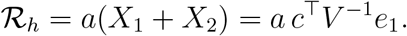

Since *FV* ^−1^ = *a e*_1_*c*^⊤^*V* ^−1^ has rank one, its only possible nonzero eigenvalue is *a c*^⊤^*V* ^−1^*e*_1_. Because *V* ^−1^ ≥0, this eigenvalue is non-negative. Hence

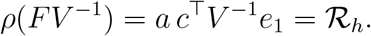

At *σ* = 0, the characteristic matrix satisfies

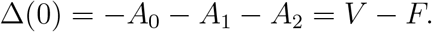

Therefore, by the matrix determinant lemma,

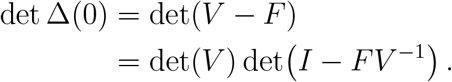

Since *FV* ^−1^ is rank one with nonzero eigenvalue ℛ_*h*_,

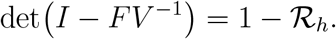

Hence

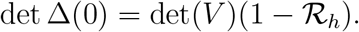

If ℛ_*h*_ *>* 1, then, since det(*V* ) *>* 0, det Δ(0) *<* 0. For real *σ*, the function

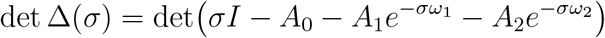

is continuous. Furthermore, as 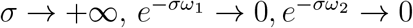, and therefore

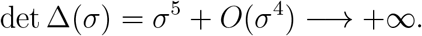

Thus, by the intermediate value theorem, there exists *σ*^*^ *>* 0 such that

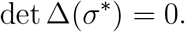

Hence the linearized delay system possesses a positive real characteristic root. By the principle of linearized instability for retarded functional differential equations [43], the disease-free equilibrium *D*_0_ is unstable.□

Thus, whenever the finite-residence-time assumptions hold, the matrix representation ℛ_*h*_ = *ρ*(*FV* ^−1^) coincides with the cumulative infectious residence-time definition of Subsection 4.1. The threshold property itself is independent of that interpretation: the identity det Δ(0) = det(*V* )(1 − ℛ_*h*_) and the instability of *D*_0_ for ℛ_*h*_ *>* 1 follow from the rank-one structure of *F* and the *M* -matrix property of *V* alone.

### 4.3 Existence and stability of endemic equilibrium

The endemic equilibrium, denoted by *D*^*^, is given by the following expression:

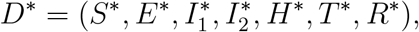

which is obtained by setting all right-hand sides of system (2.1) equal to zero and solving the resulting algebraic system. Using the earlier notations of *q*_1_, *q*_2_, *a*_1_, *a*_2_, *a*_*T*_, *a*_*H*_, and *a*_*R*_, we define

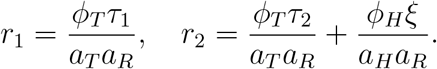

Then the recovered component at equilibrium can be written as

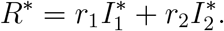

Let

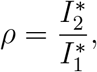

and using the *I*_2_-equilibrium equation, we obtain

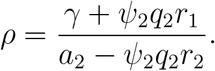

The denominator in the above expression is always positive. Indeed, since

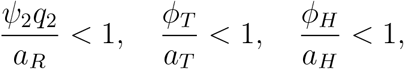

we have

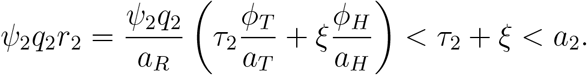

Hence *ρ >* 0 for all positive parameter values. Therefore,

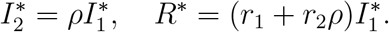

The *I*_1_-equilibrium equation gives

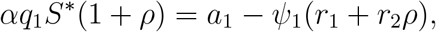

hence,

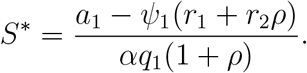

Using the susceptible equilibrium equation, we obtain

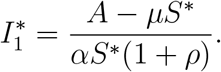

The remaining components are uniquely determined as

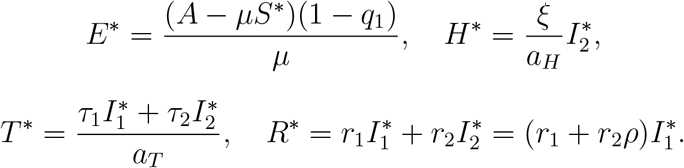

Consequently, components of the endemic equilibrium are given by

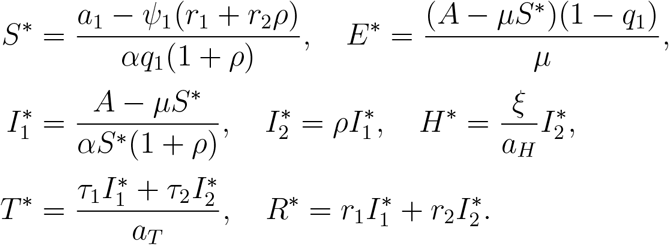

The feasibility of *D*^*^ is determined by the positivity of the infectious components. Since

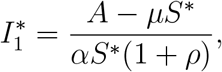

and *α >* 0, *S*^*^ *>* 0, and *ρ >* 0, it follows that 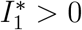 if and only if

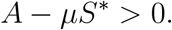

Moreover,

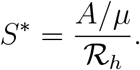

To see this, let 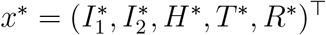. The equilibrium equations for the infected compartments read

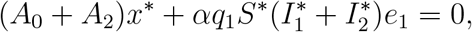

that is,

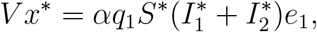

with *V* = −(*A*_0_ + *A*_2_) as in Proposition 4.2. Since *V* is a nonsingular *M* -matrix,

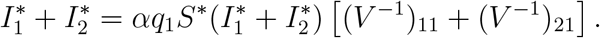

Using the expression for ℛ_*h*_, this becomes

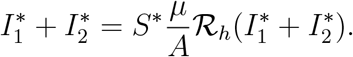

Dividing by 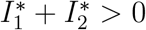 gives

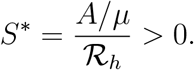

In particular,

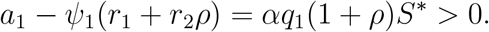

Therefore,

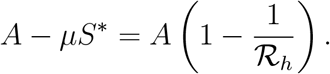

Thus, *A* − *µS*^*^ *>* 0 if and only if ℛ_*h*_ *>* 1. Consequently, 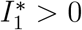 and 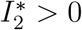 precisely when ℛ_*h*_ *>* 1.

Hence, system (2.1) admits a unique biologically feasible endemic equilibrium whenever ℛ_*h*_ *>* 1. If ℛ_*h*_ = 1, then *A* − *µS* = 0, so 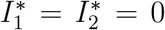, and the endemic equilibrium collapses to the disease-free equilibrium. If ℛ_*h*_ *<* 1, then *A*−*µS*^*^ *<* 0, so no biologically feasible positive endemic equilibrium exists.

To analyze the local asymptotic stability of *D*^*^ for different cases of the time delays *ω*_1_ and *ω*_2_, we obtain the following results.

#### Theorem 4.3.

*The endemic equilibrium D*^*^ *is locally asymptotically stable in the absence of time delays, i*.*e*., *for ω*_1_ = *ω*_2_ = 0, *provided that the Routh-Hurwitz conditions stated in the proof hold*.

#### Theorem 4.4.

*Suppose that ω*_2_ = 0 *and that conditions H1 and H2 hold. Let* 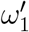 *denote the first positive critical value selected from the family of critical delays* 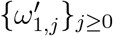. *Then the endemic equilibrium D remains locally asymptotically stable for* 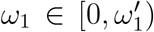. *Moreover, when* 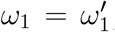, *system* (2.1) *undergoes a Hopf bifurcation at D*^*^, *and a family of periodic solutions bifurcates from D*^*^.

#### Theorem 4.5.

*Suppose that ω*_1_ = 0 *and that conditions H3 and H4 hold. Let* 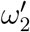 *denote the first positive critical value selected from the family of critical delays* 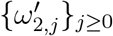. *Then the endemic equilibrium D remains locally asymptotically stable for* 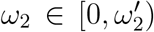. *Moreover, when* 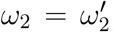, *system* (2.1) *undergoes a Hopf bifurcation at D*^*^, *and a family of periodic solutions bifurcates from D*^*^.

#### Theorem 4.6.

*Suppose that ω*_2_ *is fixed in its stable interval and that conditions H5 and H6 hold. Let* 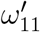 *denote the first positive critical value selected from the family of critical delays* 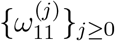. *Then the endemic equilibrium D*^*^ *remains locally asymptotically stable for* 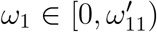. *Moreover, when* 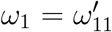, *system* (2.1) *undergoes a Hopf bifurcation at D*^*^, *and a family of periodic solutions bifurcates from D*^*^.

Theorems 4.4-4.6 show that, under the corresponding conditions H1-H6, the endemic equilibrium *D*^*^ is locally asymptotically stable while the relevant delay remains below its first positive critical value. Theorem 4.6 additionally requires *ω*_2_ to be fixed in its stable interval. These conditions guarantee the existence of admissible critical delay values and the transversality condition required for Hopf bifurcation. The detailed definitions of these conditions and the corresponding critical-delay families are given in the proofs in Appendix B.

As the relevant delay passes through its first positive critical value, a pair of characteristic roots crosses the imaginary axis and the system undergoes a Hopf bifurcation at *D*^*^. This gives rise locally to bifurcating periodic solutions. Epidemiologically, this indicates that sufficiently long effective population-level infection-to-transmission delays or delayed relapse timing can generate recurrent epidemic oscillations, rather than simply damping transmission. Such recurrent dynamics are absent from the corresponding non-delayed model under the same endemic-equilibrium stability conditions.

### 4.4 Properties of Hopf bifurcation

In the previous section, we showed that a family of periodic solutions bifurcates from the endemic equilibrium, *D*^*^, for distinct threshold values of the time-delay parameters, *ω*_1_ and *ω*_2_, in system (2.1). Here, we analyze the properties of Hopf bifurcation, particularly the direction and stability of Hopf bifurcation, at the threshold value of the time delay 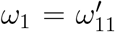 . For this, we utilize the center manifold theorem and normal form theory [45]. Throughout the analysis, it is assumed that 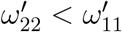, where 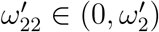.

Assume 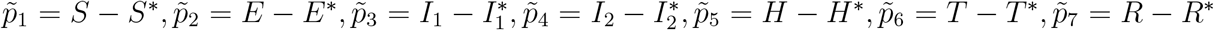 and 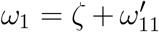, where *ζ* ∈ ℝ. Further, normalize the time delay *ω*_1_ by scaling time *t* as *t* → *t/ω*_1_. Thus, system (2.1) attains the following functional differential form:

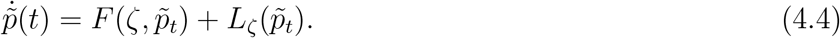

Using similar notations as in Hassard et al. [45], the detailed derivation of the corresponding linear operator formulation, adjoint system, bilinear inner form, eigenvectors, center manifold reduction, and normal form coefficients is presented in Appendix B. In particular, we determine

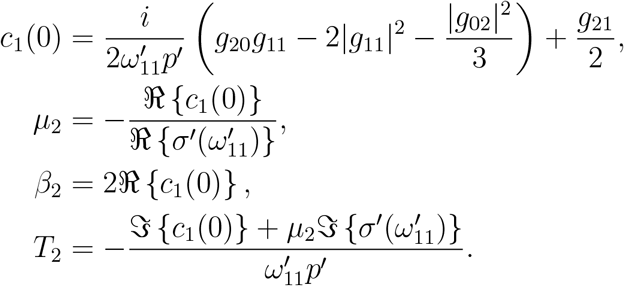

The above terms determine the behavior of bifurcating periodic solutions at the threshold value of the time delay 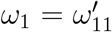 . In particular, *µ*_2_ governs the direction of Hopf bifurcation: if *µ*_2_ *>* 0 (*µ*_2_ *<* 0), then the Hopf bifurcation is supercritical (subcritical) and the bifurcating periodic solution exists for 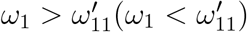. *β*_2_ governs the stability of the bifurcating periodic solution: the bifurcating periodic solution is stable (unstable) if *β*_2_ *<* 0 (*β*_2_ *>* 0). *T*_2_ determines the period of the bifurcating periodic solution: if *T*_2_ *>* 0, the period increases; otherwise, it decreases.

### 4.5 Estimation of the length of time delays to sustain stability

This section determines the length of time delays that sustain the stability of system (2.1) by utilizing the Nyquist criterion [47]. To prove this, system (2.1) and the space of all real-valued continuous functions are considered on [−*ω*, +∞) with the initial conditions provided earlier. Using the ensuing transformations 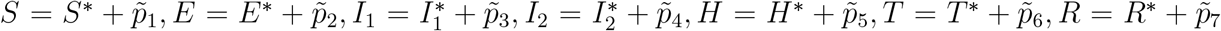, we obtain the linearized system and the corresponding characteristic equation (see Appendix B for the detailed derivation). Local stability of *D*^*^ follows if

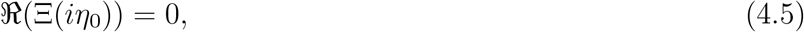

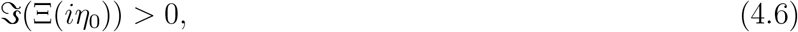

where *η*_0_ is the smallest positive root of Eq. (4.5).

Theorem 4.3 shows that *D*^*^ is locally stable when *ω*_1_ = *ω*_2_ = 0. Hence, by continuity, all eigenvalues have negative real parts for sufficiently small *ω*_1_ *>* 0, *ω*_2_ *>* 0, provided that as *ω*_1_ and *ω*_2_ increase from zero, no eigenvalues with positive real parts bifurcate from infinity. This can be verified by Butler’s lemma [48]. We next consider the case *ω*_1_ *>* 0 and *ω*_2_ in its stable interval, as in Theorem 4.6. If the resulting inequalities (derived in Appendix B) are satisfied simultaneously, then these conditions are sufficient to guarantee stability. We will utilize these to obtain an estimation of the length of the time delay. Our main aim is to find an upper bound *η*^+^ on *η*_0_ independent of *ω*_1_ and *ω*_2_, and then estimate *ω*_1_ so that Eq. (4.6) is satisfied for all values of *η*, 0≤*η*≤*η*^+^, and hence in particular at *η* = *η*_0_. The resulting bound is a sufficient condition. It certifies a delay interval on which stability is guaranteed, without requiring the transcendental characteristic equation to be solved. It is therefore expected to be conservative relative to the exact stability boundary, and its value lies in providing a closed-form guarantee rather than a sharp estimate.

Consequently, we obtain

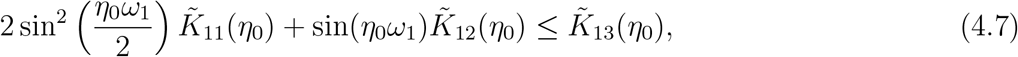

where 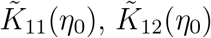,and 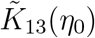 are given in Appendix B. Now, from inequality (4.7), we have

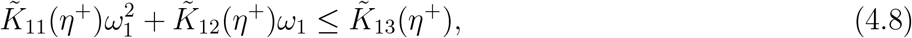

where 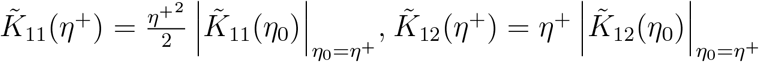, and 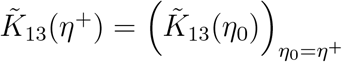. Hence, if

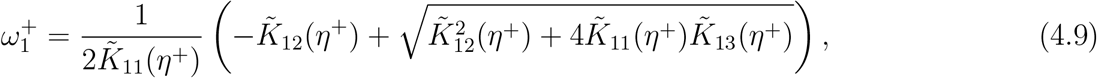

then stability is preserved for 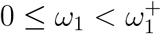.

A similar procedure can be followed to compute the length of the time delay *ω*_2_ for which stability is sustained, considering *ω*_1_ in its stable interval.

### 4.6 Transcritical bifurcation

The previous spectral analysis established local asymptotic stability of the disease-free equilibrium when all characteristic roots of the linearized infection-feedback subsystem lie in the open left half-plane. We now show that the threshold ℛ_*h*_ = 1 corresponds to a forward transcritical bifurcation, considering the transmission coefficient *α* as the bifurcation parameter.

#### Theorem 4.7.

*Assume that all parameters except α are fixed and positive, and that the biologically admissible conditions ensuring finite residence times are satisfied. At the critical value α* = *α*^*^, *equivalently* ℛ_*h*_ = 1, *the characteristic equation of the linearized system at D*_0_ *has a simple zero characteristic root. Moreover, D*_0_ *is unstable whenever* ℛ_*h*_ *>* 1. *If all remaining characteristic roots have negative real parts at* ℛ_*h*_ = 1, *then model* (2.1) *undergoes a forward transcritical bifurcation at D*_0_. *In particular, the endemic equilibrium branch bifurcates from D*_0_ *and is biologically feasible for* ℛ_*h*_ *>* 1.

*Proof*. The human-to-human transmission threshold can be written as

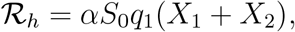

where *X*_1_ and *X*_2_ are the effective residence times in the infectious classes *I*_1_ and *I*_2_, respectively. Here *S*_0_ = *A/µ*, 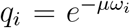, *i* = 1, 2, and the quantities *a*_1_, *a*_2_, *a*_*H*_, *a*_*T*_, *a*_*R*_ are as defined in Section 4.1. Hence the critical transmission coefficient satisfying ℛ_*h*_ = 1 is

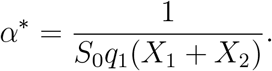

At *D*_0_, the threshold dynamics are determined by the infection-feedback block

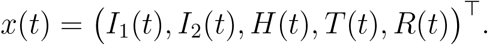

Seeking solutions of the linearized infection subsystem of the form *x*(*t*) = *ve*^*σt*^, and setting *σ* = 0, gives

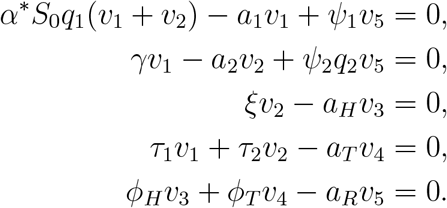

Let

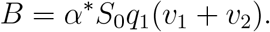

Then the above system is equivalent to

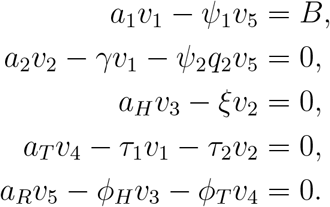

The residence-time quantities *X*_1_, *X*_2_, *X*_*H*_, *X*_*T*_, *X*_*R*_, generated by one unit entry into *I*_1_, satisfy

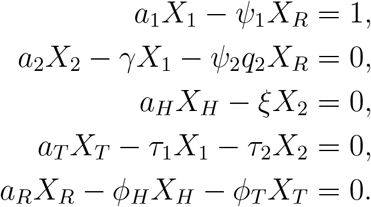

Thus, for input *B*,

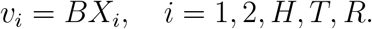

Substituting *v*_1_ = *BX*_1_ and *v*_2_ = *BX*_2_ into the definition of *B* yields

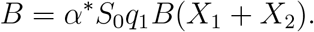

For a nontrivial characteristic vector, *B*≠0. Therefore,

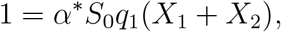

which is equivalent to ℛ_*h*_ = 1. Conversely, if ℛ_*h*_ = 1, then

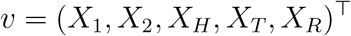

satisfies the characteristic system at *σ* = 0. Hence, *σ* = 0 is a characteristic root if and only if ℛ_*h*_ = 1.

We next show that the zero characteristic root is simple. Near *σ* = 0, the threshold part of the characteristic equation can be written as

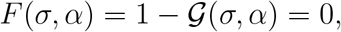

Where

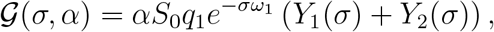

With

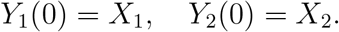

Since *Y*_1_(*σ*) + *Y*_2_(*σ*) is the Laplace transform of the infectious residence-time kernel *k*(*s*) = *I*_1_(*s*) + *I*_2_(*s*), where (*I*_1_, *I*_2_) are the first two components of the solution of the disease-transition subsystem *x*^′^(*t*) = *A*_0_*x*(*t*) + *A*_2_*x*(*t*−*ω*_2_) with *x*(0) = *e*_1_ and zero prehistory, and since this kernel is nonnegative under condition (2.4), there exists a nonnegative kernel *k*(*s*), not identically zero, such that

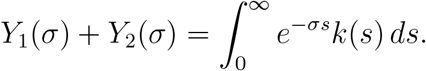

Therefore,

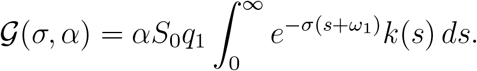

Differentiating with respect to *σ* at (0, *α*^*^) gives

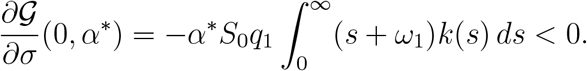

Consequently,

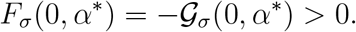

Thus *F*_*σ*_(0, *α*^*^) = 0, and the zero characteristic root is simple. Moreover,

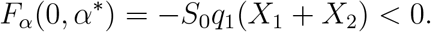

Hence, by the implicit function theorem,

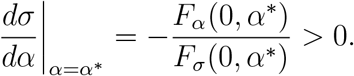

Thus, as *α* increases through *α*^*^, equivalently as ℛ_*h*_ increases through one, the simple characteristic root crosses the imaginary axis from left to right.

To show instability for ℛ_*h*_ *>* 1, note that

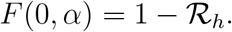

If ℛ_*h*_ *>* 1, then *F* (0, *α*) *<* 0. On the other hand,

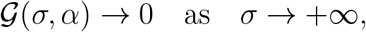

and hence

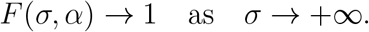

By continuity, there exists *σ*^*^ *>* 0 such that

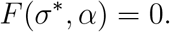

Thus the characteristic equation has a positive real characteristic root, and *D*_0_ is unstable whenever ℛ_*h*_ *>* 1.

Finally, the endemic equilibrium calculation gives

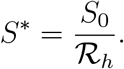

Therefore,

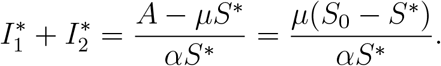

Thus,

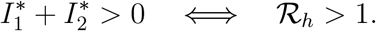

Moreover,

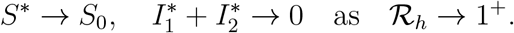

The remaining infected components are determined linearly by 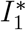 and 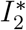, and therefore also tend to zero as ℛ_*h*_ → 1^+^. Hence the endemic branch intersects the disease-free branch at ℛ_*h*_ = 1.

Since the zero root is simple and all remaining characteristic roots are assumed to have negative real parts at ℛ_*h*_ = 1, the center manifold at (*D*_0_, *α*^*^) is one-dimensional. The crossing condition

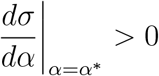

together with the emergence of a positive endemic branch for ℛ_*h*_ *>* 1 implies a forward transcritical bifurcation. Therefore, model (2.1) undergoes a forward transcritical bifurcation at *D*_0_ when ℛ_*h*_ = 1.□

The forward transcritical bifurcation establishes a critical threshold: when ℛ_*h*_ crosses unity, the endemic equilibrium emerges from the disease-free state. Epidemiologically, this means even small increases in transmission, relapse frequency, or infectious residence time can tip the system from disease elimination to endemic persistence. For Nipah virus, this implies that interventions reducing ℛ_*h*_ below unity remain essential for control.

## 5 Model fitting to NiV incidence data and identifiability analysis

### 5.1 Data and fitting procedure

We use NiV incidence data from Bangladesh as the primary dataset for model fitting and parameter estimation. The calibration is used to identify an empirically informed parameter regime for the human-to-human delay model and to evaluate the contribution of relapse and time delays to secondary transmission. The model is not intended to reproduce every year-to-year fluctuation in reported incidence, since sporadic outbreak timing may also be shaped by zoonotic spillover, seasonal exposure, and stochastic introductions that are outside the present model framework. The dataset consists of yearly case counts from 2001 to 2024, reported for Bangladesh and collected from the official website of the Institute of Epidemiology, Disease Control and Research (IEDCR), Bangladesh [49]. These yearly cases, presented in Table 2, serve as the empirical basis for estimating key epidemiological quantities, including the transmission rate, relapse rates, disease-induced death rate, time delays, and the initial infectious populations.

**Table 2:** Number of NiV cases per year (incidence) in Bangladesh from 2001 to 2024 adopted from [49].

| Year | 2001 | 2002 | 2003 | 2004 | 2005 | 2006 | 2007 | 2008 | 2009 | 2010 | 2011 | 2012 |
| --- | --- | --- | --- | --- | --- | --- | --- | --- | --- | --- | --- | --- |
| Cases | 13 | 0 | 12 | 67 | 12 | 0 | 18 | 11 | 4 | 18 | 43 | 17 |
| Year | 2013 | 2014 | 2015 | 2016 | 2017 | 2018 | 2019 | 2020 | 2021 | 2022 | 2023 | 2024 |
| Cases | 31 | 37 | 15 | 0 | 3 | 4 | 8 | 7 | 2 | 3 | 13 | 5 |

The model (2.1) has multiple parameters, some of which are fixed using demographic information, values reported in the NiV literature, or biologically plausible assumptions. These fixed parameter values are provided in Table 3. All rate parameters are expressed in units of year^−1^, since the model is fitted to annual incidence data. The recruitment rate *A* is obtained from the disease-free population balance *A/µ* = *N* (0), i.e., before the emergence of NiV disease. Since the total population of Bangladesh in 2001 was 136,805,810 [50], and 1*/µ* is the average life expectancy, taken to be 73.57 years [28], we have *µ* = 1*/*73.57 and *A* = *N* (0) × *µ* = 1.8595 × 10^6^ individuals per year.

**Table 3:** Fixed numerical values of the parameters used in model (2.1). Mean durations (1*/*rate) are given in brackets.

| Parameter | Numerical value | Unit | Source |
| --- | --- | --- | --- |
| $A$ | $1.8595 \times 10^6$ | individuals $\text{year}^{-1}$ | $\mu \times N(0)$ |
| $\mu$ | $1/73.57$ (73.57 years) | $\text{year}^{-1}$ | [28] |
| $\tau_1$ | 116.32 (3.1 days) | $\text{year}^{-1}$ | Assumed |
| $\gamma$ | 44.26 (8.2 days) | $\text{year}^{-1}$ | [5] |
| $\tau_2$ | 139.44 (2.6 days) | $\text{year}^{-1}$ | [28] |
| $\xi$ | 146 (2.5 days) | $\text{year}^{-1}$ | [28] |
| $\phi_H$ | 26.60 (13.7 days) | $\text{year}^{-1}$ | Assumed |
| $v$ | 0.73 | dimensionless | Assumed |
| $\phi_T$ | 65.04 (5.6 days) | $\text{year}^{-1}$ | Assumed |

The progression rate *γ* from the early infectious stage *I*_1_ to the later symptomatic infectious stage *I*_2_ is fixed at *γ* = 44.26 year^−1^, corresponding to an average progression time of approximately 365*/*44.26 = 8.25 days. This time scale is consistent with the reported clinical progression period for NiV infection [5]. The treatment or clinical-management rates for early-stage and later-stage infectious individuals are fixed at *τ*_1_ = 116.32 year^−1^ and *τ*_2_ = 139.44 year^−1^, respectively. These correspond to average time scales of approximately 365*/*116.32 = 3.14 days and 365*/*139.44 = 2.62 days. The hospitalization rate of symptomatic individuals is fixed at *ξ* = 146 year^−1^, corresponding to an average hospitalization-entry time of 365*/*146 = 2.5 days [28]. The recovery rates from the hospitalized and treatment compartments are fixed at *ϕ*_*H*_ = 26.60 year^−1^ and *ϕ*_*T*_ = 65.04 year^−1^, respectively, corresponding to average recovery times of approximately 13.72 days and 5.61 days. The parameter *v* = 0.73, which scales disease-induced mortality in the hospitalized class, is assumed because precise NiV-specific information for this modifier is not available.

At the beginning of the simulation, the initial values of the system’s state variables are either obtained from demographic information, assumed based on the early outbreak setting, or estimated during model calibration. The initial values of the early-stage infectious individuals *I*_1_(0) and later-stage infectious individuals *I*_2_(0) are estimated together with the epidemiological parameters. The initial value of the exposed individuals *E*(0) is determined by the compatibility condition (2.3) from the estimated parameters and *I*_1_(0), *I*_2_(0), so that the initial data are consistent with the delayed infection term. The susceptible population in 2001 is then given by *S*(0) = *N* (0)−*E*(0) − *I*_1_(0)−*I*_2_(0)−*H*(0)−*T*(0)−*R*(0). Because detailed information on the number of hospitalized and clinically managed individuals at the start of the outbreak is not available, we assume small nonzero baseline values for these compartments, namely *H*(0) = 1 and *T* (0) = 3. These values represent a limited number of severe and clinically managed cases at the beginning of the outbreak and are small relative to the total population size. Since no recoveries have occurred at the initial time, we set *R*(0) = 0.

We estimate the model parameters by solving a bounded least-squares minimization problem using MATLAB’s built-in *lsqnonlin* function [51], which supports parameter estimation within bounded ranges. We select eight quantities for estimation: the transmission rate (*α*), disease-induced death rate (*δ*), relapse rates (*ψ*_1_, *ψ*_2_), effective population-level infection-to-transmission delay (*ω*_1_), post-recovery delay associated with delayed-onset encephalitic complications (*ω*_2_), and the two initial conditions *I*_1_(0) and *I*_2_(0). The initial cumulative-incidence value is fixed at the first reported observation, *C*(0) = *Y*_1_ = 13. A single observation provides no information about the transmission dynamics, so estimating *C*(0) would only reproduce the 2001 count; the 2001 observation is therefore used to set the initial condition and is excluded from the objective. System (2.1) is solved with MATLAB’s *dde23* solver (relative and absolute tolerances 10^−6^ and 10^−8^, maximum step size 0.01 year).

To compare the DDE model output with the reported annual NiV cases, we introduce a cumulative incidence counter *C*(*t*), where

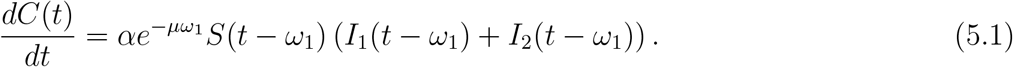

Let *Y*_*i*_, *i* = 1, 2, …, *n*, denote the reported annual NiV cases, and let *Ŷ*_*i*_ denote the corresponding modelpredicted annual cases. Since the first annual incidence value is fixed through *C*(0), the model-predicted annual cases are defined as

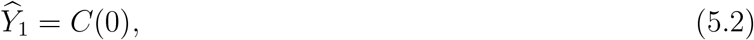

and, for *i* = 2, 3, …, *n*,

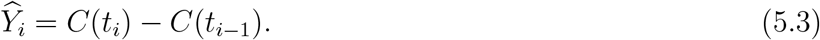

Thus, the first annual case count is set directly by the initial cumulative-incidence counter, while the remaining annual cases are obtained by differencing the cumulative incidence generated by the DDE system. The corresponding model-predicted cumulative cases are given by

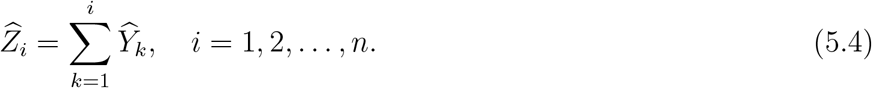

The least-squares objective minimized by *lsqnonlin* is

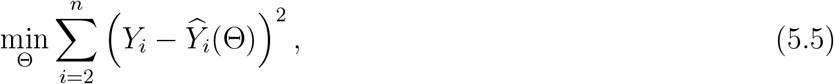

Where

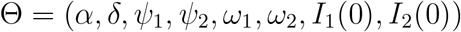

is the vector of estimated quantities, and the sum runs over the *n*−1 = 23 observations from 2002 to 2024. The minimization is restricted to biologically admissible trajectories. Two penalty terms are added to the objective: one proportional to any negative value of the state variables *S, E, I*_1_, *I*_2_, *H, T, R*, and one proportional to any violation of condition (2.4), both evaluated on a grid of 2000 time points over the fitting interval. Both penalties vanish at admissible parameter values, so at the reported estimate the objective equals the sum of squared residuals. Because each evaluation of the objective requires a numerical solution of the DDE system, gradients are approximated by finite differences with relative step 10^−3^, which is well above the solver error. After parameter estimation, the goodness of fit is summarized using the Root Mean Square Error (RMSE), defined by

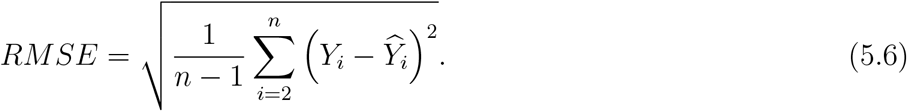

We also compute the Root Mean Square Logarithmic Error (RMSLE),

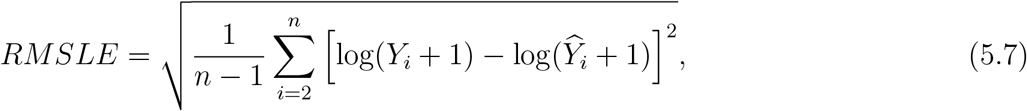

which provides an additional goodness-of-fit measure on a logarithmic scale. The residuals are defined as

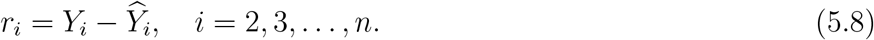

To quantify parameter uncertainty under data variability, we employ a residual bootstrap procedure with 1000 replicates. Residuals from the fitted model are first centered by subtracting their mean. For each replicate, residuals are sampled with replacement and added to the fitted annual incidence values to generate a synthetic dataset, and the model is refitted using the same bounded least-squares procedure, starting from the estimate. A replicate in which the optimizer terminated within 0.1% of its starting point in every parameter was refitted from up to three randomly perturbed starting points, and the fit with the smallest objective was retained; replicates that remained at the starting point were excluded. For each replicate, *E*(0) is recomputed from condition (2.3) and ℛ_*h*_ from the fitted parameters. We report 95% bootstrap confidence intervals for all estimated parameters, taken as the 2.5th and 97.5th percentiles of the resulting parameter distributions.

For each estimated quantity we show the bootstrap distribution as a histogram with the fitted estimate and confidence limits marked. Using the calibrated model, we then project NiV cases in Bangladesh through 2030, propagating uncertainty by simulating forward for each of the bootstrap parameter sets and taking the 95% band as the 2.5th and 97.5th percentiles of the resulting trajectories.

### 5.2 Parameter estimates and uncertainty

The fitted model captures the broad temporal pattern of the NiV cases in Bangladesh (Figure 2(a)), with SSE = 4030.893, RMSE = 13.238, and RMSLE = 1.180. The residual error largely reflects yearto-year irregularity in reported case counts; individual outbreak years are driven by discrete spillover events and localized transmission clusters, which a deterministic population-level model does not resolve. The model is therefore intended to reproduce the overall level and timing of transmission rather than the timing and magnitude of individual annual peaks. Under the calibrated model assumptions, the fitted trajectory displays a secondary rise that peaks around 2023-2024, after the decline of the main outbreak phase, and then decreases through 2030. This suggests that relapse and delay mechanisms can produce a later recurrent rise in incidence rather than a simple monotone decline after the main outbreak phase. Because the model does not include repeated zoonotic introductions, this recurrence, produced jointly by the delays and the relapse pathway, may partly absorb spillover-driven incidence; the projected resurgence therefore reflects human-to-human transmission dynamics only. The residuals fluctuate around zero without systematic trend, with the largest values confined to years of abrupt outbreak peaks (Figure 2(b)). We also plot the reported and model-predicted cumulative cases to visualize the epidemic burden implied by the fitted annual cases (Figure 2(c)). During calibration, trajectories were satisfied condition (2.4) and remain non-negative. Both properties were evaluated along the calibrated solution on a grid of 10,000 time points with tighter solver tolerances (relative and absolute tolerances 10^−8^ and 10^−10^), and the minimum value of

**Figure 2:**
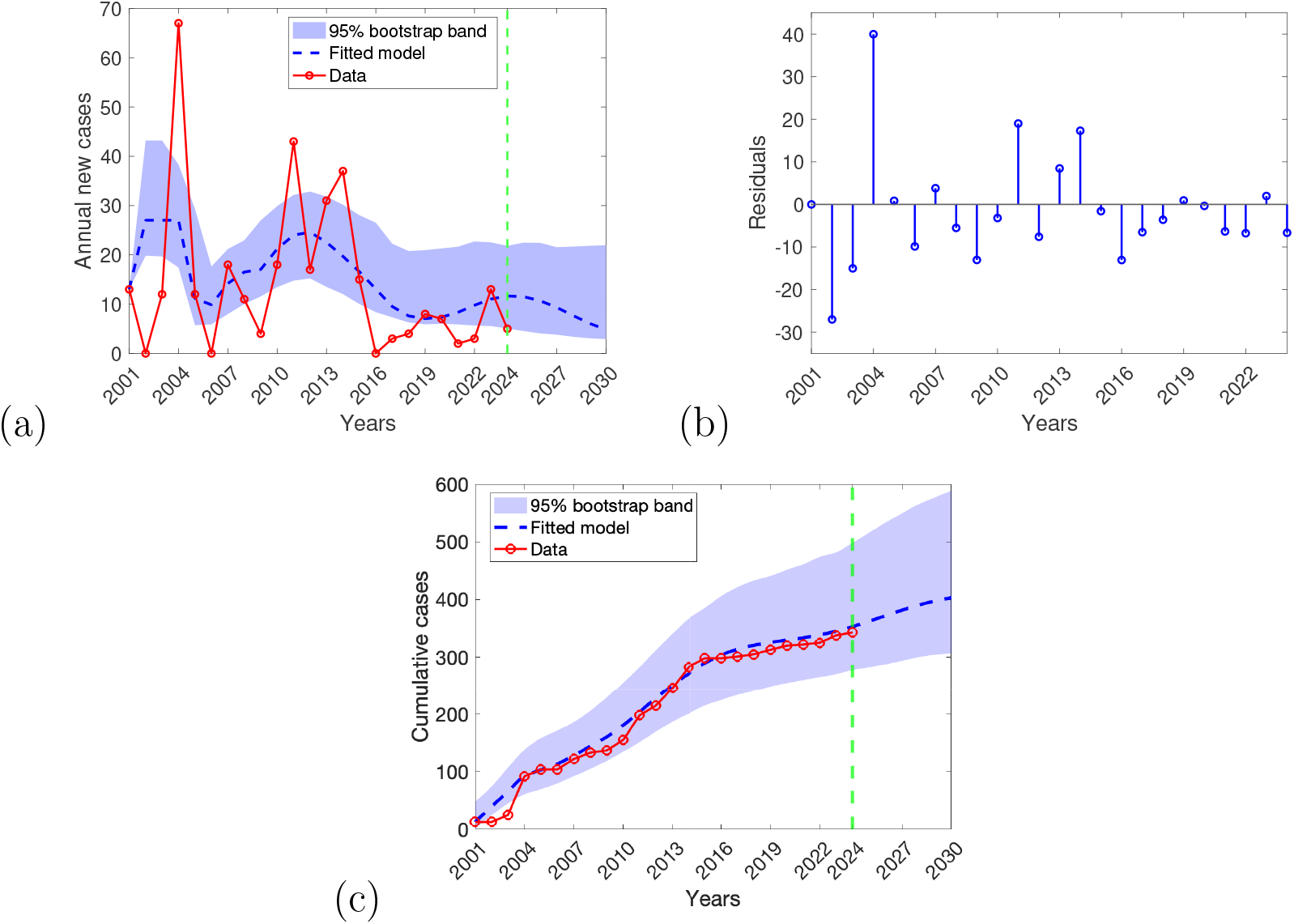
Data fitting of model (2.1) using annual reported cases of NiV disease in Bangladesh from 2001 to 2024. (a) Least-squares fit of model (2.1), showing the model-generated annual new cases (blue dashed curve) compared with the reported annual NiV cases in Bangladesh (red circles), with the model projected to 2030. The blue shaded region represents the 95% bootstrap confidence band for the model trajectory, obtained from 1000 residual bootstrap replicates reflecting uncertainty in the fitted curve. The vertical green dashed line indicates the end of the fitting period and the beginning of the prediction period. (b) Residuals of the fitted model, computed as the difference between the reported and model-predicted annual cases. (c) Cumulative reported and model-predicted cases, computed from the annual case counts over time, with the corresponding 95% bootstrap confidence band.

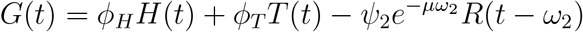

remained non-negative throughout the calibration interval. For the calibrated solution, the minimum admissibility margin was min_*t*_ *G*(*t*) = 0.0094 *>* 0, confirming that condition (2.4) was satisfied along the calibrated trajectory. All state variables also remained non-negative, with the smallest infectious value min_*t*_ *I*_2_(*t*) = 0.032, and both properties continued to hold along the projection to 2030.

The estimated parameter values and their 95% bootstrap confidence intervals are reported in Table 4. Figure 3 presents the empirical bootstrap distributions of the estimated parameters, of *E*(0) and of ℛ_*h*_. In each panel, the red vertical line represents the least-squares estimate, whereas the black dashed vertical lines indicate the lower and upper limits of the corresponding 95% bootstrap confidence interval. The distributions of *α, ψ*_1_, *ω*_1_ and *ω*_2_ are centered near their least-squares estimates, indicating stable estimation under residual resampling. The bootstrap distributions of *δ, E*(0), *I*_1_(0) and *I*_2_(0) are right-skewed, whereas that of *ψ*_2_ is broad, indicating asymmetric uncertainty. However, all bootstrap distributions remain unimodal and concentrated around the fitted estimates. The broad distribution of *ψ*_2_ is consistent with the practical non-identifiability of the relapse rates discussed in Section 5.3. The estimated human reproduction number is ℛ_*h*_ = 0.826, with 95% bootstrap confidence interval (0.738, 1.026), and 96.0% of the bootstrap replicates give ℛ_*h*_ *<* 1. The data therefore support a human reproduction number below one, consistent with sporadic, spillover-driven outbreaks that do not sustain human-to-human transmission on their own.

**Table 4:** Estimated parameter values in model (2.1) and their 95% residual-bootstrap confidence intervals. *E*(0) is derived from the compatibility condition (2.3), and ℛ_*h*_ is computed from the estimated parameters.

| Parameters | Estimated value | 95% bootstrap confidence interval |
| --- | --- | --- |
| $\alpha$ | $1.237 \times 10^{-7}$ | $(0.892 \times 10^{-7}, 1.692 \times 10^{-7})$ |
| $\delta$ | 9.072 | (5.226, 15.846) |
| $\psi_1$ | 1.712 | (0.832, 2.593) |
| $\psi_2$ | 3.721 | (1.732, 4.632) |
| $\omega_1$ | 3.247 | (2.540, 4.108) |
| $\omega_2$ | 4.302 | (2.660, 5.845) |
| $I_1(0)$ | 0.781 | (0.118, 1.802) |
| $I_2(0)$ | 0.887 | (0.226, 2.324) |
| $E(0)$ | 89.70 | (61.52, 140.51) |
| $\mathcal{R}_h$ | 0.826 | (0.738, 1.026) |

**Figure 3:**
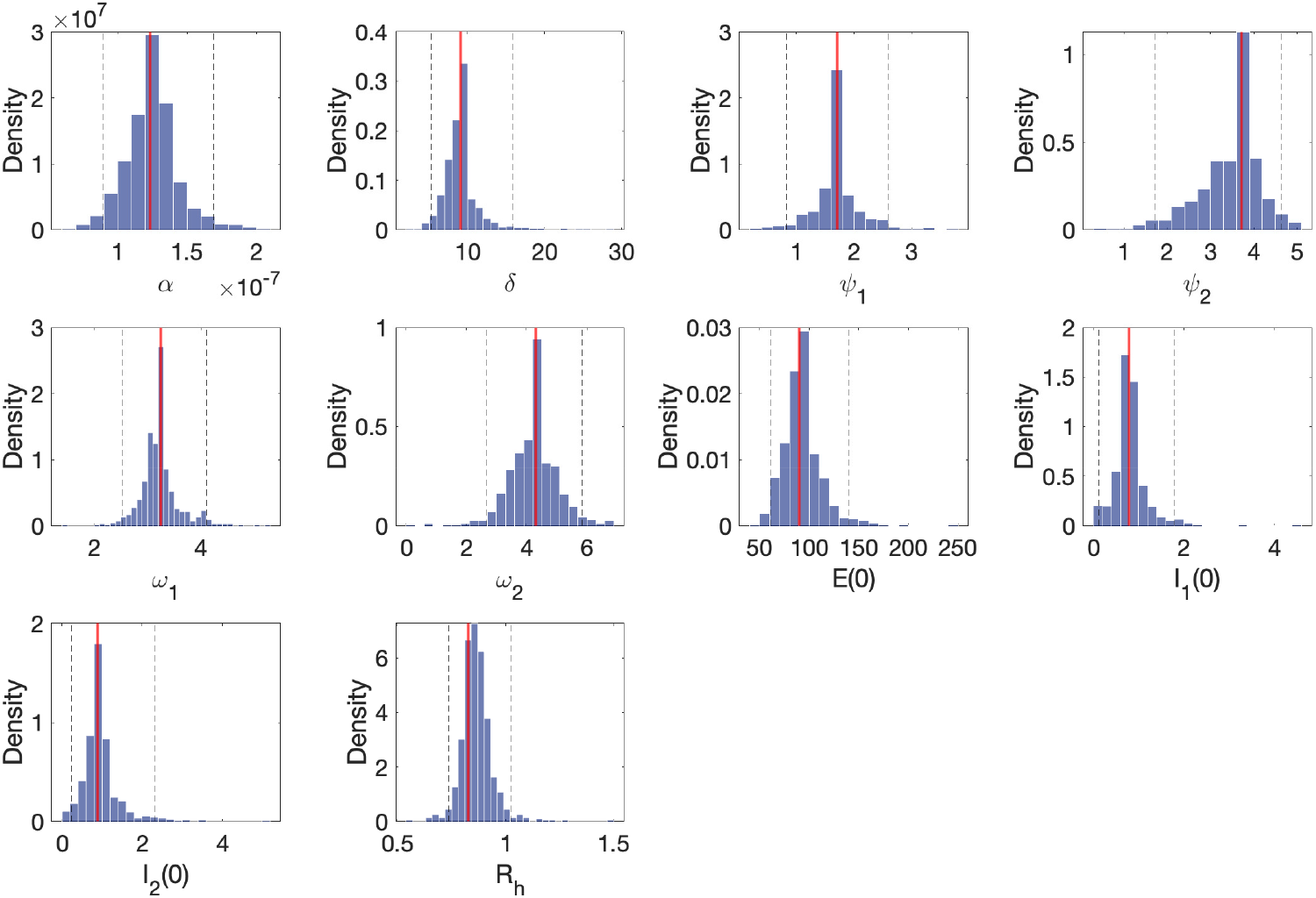
Bootstrap distributions of the estimated parameters, of the initial exposed population *E*(0) (derived from condition (2.3)) and of the reproduction number ℛ_*h*_, obtained from 1000 residual bootstrap replicates. The histograms show the empirical distributions of the bootstrap estimates. The red vertical line represents the least-squares estimate, while the black dashed vertical lines represent the lower and upper limits of the 95% bootstrap confidence interval (2.5th and 97.5th percentiles).

### 5.3 Identifiability analysis

We assessed the identifiability of the reduced parameter set Θ = {*α, δ, ψ*_1_, *ψ*_2_, *ω*_1_, *ω*_2_, *I*(0)}, where *I*(0) = *I*_1_(0) + *I*_2_(0) with the split fixed at the ratio obtained in the primary fit, and *E*(0) is determined by compatibility condition (2.3). The two initial infectious classes enter the force of infection and condition (2.3) only through their sum, and both classes are left within days (mean residence time of about two days in *I*_1_); annual incidence therefore constrains *I*(0) = *I*_1_(0) + *I*_2_(0) = 1.669 but not its split between the two classes, and the separate values of *I*_1_(0) and *I*_2_(0) in Table 4 should be read as one admissible split of this total. Since symbolic structural identifiability methods are not readily applicable to delay differential equations, we combined a local sensitivity analysis with profile likelihood [52].

#### Local sensitivity and Fisher information

All seven parameters influence the annual incidence appreciably, with *α* and *δ* the most influential (Figure 4(a)). The Fisher information matrix (FIM) of the annual incidence data has full rank at the estimate, so all seven quantities are locally identifiable in the linearized sense. Its smallest eigenvalue is, however, 1.5 × 10^−5^ of the largest (condition number 6.7 × 10^4^), revealing a single weak direction (Figure 4(c)). This direction has loadings (+0.40, +0.52, −0.48, −0.43, −0.40) on (log *α*, log *δ*, log *ψ*_1_, log *ψ*_2_, log *I*(0)) and on *ω*_1_ and *ω*_2_, along it, higher transmission is partly compensated by faster disease-induced removal, reduced relapse and a smaller initial infectious population. Consistently, the collinearity index of the full parameter set is 115, whereas every six-parameter subset that omits one of these five parameters has a collinearity index below 15. Pairwise collinearity indices are all small (Figure 4(b)), confirming that the compensation involves these parameters jointly rather than any single pair.

**Figure 4:**
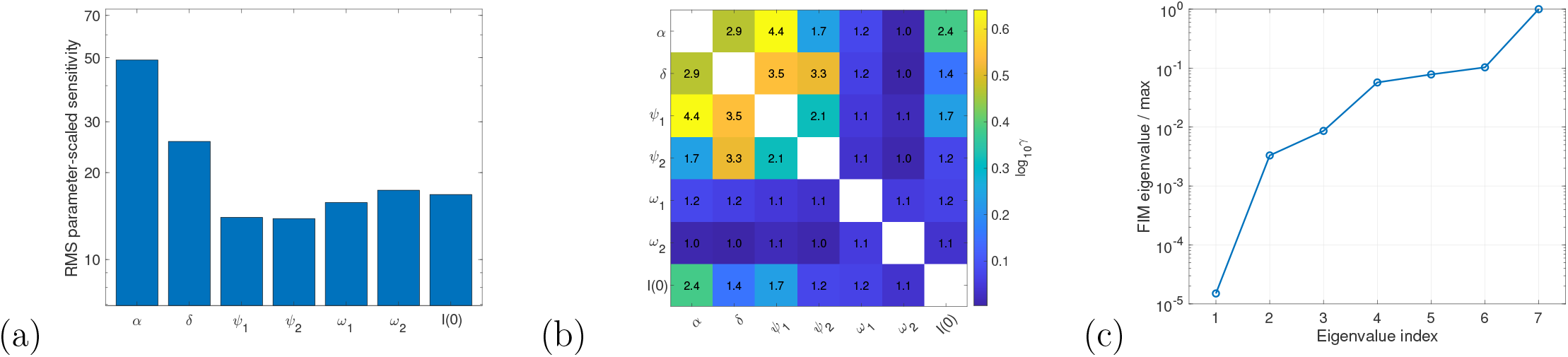
Local sensitivity analysis at the estimate for the reduced parameter set, where *I*(0) = *I*_1_(0)+*I*_2_(0) with the fraction *I*_1_(0)*/I*(0) fixed at its estimated value. (a) Root-mean-square parameter-scaled sensitivity of annual incidence (logarithmic scale). (b) Pairwise collinearity indices *γ* (values in the cells; colour shows log_10_ *γ*). All values are below 4.5, well under the commonly used threshold of 15, so no pair of parameters is strongly collinear. (c) Eigenvalues of the Fisher information matrix relative to the largest. The smallest eigenvalue (1.5×10^−5^) identifies a single weakly identified parameter direction; since no pair of parameters is strongly collinear, this direction involves more than two parameters.

#### Profile likelihood

Profile likelihoods (Figure 5, Table 5) show that *α, δ, ω*_1_, *ω*_2_ and *I*(0) are practically identifiable, with finite 95% confidence intervals, whereas the relapse rates are not, as the profile of *ψ*_1_ remains below the 95% threshold above its estimate, and that of *ψ*_2_ below its estimate, throughout the admissible range. The data therefore bound these parameters on one side only, giving *ψ*_1_ ≥ 0.401 and *ψ*_2_ ≤ 4.338 at the 95% level (Table 5). The profile of *ω*_2_ levels off moderately above the threshold for *ω*_2_ *<* 2 years, so its lower bound is less sharply determined than the other bounds. Along several profiles, a marginally better fit was found (at most Δ(−2 log *L*) =−0.18, below 5% of the 95% threshold), which does not affect any confidence interval. Annual incidence data therefore do not constrain the relapse rates, and their individual values should not be interpreted as epidemiological estimates.

**Table 5:** Profile-likelihood practical identifiability of the reduced parameter set with *I*(0) = *I*_1_(0) + *I*_2_(0). “Open” indicates that the profile did not cross the 95% threshold 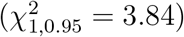 before reaching the end of the explored range. The last column gives the range of ℛ_*h*_ over all parameter sets in each profile’s 95% confidence region.

| Parameter | 95% profile CI | Identifiable | $\mathcal{R}_h$ range |
| --- | --- | --- | --- |
| $\alpha$ | $(0.339, 1.416) \times 10^{-7}$ | yes | (0.673, 0.866) |
| $\delta$ | (2.488, 10.456) | yes | (0.793, 0.876) |
| $\psi_1$ | (0.401, open) | no (upper side) | (0.804, 0.947) |
| $\psi_2$ | (open, 4.338) | no (lower side) | (0.770, 0.900) |
| $\omega_1$ | (2.638, 4.153) | yes | (0.818, 0.873) |
| $\omega_2$ | (2.491, 5.570) | yes | (0.786, 0.868) |
| $I(0)$ | (0.679, 2.184) | yes | (0.825, 0.937) |

**Figure 5:**
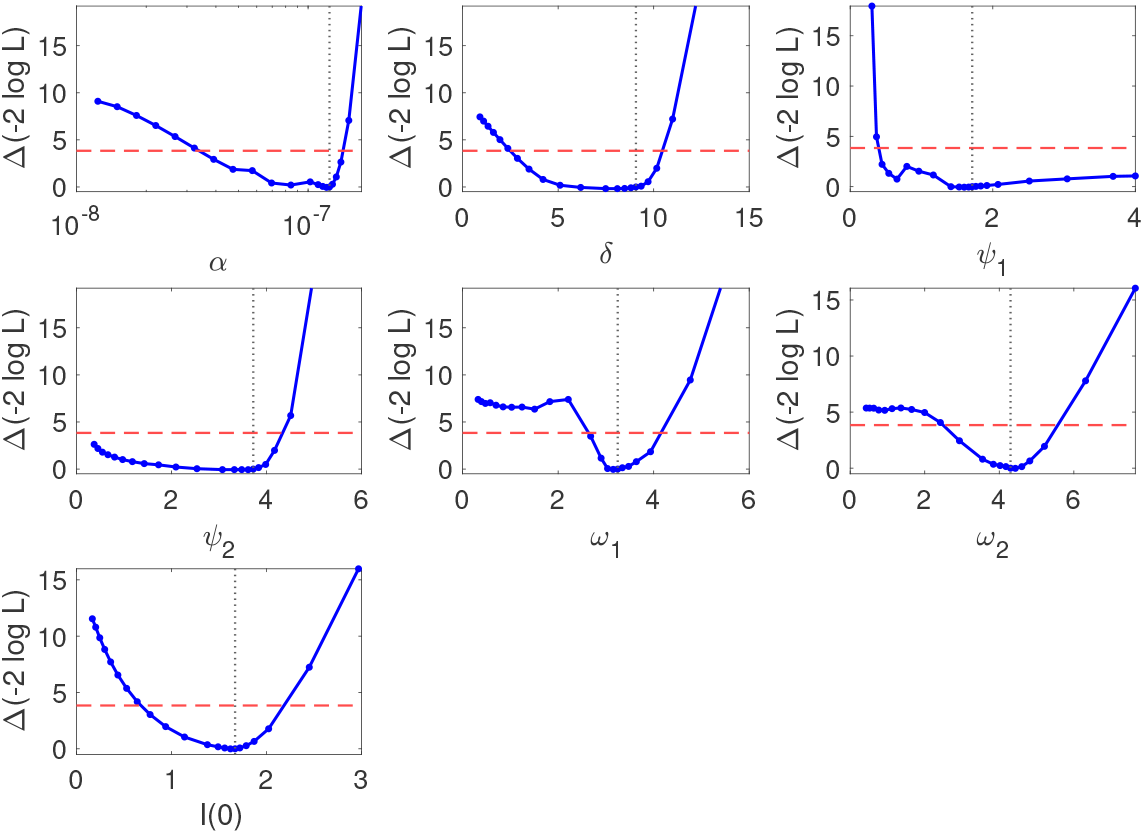
Profile likelihoods of the reduced parameter set, with *I*(0) = *I*_1_(0) + *I*_2_(0). Each panel shows Δ(−2 log *L*) as the indicated parameter is fixed on a grid and all others are re-optimized (*α* on a logarithmic scale). The dashed red line marks the 95% threshold 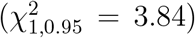 and the dotted vertical line the estimate. The profiles of *α, δ, ω*_1_, *ω*_2_ and *I*(0) exceed the threshold on both sides of the estimate, so these parameters are practically identifiable; the profiles of *ψ*_1_ and *ψ*_2_ remain below the threshold on one side (upper and lower, respectively), so the relapse rates are not practically identifiable.

#### Robustness of ℛ_*h*_

The gradient of log ℛ_*h*_ is nearly orthogonal to the weak FIM direction (| cos *θ*| = 0.002), and across the 95% confidence regions of all seven profiles ℛ_*h*_ ranges from 0.67 to 0.95, remaining below unity throughout. Together with the bootstrap analysis (96% of replicates with ℛ_*h*_ *<* 1), this shows that the conclusion of subcritical human-to-human transmission is robust to the practical nonidentifiability of the relapse parameters. This orthogonality explains why the two findings are compatible: a derived quantity can be well determined even when its constituent parameters are not, provided its gradient has negligible projection onto the weakly determined direction. To first order, the relapse rates can therefore vary along that direction with little change in ℛ_*h*_, and the profile ranges above confirm that this holds over the full 95% confidence regions rather than only locally.

## 6 Numerical simulation

To explore the epidemic dynamics of NiV under varying relapse rates and time delays, we conduct a series of numerical simulations across alternative epidemiological scenarios, including different levels of transmission intensity and clinical-management-related rates. Numerical solutions of system (2.1) are obtained using MATLAB’s built-in delay differential equation solver <monospace>dde23</monospace>, which is well suited for systems with discrete time delays [51,53]. Unless otherwise stated, simulations use the fixed parameter values in Table 3 together with the estimates in Table 4. To examine the sensitivity of epidemic outcomes to key mechanisms, we systematically vary selected parameters around these baseline values while holding all others fixed, allowing us to identify parameter regimes associated with heightened or reduced disease burden.

### 6.1 Impact of relapse and time delays on long-term NiV incidence

To examine how relapse and time delays shape epidemic trajectories, we vary the relapse rate *ψ*_1_, the effective population-level infection-to-transmission delay *ω*_1_, and the delayed relapse time *ω*_2_ by ±10%, ±20%, and ±30% around their baseline estimated values, while holding all other parameters fixed. Model outputs are evaluated using time-series dynamics and cumulative incidence over 2001-2030.

The three mechanisms act on different phases of the epidemic (Figure 6). Varying *ψ*_1_ leaves the initial outbreak essentially unchanged but scales the late-period hump monotonically, since relapse can contribute only after a recovered population has accumulated. In contrast, *ω*_1_ affects both the timing and height of all epidemic waves, reflecting its direct control over entry into infectious transmission; larger values of *ω*_1_ delay and amplify the later peaks. Varying *ω*_2_ shifts the timing of later incidence fluctuations with little effect on their magnitude, consistent with its role in determining when, rather than how many, recovered individuals return to the infectious compartments. Thus, *ω*_1_ shapes the entire trajectory, whereas *ψ*_1_ and *ω*_2_ govern the magnitude and timing of post-peak dynamics, respectively.

**Figure 6:**
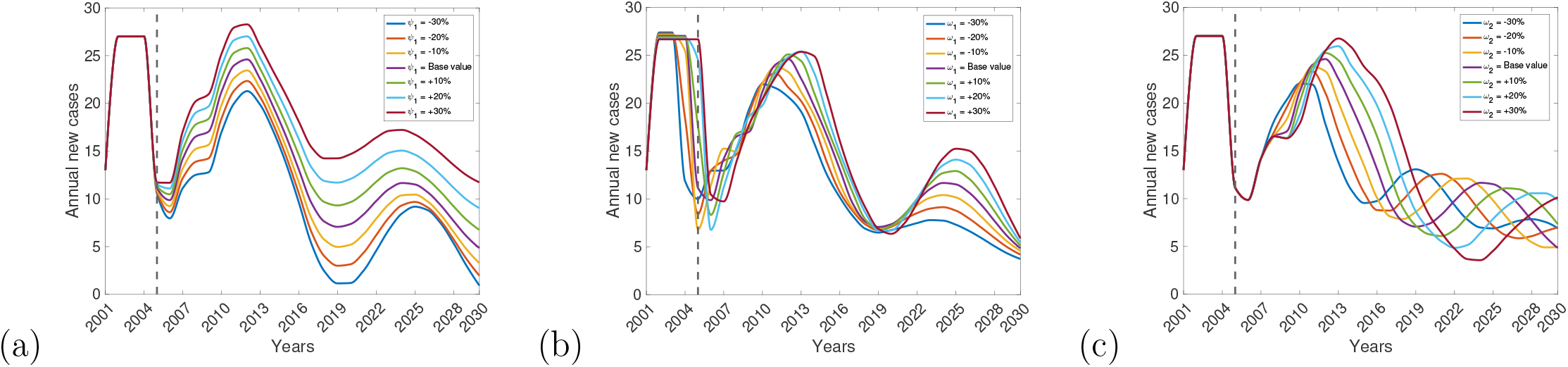
Time-series sensitivity analysis of annual NiV cases from 2001 to 2030 under variations in the (a) immediate relapse rate *ψ*_1_, (b) effective population-level infection-to-transmission delay *ω*_1_, and (c) delayed relapse time *ω*_2_. Each parameter is varied by ±10%, ±20%, and ±30% relative to its baseline value, while all other parameters are held fixed. The vertical dashed line indicates the approximate year after which the scenario trajectories begin to separate.

To quantify the long-term impact of relapse and delay-related parameters, we computed the change in cumulative simulated NiV cases over 2001-2030 relative to the calibrated baseline scenario. The immediate relapse rate *ψ*_1_ has the largest effect: a 30% reduction lowers cumulative incidence by 21.64%, corresponding to approximately 92.4 cases averted, while reductions of 10% and 20% yield decreases of 8.07% and 15.27%, respectively. Increases in *ψ*_1_ produce a monotonic rise in cumulative incidence, with a 30% increase producing approximately 30.05% more cases relative to baseline (Figure 7). The effective population-level infection-to-transmission delay *ω*_1_ also acts monotonically in the same direction, but with a weaker effect than *ψ*_1_. A 30% reduction in *ω*_1_ decreases cumulative cases by 14.90%, corresponding to approximately 63.6 cases averted, whereas a 30% increase raises cumulative cases by 10.94%. Changes in the delayed-relapse time *ω*_2_ have the smallest effect across the same perturbation range: a 30% reduction decreases cumulative cases by 5.84%, while a 30% increase raises cumulative cases by 4.54%. These results indicate that *ψ*_1_ exerts the strongest influence on cumulative incidence, followed by *ω*_1_, with *ω*_2_ having the weakest effect. Absolute cumulative case counts under each scenario are provided in Appendix C (Figure 16 and Tables 7, 8, and 9).

**Figure 7:**
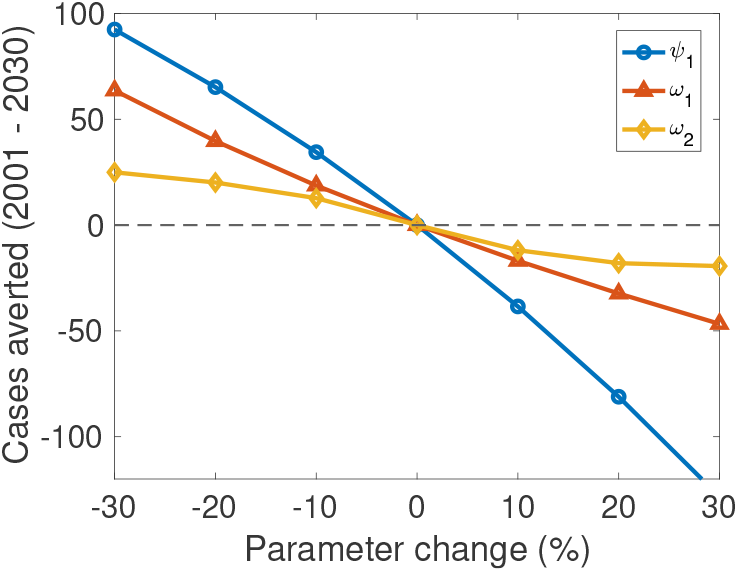
Total simulated NiV cases averted over 2001-2030 under variations in key epidemiological parameters. Cases averted are computed relative to the baseline scenario as a function of the immediate relapse rate (*ψ*_1_), the effective population-level infection-to-transmission delay (*ω*_1_), and the delayed-relapse time (*ω*_2_). Positive values indicate fewer simulated cases than baseline, while negative values indicate additional simulated cases relative to baseline.

To examine how relapse-related parameters influence infectious replenishment, we performed a sensitivity analysis in which the total relapse rate 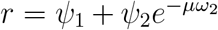 was varied while all other parameters were fixed at their estimated values. For each value of *r*, we computed the corresponding relapse contribution fraction and ℛ_*h*_ (Figure 8); both increase with *r*. At low values of *r*, ℛ_*h*_ remains close to its value without relapse (ℛ_*h*_≈0.12), so that transmission-generated infections decline rapidly; the relapse fraction, although of order 0.4, then describes the composition of a small and vanishing infectious inflow. As *r* increases, the relative contribution of relapse-mediated return to the infectious compartments becomes progressively larger, and ℛ_*h*_ approaches its calibrated value 0.826 only at the fitted relapse rates.

**Figure 8:**
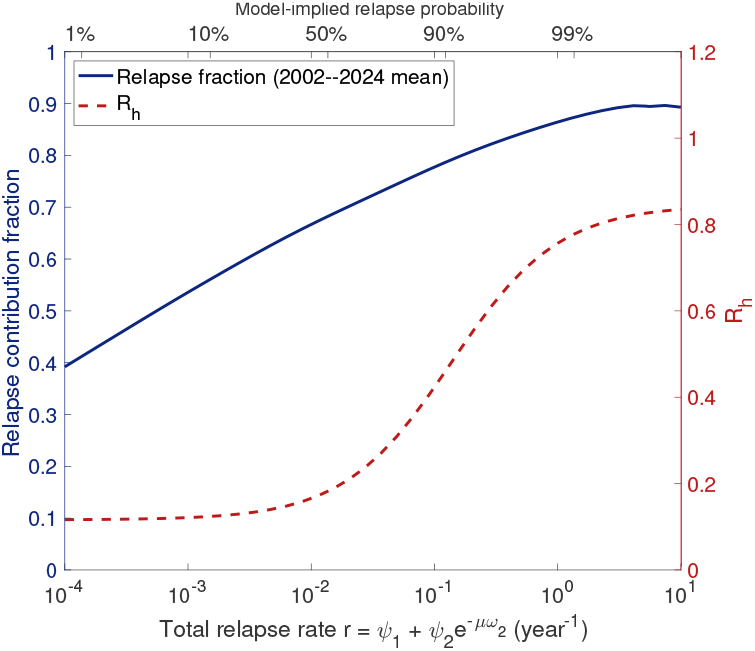
Sensitivity of the relapse contribution fraction and ℛ_*h*_ to the total relapse rate 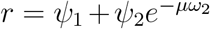 . The relapse contribution fraction increases strongly with the total relapse rate, indicating that the high relapse contribution inferred in the fitted model is driven by the relapse terms. This quantity should be interpreted as an effective model-derived infectious-replenishment contribution, rather than as a direct estimate of clinically documented relapse frequency.

The high relapse fraction under the calibrated baseline regime therefore reflects a change in the composition of infectious inflow rather than direct evidence for frequent clinical relapse. Clinically documented relapse or late-onset neurological disease after Nipah virus infection has been reported, but its frequency is not well quantified across outbreak settings, and the available quantitative evidence comes mainly from limited outbreak-specific data. For example, Tan et al. [20] analysed relapsed and late-onset encephalitis during the 1998-1999 Malaysia outbreak, a setting that may not be directly transferable to Bangladesh-type Nipah virus transmission. A recent systematic review further emphasized that post-acute sequelae after Nipah virus infection remain incompletely documented, with additional research needed in Bangladeshi and Indian outbreak settings [21]. The relapse contribution reported here should therefore be read as a model-derived effective contribution to infectious replenishment. Within the model, relapse-mediated return acts as an effective replenishment mechanism that can support persistence of infectious dynamics even when primary human-to-human transmission alone is insufficient to sustain epidemic growth; because repeated zoonotic introductions are not included, this mechanism, together with the delays, may partly absorb spillover-driven recurrence in the calibrated fit.

### 6.2 Impact of relapse and time delays under alternative epidemiological scenarios

The joint perturbations reveal a consistent asymmetry between the early and later epidemic phases. The initial outbreak peak is governed largely by transmission and progression to infectiousness, whereas the later humps are increasingly shaped by relapse. Figure 9 illustrates this pattern for five parameter pairs, with each parameter varied by ±20% around its baseline value. The shaded region represents the minimum-maximum envelope of trajectories generated over the corresponding scenario grid, while the solid curve denotes the baseline solution. These envelopes represent deterministic perturbation scenarios rather than a quantification of parameter uncertainty.

**Figure 9:**
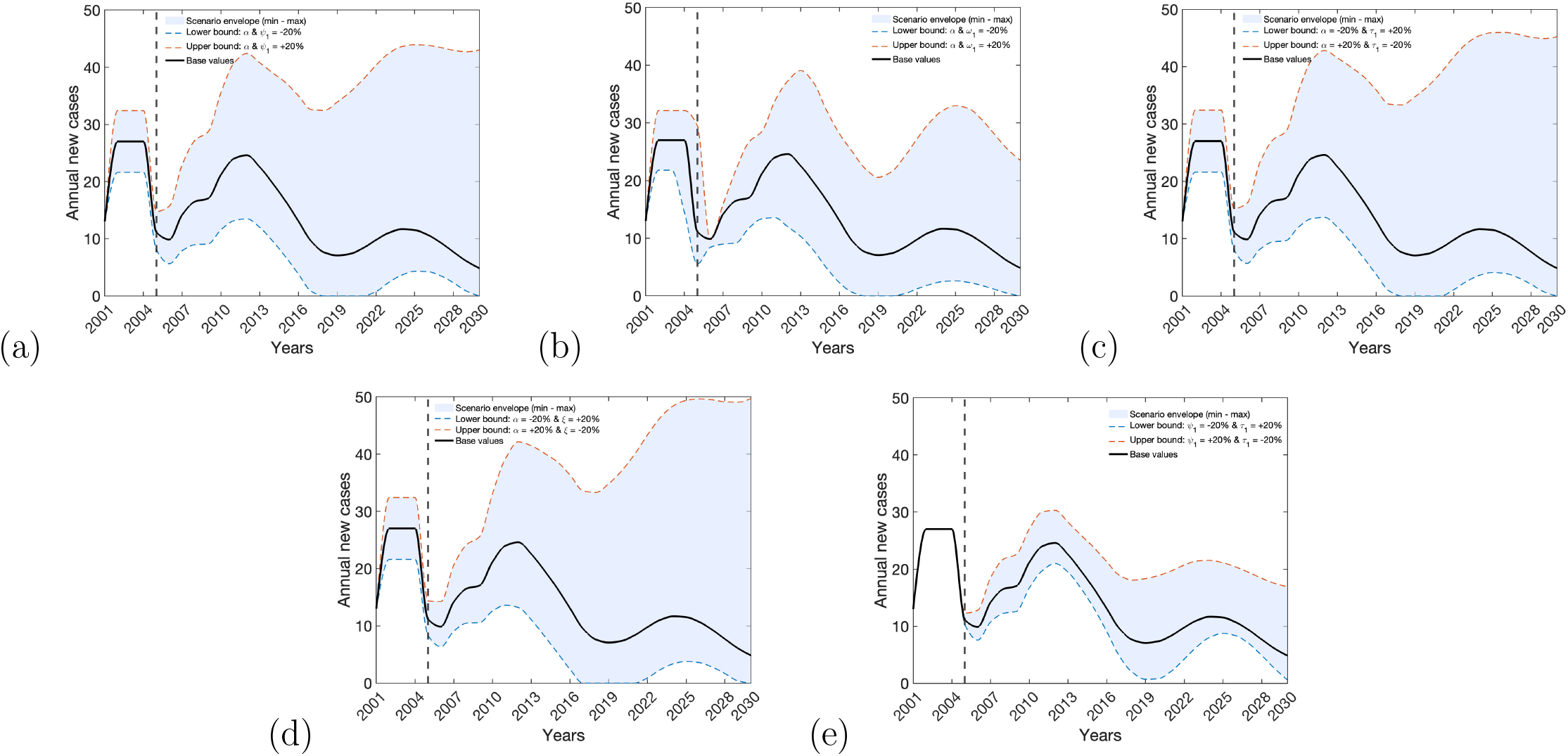
Joint sensitivity envelopes showing the combined effects of paired epidemiological parameters on simulated annual NiV cases. Each parameter pair is varied by ±20% around its baseline value. The shaded regions represent the minimum-maximum envelopes of the resulting annual case trajectories, while the solid black curve denotes the baseline scenario. Panels (a)-(e) correspond to simultaneous variation in: (a) the transmission rate *α* and immediate relapse rate *ψ*_1_; (b) *α* and the effective population-level infection-to-transmission delay *ω*_1_; (c) *α* and the treatment or clinical-management rate *τ*_1_ of early-stage infectious individuals; (d) *α* and the hospitalization rate *ξ* of later-stage infectious individuals; and (e) *ψ*_1_ and *τ*_1_. The vertical dashed line indicates the approximate year after which the scenario trajectories begin to diverge.

The asymmetry is most evident in Figure 9(e), where joint variation of the immediate relapse rate *ψ*_1_ and the early-stage clinical-management rate *τ*_1_ leaves the initial trajectory nearly unchanged but visibly alters the later humps. This pattern is consistent with relapse becoming epidemiologically relevant only after a recovered population has accumulated. By contrast, varying the transmission rate *α* jointly with either *ψ*_1_ or *ω*_1_ (Figures 9(a) and 9(b)) affects both the early and later phases. Pairing a reduced *α* with an increased *τ*_1_ or hospitalization rate *ξ* (Figures 9(c) and 9(d)) lowers the initial peak and attenuates the later humps. Across the parameter pairs, the trajectory envelopes widen after the initial outbreak phase, suggesting that later epidemic dynamics are more sensitive to parameter variation than the early trajectory. The individual trajectories underlying these envelopes are provided in Appendix C, Figure 17.

### 6.3 Sensitivity analysis of ℛ_*h*_

To identify which parameters most influence the human-to-human transmission threshold ℛ_*h*_, we conducted a global sensitivity analysis using partial rank correlation coefficients (PRCCs) with Latin hypercube sampling (LHS). PRCCs measure the strength and direction of the monotonic association between each parameter and ℛ_*h*_ after controlling for the remaining sampled parameters. A total of *N* = 10,000 valid LHS samples were generated. Each parameter was sampled independently from a uniform distribution over 50%-150% of its baseline value, except *v*, which was restricted to its biologically admissible range. The parameter ranges used in the LHS analysis are reported in Table 6. Statistical significance of the PRCCs was assessed using two-sided *t*-tests with Bonferroni correction for fifteen comparisons. To check numerical stability, we recomputed the PRCCs using half of the LHS sample; the maximum absolute change was max |PRCC_*N*_ − PRCC_*N/*2_| = 0.0110, indicating stable PRCC estimates.

**Table 6:** Parameter ranges and PRCC results for the LHS-based global sensitivity analysis of ℛ_*h*_ using *N* = 10,000 valid samples. Each parameter was sampled independently from a uniform distribution over 50%-150% of its baseline value, except *v*, for which the upper bound was restricted to its biologically admissible maximum of 1. Units are as in Tables 3 and 4. Adjusted *p*-values are Bonferroni-corrected for fifteen comparisons.

| Parameter | Baseline | Sampling range | PRCC | Adjusted $p$-value |
| --- | --- | --- | --- | --- |
| $\alpha$ | $1.237 \times 10^{-7}$ | $[6.185 \times 10^{-8}, 1.856 \times 10^{-7}]$ | +0.888 | $< 10^{-4}$ |
| $A$ | $1.859 \times 10^6$ | $[9.297 \times 10^5, 2.789 \times 10^6]$ | +0.887 | $< 10^{-4}$ |
| $\mu$ | 0.01359 | $[6.796 \times 10^{-3}, 0.02039]$ | -0.896 | $< 10^{-4}$ |
| $\omega_1$ | 3.247 | [1.623, 4.870] | -0.080 | $< 10^{-4}$ |
| $\omega_2$ | 4.302 | [2.151, 6.453] | +0.049 | $< 10^{-4}$ |
| $\tau_1$ | 116.3 | [58.16, 174.5] | -0.505 | $< 10^{-4}$ |
| $\tau_2$ | 139.4 | [69.72, 209.2] | +0.307 | $< 10^{-4}$ |
| $\gamma$ | 44.26 | [22.13, 66.39] | -0.323 | $< 10^{-4}$ |
| $\xi$ | 146 | [73, 219] | -0.649 | $< 10^{-4}$ |
| $\delta$ | 9.072 | [4.536, 13.61] | -0.814 | $< 10^{-4}$ |
| $\phi_H$ | 26.6 | [13.3, 39.9] | +0.692 | $< 10^{-4}$ |
| $\phi_T$ | 65.04 | [32.52, 97.56] | +0.021 | 0.6026 |
| $v$ | 0.73 | [0.365, 1] | -0.664 | $< 10^{-4}$ |
| $\psi_1$ | 1.712 | [0.856, 2.568] | +0.524 | $< 10^{-4}$ |
| $\psi_2$ | 3.721 | [1.861, 5.582] | -0.496 | $< 10^{-4}$ |

**Table 7:** Effect of varying the immediate relapse rate *ψ*_1_ on total simulated NiV cases over 2001-2030.

| % change | $\psi_1$ | Cumulative cases | Cases averted | Reduction (%) |
| --- | --- | --- | --- | --- |
| -30 | 1.198 | 334.55 | 92.41 | 21.644 |
| -20 | 1.370 | 361.76 | 65.195 | 15.270 |
| -10 | 1.541 | 392.49 | 34.466 | 8.072 |
| Base value | 1.712 | 426.96 | 0 | 0 |
| +10 | 1.883 | 465.40 | -38.446 | -9.005 |
| +20 | 2.054 | 508.08 | -81.124 | -19.000 |
| +30 | 2.226 | 555.25 | -128.29 | -30.048 |

**Table 8:** Effect of varying the effective population-level infection-to-transmission delay *ω*_1_ on total simulated NiV cases over 2001-2030.

| % change | $\omega_1$ | Cumulative cases | Cases averted | Reduction (%) |
| --- | --- | --- | --- | --- |
| -30 | 2.273 | 363.34 | 63.617 | 14.900 |
| -20 | 2.598 | 387.25 | 39.701 | 9.299 |
| -10 | 2.922 | 408.27 | 18.683 | 4.376 |
| Base value | 3.247 | 426.96 | 0 | 0 |
| +10 | 3.572 | 443.84 | -16.882 | -3.954 |
| +20 | 3.896 | 459.29 | -32.329 | -7.572 |
| +30 | 4.221 | 473.65 | -46.697 | -10.937 |

**Table 9:** Effect of varying the delayed-relapse time *ω*_2_ on total simulated NiV cases over 2001-2030.

| % change | $\omega_2$ | Cumulative cases | Cases averted | Reduction (%) |
| --- | --- | --- | --- | --- |
| -30 | 3.011 | 402.01 | 24.942 | 5.842 |
| -20 | 3.442 | 406.84 | 20.117 | 4.712 |
| -10 | 3.872 | 414.22 | 12.735 | 2.983 |
| Base value | 4.302 | 426.96 | 0 | 0 |
| +10 | 4.732 | 438.83 | -11.872 | -2.781 |
| +20 | 5.162 | 444.96 | -18.004 | -4.217 |
| +30 | 5.593 | 446.34 | -19.385 | -4.540 |

The results separate into parameters that increase and parameters that decrease the human-to-human transmission threshold (Figure 10(a)). The transmission rate *α*, recruitment rate *A*, and immediate relapse rate *ψ*_1_ show positive associations with ℛ_*h*_. The strong positive effects of *α* and *A*, together with the strong negative effect of *µ*, are consistent with their combined role through the factor *αA/µ* in the expression for ℛ_*h*_. For *ψ*_1_, the positive association reflects the increased total expected infectious residence time produced by repeated return from the recovered compartment to the early infectious class. The recovery rate of hospitalized individuals *ϕ*_*H*_ is also positively associated, since higher *ϕ*_*H*_ increases the probability that hospitalized individuals survive to enter the recovered class, from which relapse returns them to infectiousness. Similarly, the treatment rate *τ*_2_ has a positive association with ℛ_*h*_, because increased movement from the severe infectious class into recovery can enlarge the pool available for relapse-mediated return.

**Figure 10:**
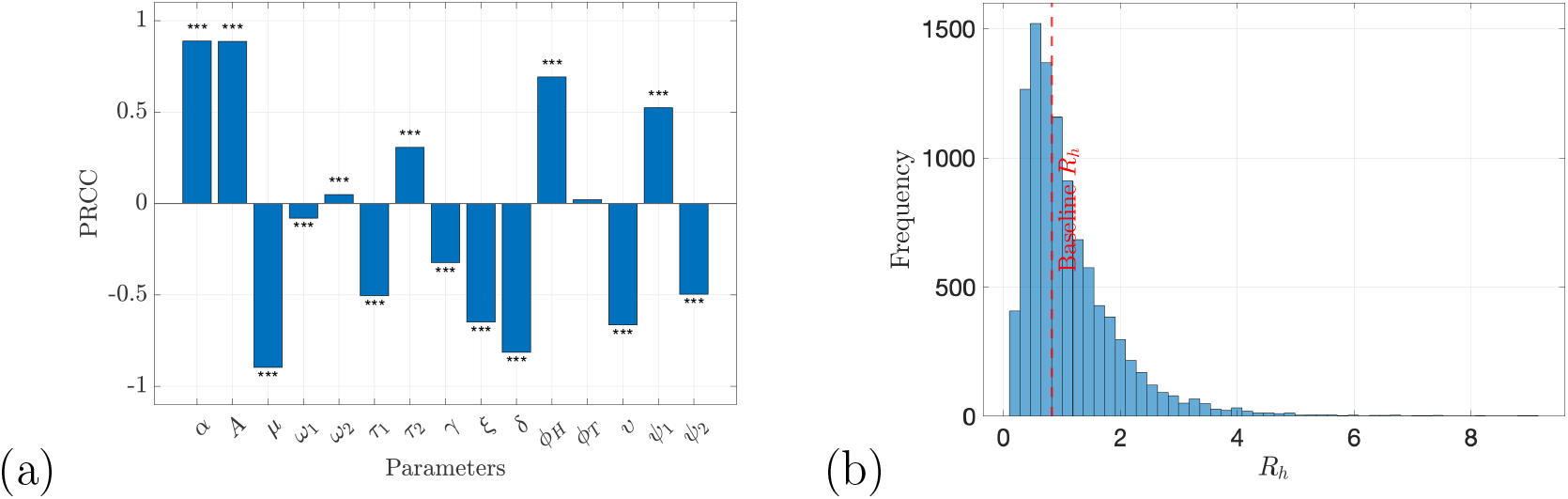
PRCC-based global sensitivity analysis of ℛ_*h*_ using Latin hypercube sampling with *N* = 10,000 valid samples. Parameters were sampled independently from uniform distributions over 50%-150% of their baseline values, with biologically admissible bounds imposed where needed; the resulting ranges are reported in Table 6. (a) Partial rank correlation coefficients (PRCCs) between each parameter and ℛ_*h*_. Statistical significance was assessed using two-sided *t*-tests with Bonferroni correction for fifteen comparisons: ^*, **^, and ^***^ denote adjusted *p <* 0.05, *p <* 0.01, and *p <* 0.001, respectively; ns denotes not significant. Recomputing the PRCCs using half of the LHS sample changed the estimates by at most 0.011. (b) Distribution of ℛ_*h*_ across the LHS samples. The red dashed vertical line marks the baseline value ℛ_*h*_ = 0.826, and 43% of samples produced ℛ_*h*_ *>* 1.

Conversely, the natural mortality rate *µ*, disease-induced death rate *δ*, early-stage clinical-management rate *τ*_1_, hospitalization rate *ξ*, progression rate *γ*, disease-induced mortality modifier *v*, and delayed relapse rate *ψ*_2_ show negative associations. These effects are consistent with their roles in shortening infectious residence times, moving individuals away from transmission-producing states, or altering the balance of recovery and relapse-mediated return. The negative PRCC for *ψ*_2_ should be interpreted in this conditional PRCC sense: after controlling for the other sampled parameters, increasing the delayed relapse rate is associated with a lower ℛ_*h*_ over the sampled parameter space, reflecting the nonlinear dependence of ℛ_*h*_ on the relapse pathway and the delayed survival factor. The delay parameters are comparatively unimportant: *ω*_1_ has only a weak negative association, entering ℛ_*h*_ through the survival factor *e*^−*µω*1^, whereas *ω*_2_ shows a weak positive association. After Bonferroni correction, all parameters except *ϕ*_*T*_ remain statistically significant.

For the estimated baseline parameter values, we obtain ℛ _*h*_≈0.826 *<* 1, indicating that the calibrated human-to-human transmission process is subcritical under the baseline assumptions. The distribution of ℛ_*h*_ under simultaneous parameter variation is shown in Figure 10(b). Across the *N* = 10,000 LHS samples, ℛ_*h*_ ranges from 0.101 to 9.114, with median 0.890 and mean 1.112; 43.23% of sampled parameter sets produce ℛ_*h*_ *>* 1. This broad spread should be interpreted as an exploratory global sensitivity result under simultaneous 50%-150% variation of all sampled parameters, including demographic parameters such as *A* and *µ*, rather than as a direct measure of data-based uncertainty in ℛ_*h*_. In contrast, the data-based uncertainty analyses remain below the threshold in most cases: 96% of bootstrap replicates give ℛ_*h*_ *<* 1, and ℛ_*h*_ ranges from 0.67 to 0.95 across the profile-likelihood analyses. Thus, the baseline and data-supported estimates indicate subcritical human-to-human transmission, while the LHS analysis identifies which parameter directions could shift the system toward supercritical behavior under broader exploratory perturbations.

The condition ℛ_*h*_ *<* 1 does not contradict the delayed incidence peaks observed in the simulations. The threshold ℛ_*h*_ characterizes the local invasion capacity of infection near the disease-free equilibrium, whereas the simulated trajectories over 2001-2030 also reflect the initial conditions, discrete delays, and relapse-mediated returns to the infectious compartments. Consequently, relapse and delay mechanisms can prolong transient transmission and generate later recurrent incidence peaks over a finite time horizon even when the baseline value of ℛ_*h*_ remains below unity.

We further assess the joint effects of selected parameter pairs on ℛ_*h*_ using two-parameter contour plots. Figure 11(a) shows that ℛ_*h*_ increases with both the effective transmission rate *α* and the immediate relapse rate *ψ*_1_. The oblique orientation of the threshold contour ℛ_*h*_ = 1 indicates a compensatory interaction between these parameters: higher values of *ψ*_1_ reduce the increase in *α* required for ℛ_*h*_ to exceed unity. At the baseline value of *ψ*_1_, an approximately 21% increase in *α* is required to cross the threshold ℛ_*h*_ = 1, whereas a 30% increase in *ψ*_1_ lowers the required increase in *α* to approximately 10%. Because *ψ*_1_ is bounded only from below by the data (Section 5.3), these percentages characterize the local geometry of the ℛ_*h*_ surface at the calibrated parameter set rather than data-supported estimates of invasion risk. Within the model, immediate relapse enhances invasion potential by increasing relapse-mediated replenishment of the infectious classes.

**Figure 11:**
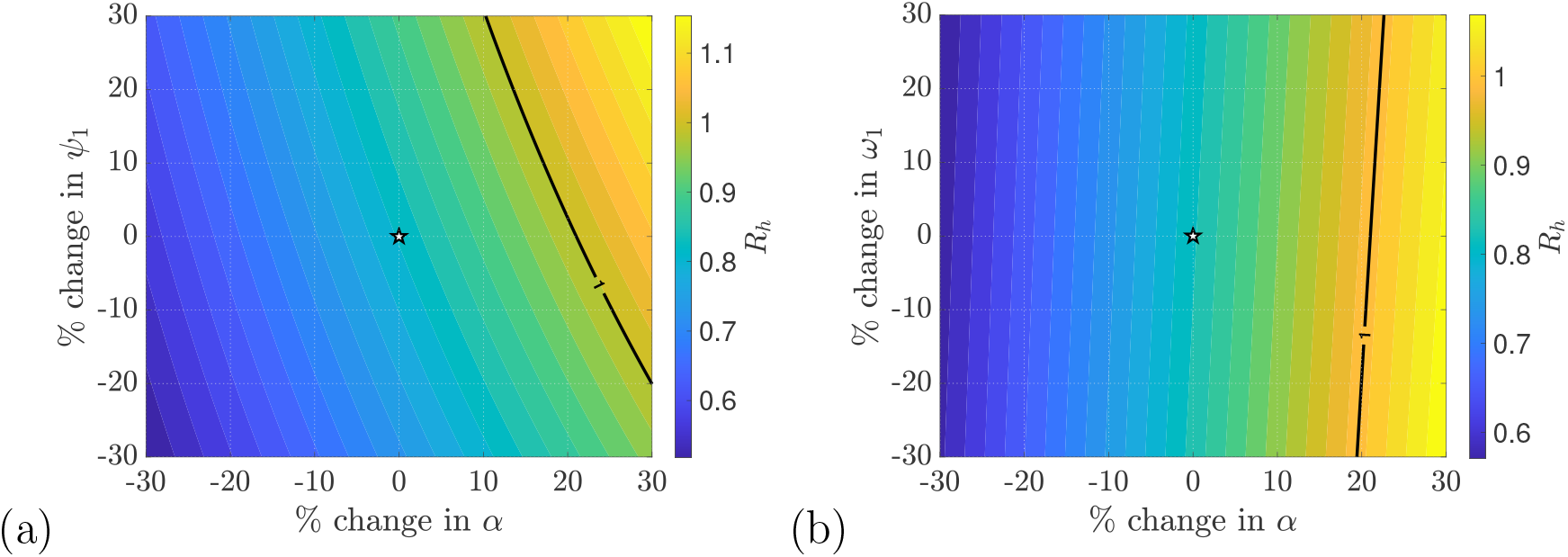
Two-parameter contour plots of the human-to-human transmission threshold ℛ_*h*_. Panel (a) shows the variation of ℛ_*h*_ under simultaneous changes in the effective human-to-human transmission rate *α* and the immediate relapse rate *ψ*_1_, while panel (b) shows the corresponding variation with respect to *α* and the effective population-level infection-to-transmission delay *ω*_1_. The colour scale represents the magnitude of ℛ_*h*_, and the solid black contour denotes the threshold ℛ_*h*_ = 1, separating the subcritical region (ℛ_*h*_ *<* 1) from the supercritical region (ℛ_*h*_ *>* 1). The star indicates the calibrated baseline parameter combination, corresponding to ℛ_*h*_ ≈ 0.826.

Figure 11(b) shows that *α* remains the dominant determinant of ℛ_*h*_ when varied jointly with *ω*_1_. The nearly vertical threshold contour indicates that variation in *ω*_1_ has comparatively little influence on ℛ_*h*_. Increasing *ω*_1_ shifts the threshold contour slightly toward larger values of *α*, consistent with the reduction in ℛ_*h*_ produced by the survival factor 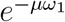. Across the explored ±30% range of *ω*_1_, the critical value of *α* changes only modestly. In both panels, the baseline parameter combination lies within the subcritical region ℛ_*h*_ *<* 1.

### 6.4 Numerical simulations of Hopf bifurcation induced by time delays

To investigate the broader long-term dynamical behavior permitted by the model, including sustained oscillations and Hopf bifurcation, we consider an illustrative parameter regime distinct from the calibrated baseline values. The parameters are selected to satisfy the biological feasibility conditions and to place the endemic equilibrium near a stability boundary. We then systematically vary the effective population-level infection-to-transmission delay *ω*_1_ and the relapse-associated delay *ω*_2_ to characterize the asymptotic behavior of the system under alternative delay scenarios. The numerical simulations show a transition from a stable endemic equilibrium to sustained oscillatory dynamics as the delay parameters cross their respective critical values, consistent with the Hopf bifurcation conditions obtained from the theoretical analysis. Thus, these results characterize the qualitative dynamical capacity of the model under a hypothetical scenario.

Unless otherwise stated, the illustrative parameter values are:

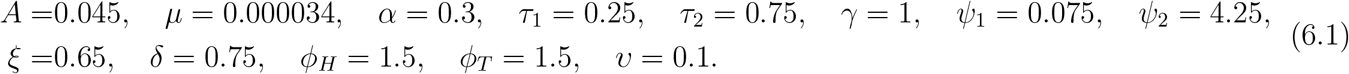

For *ω*_1_ = 0, newly infected individuals enter the early infectious class *I*_1_ directly, consistent with system (2.1) and the analysis in Theorem 4.5.

Figure 12 shows the effect of varying relapse rate *ψ*_1_ on the total infected population *I*_1_ + *I*_2_ in the absence of explicit delays, i.e. *ω*_1_ = *ω*_2_ = 0. The trajectories exhibit short-term damped oscillations before converging to endemic steady states. As *ψ*_1_ increases, the long-term level of *I*_1_ + *I*_2_ also increases, indicating that relapse from the recovered class into the early infectious compartment *I*_1_ enhances disease persistence. This occurs because a larger *ψ*_1_ replenishes the early infectious stage and thereby prolongs the infectious pathway. Thus, increasing *ψ*_1_ increases the endemic disease burden but does not destabilize the endemic equilibrium.

**Figure 12:**
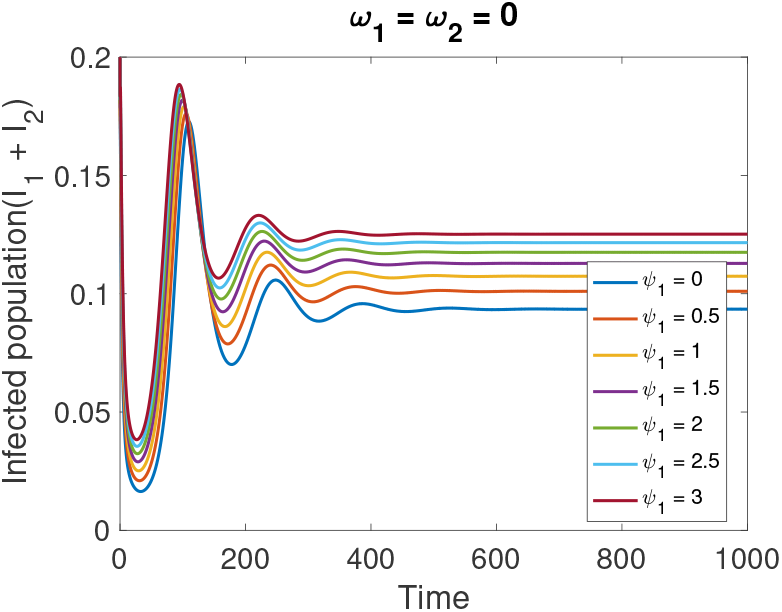
Effect of varying the relapse rate *ψ*_1_ on the total infected population *I*_1_ + *I*_2_ in the no-delay case (*ω*_1_ = *ω*_2_ = 0). Increasing *ψ*_1_ raises the long-term infected burden. Although damped oscillations occur during the initial transient phase, the trajectories converge to endemic steady states, and no sustained oscillations are observed over the parameter range considered. All remaining parameters are fixed at the illustrative values specified in Eq. (6.1).

Figure 13 illustrates the dynamics of the total infectious population (*I*_1_ + *I*_2_) under different delay configurations, with each simulation started from a small perturbation of *D*^*^. The critical delays were obtained from the characteristic equation at *D*^*^. When *ω*_2_ = 0 (Figure 13(a)), *D*^*^ loses stability at 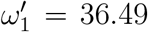. When *ω*_2_ = 10 (Figure 13(b)), *D*^*^ loses stability at 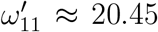. Similarly, increasing *ω*_2_ destabilizes *D*^*^ at 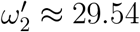 when *ω*_1_ = 0 (Figure 13(c)) and at 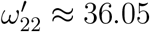 when *ω*_1_ = 10 (Figure 13(d)). Below each critical delay, trajectories return to *D*^*^; beyond it, oscillations emerge, indicating a Hopf bifurcation. The numerical simulations align with the previously derived theoretical results for Hopf bifurcation in each instance.

**Figure 13:**
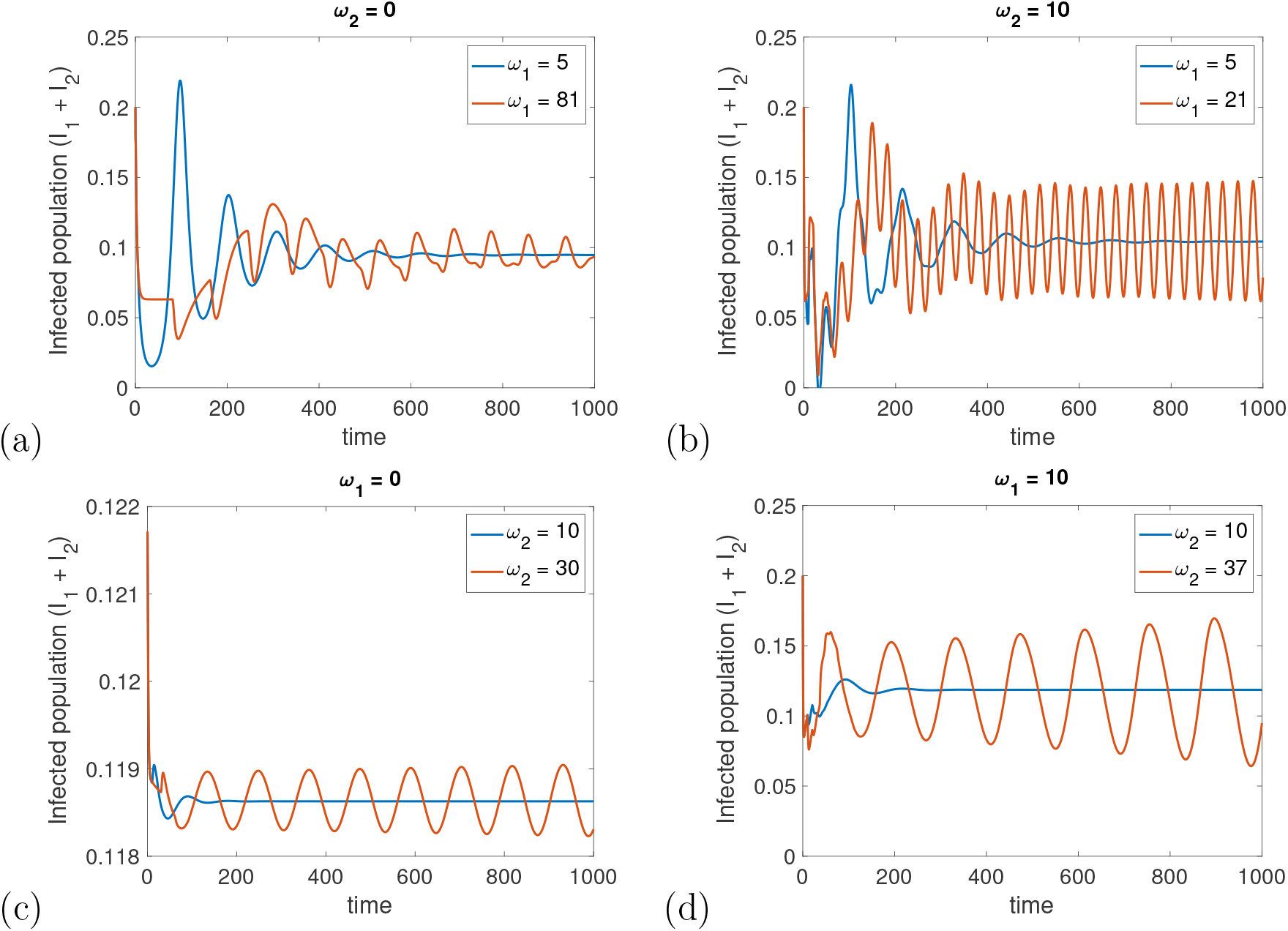
Long-term dynamics of the infectious population *I*_1_ + *I*_2_ under different combinations of time delays. Panels (a)-(b) show trajectories for varying *ω*_1_, with the relapse delay fixed at *ω*_2_ = 0 and *ω*_2_ = 10, using *ψ*_2_ = 4.25 and *ψ*_2_ = 0.43, respectively. Panels (c)-(d) show trajectories for varying *ω*_2_, with *ω*_1_ = 0 and *ω*_1_ = 10, using *α* = 0.5 and *ψ*_2_ = 0.15. In each panel, the remaining parameters take the illustrative values given in Eq. (6.1). The parameters *ψ*_2_ and *α* are adjusted between panels so that each delay combination lies near its corresponding stability boundary.

Furthermore, Figure 14 illustrates the Hopf bifurcation framework across different delay settings. In Figure 14(a) and (b), increasing the effective population-level infection-to-transmission delay *ω*_1_ past its critical threshold results in a transition from asymptotic stability to periodic oscillations when *ω*_2_ = 0 and when *ω*_2_ is maintained at a positive value, respectively. Similarly, Figure 14(c) and (d) demonstrate that increasing *ω*_2_ beyond its critical threshold destabilizes the endemic equilibrium when *ω*_1_ = 0 and *ω*_1_ = 10, respectively. These results collectively indicate that, for this parameter set, the effective population-level infection-to-transmission delay and post-recovery delay can each destabilize the endemic equilibrium, with their critical thresholds depending on the configuration of the other delay.

**Figure 14:**
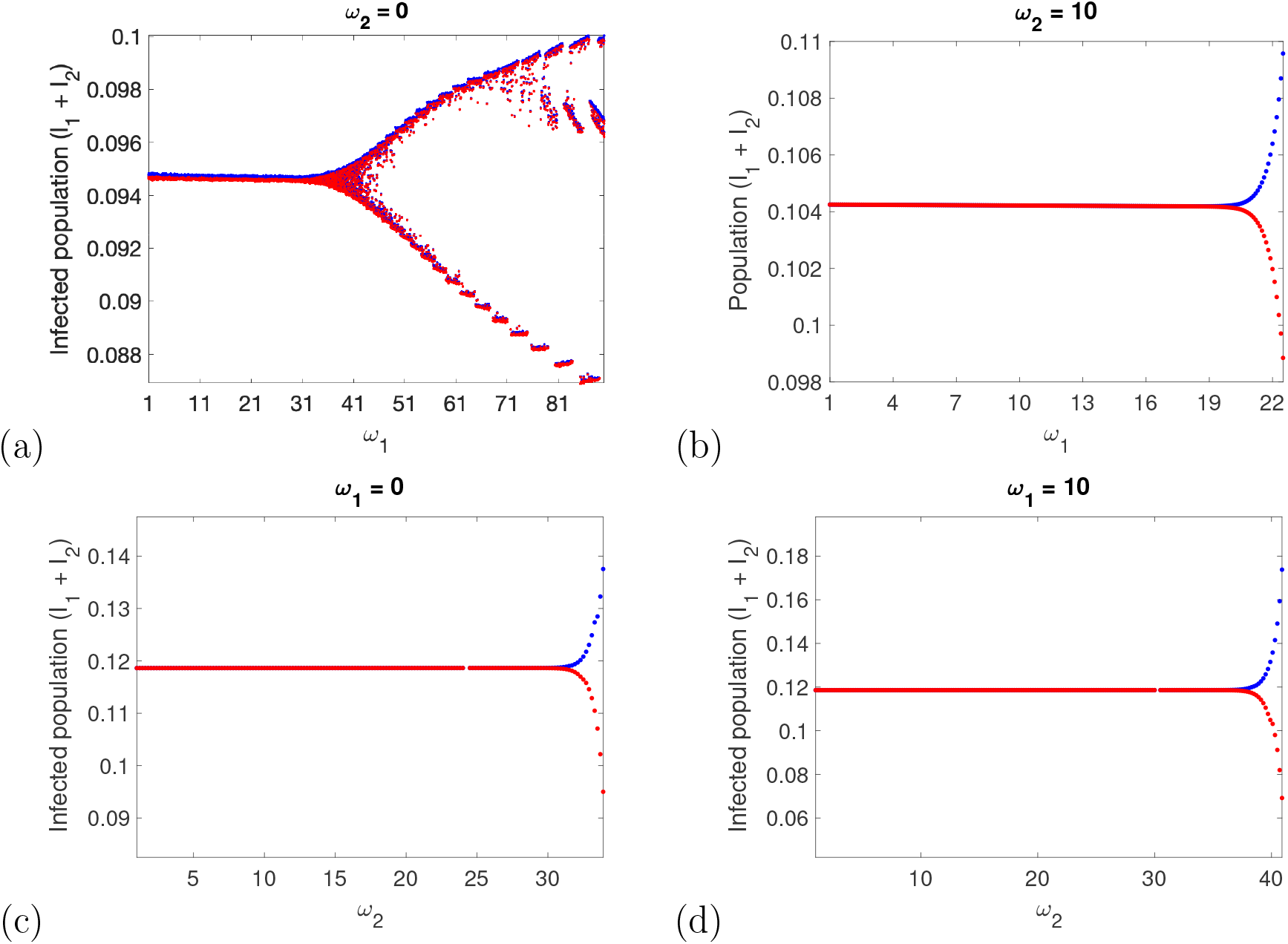
Hopf bifurcation diagrams of the infectious population (*I*_1_ + *I*_2_) with respect to the time-delay parameters. Panels (a)-(b) show bifurcation diagrams with respect to *ω*_1_, with the relapse delay fixed at *ω*_2_ = 0 and *ω*_2_ = 10, using *ψ*_2_ = 4.25 and *ψ*_2_ = 0.43, respectively. Panels (c)-(d) show bifurcation diagrams with respect to *ω*_2_, with *ω*_1_ = 0 and *ω*_1_ = 10, using *α* = 0.5 and *ψ*_2_ = 0.15. In each panel, the remaining parameters take the illustrative values given in Eq. (6.1). The blue and red curves give the upper and lower envelopes of the resulting oscillatory solutions

These oscillatory regimes arise only under hypothetical parameter combinations that exceed empirical estimates for Nipah, underscoring that the Hopf bifurcation serves primarily as a theoretical demonstration of the dynamical capacity of the DDE framework rather than a prediction of recurring epidemic cycles under current epidemiological conditions for NiV disease.

To verify the theoretical critical threshold, we consider the case in Sections 4.4 and 4.5, where *ω*_1_ is the bifurcation parameter and *ω*_2_ is fixed in its stable interval. For the case shown in Figure 13(b), the analytical sufficient bound obtained from Eq. (4.9) is

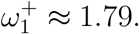

To compare this analytical bound with the exact stability boundary, we solved the characteristic equation at the endemic equilibrium *D*^*^. The leading pair of characteristic roots crosses the imaginary axis at the Hopf point 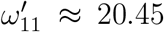, *p′* ≈ 0.190442. To confirm this threshold numerically, system (2.1) was simulated from a small perturbation of *D*^*^, using an initial history that satisfies conditions (2.3) and (2.4), and the exponential growth or decay rate of the oscillation amplitude of *I*_1_ + *I*_2_ was estimated after discarding transients. The numerically estimated rates agree with the real part of the leading characteristic root to three significant figures and change sign at *ω*_1_ ≈ 20.45, in agreement with 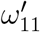 (Figure 14(b)). Condition (2.4) was verified along all trajectories shown. Hence,

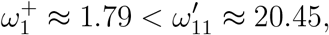

showing that the analytical bound lies well inside the stability region. The gap is expected: 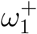 is obtained from sufficient bounding inequalities and certifies a guaranteed stability interval in closed form, whereas 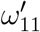 is the exact boundary obtained by solving the characteristic equation numerically. The two results are therefore complementary, the former providing an a priori guarantee and the latter the sharp threshold.

To further characterize the local Hopf bifurcation in Figure 13(b), we evaluated the normal-form coefficients derived in Section 4.4 for the representative case in which *ω*_2_ = 10 is fixed and *ω*_1_ is used as the bifurcation parameter. At the Hopf point 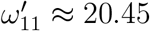, the corrected normal-form expressions give

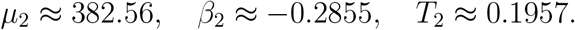

Therefore, *µ*_2_ *>* 0 indicates that the Hopf bifurcation is supercritical, whereas *β*_2_ *<* 0 indicates that the bifurcating small-amplitude periodic solutions near the Hopf point are stable. In addition, *T*_2_ *>* 0 implies that the period of these bifurcating periodic solutions increases locally as *ω*_1_ crosses the Hopf threshold. Hence, the normal-form calculation confirms that the characteristic Hopf point gives rise locally to a stable small-amplitude oscillatory branch. These properties are local and hold in a neighbourhood of 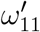 .

Figure 15 illustrates the transcritical bifurcation with respect to the human-to-human transmission threshold ℛ_*h*_, as obtained in Theorem 4.7. No biologically feasible positive endemic equilibrium exists when ℛ_*h*_ *<* 1, and near the threshold the disease-free equilibrium is locally asymptotically stable. When ℛ_*h*_ crosses unity, the disease-free equilibrium loses its stability and a positive endemic equilibrium emerges. More precisely, the endemic equilibrium branch satisfies 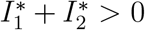 only for ℛ_*h*_ *>* 1, while 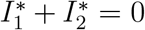 at ℛ_*h*_ = 1. Thus, the disease-free and endemic equilibrium branches exchange stability at ℛ_*h*_ = 1, confirming that model (2.1) undergoes a forward transcritical bifurcation at the disease-free equilibrium.

**Figure 15:**
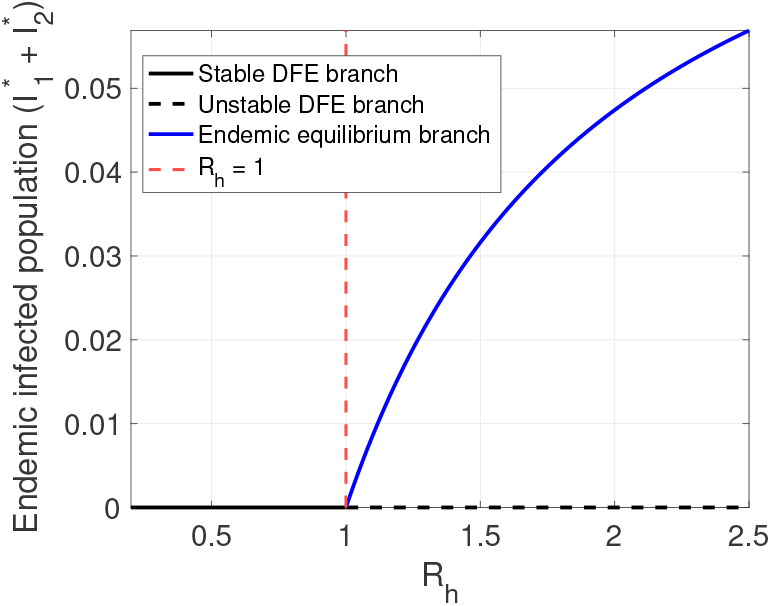
Transcritical bifurcation diagram with respect to the human-to-human transmission threshold ℛ_*h*_. The black solid curve represents the stable disease-free equilibrium for ℛ_*h*_ *<* 1, the black dashed curve the unstable disease-free equilibrium for ℛ_*h*_ *>* 1. The blue curve shows the positive endemic equilibrium branch, which appears when ℛ_*h*_ *>* 1. The red dashed vertical line marks the critical threshold ℛ_*h*_ = 1. All remaining parameter values are as specified in Eq. (6.1), with *ω*_1_ = *ω*_2_ = 10.

**Figure 16:**
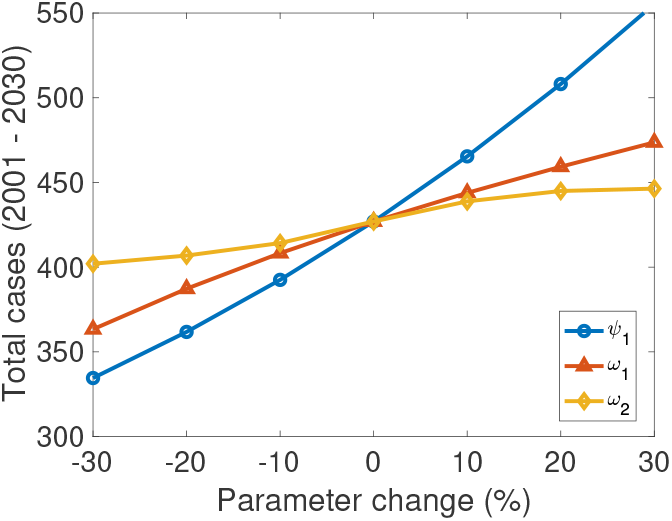
Total simulated NiV cases over 2001-2030 under parameter perturbations. The figure shows the cumulative number of cases generated by the model for varying values of the immediate relapse rate (*ψ*_1_), the effective population-level infection-to-transmission delay (*ω*_1_), and the delayed-relapse time (*ω*_2_), with all other parameters fixed at their baseline values.

**Figure 17:**
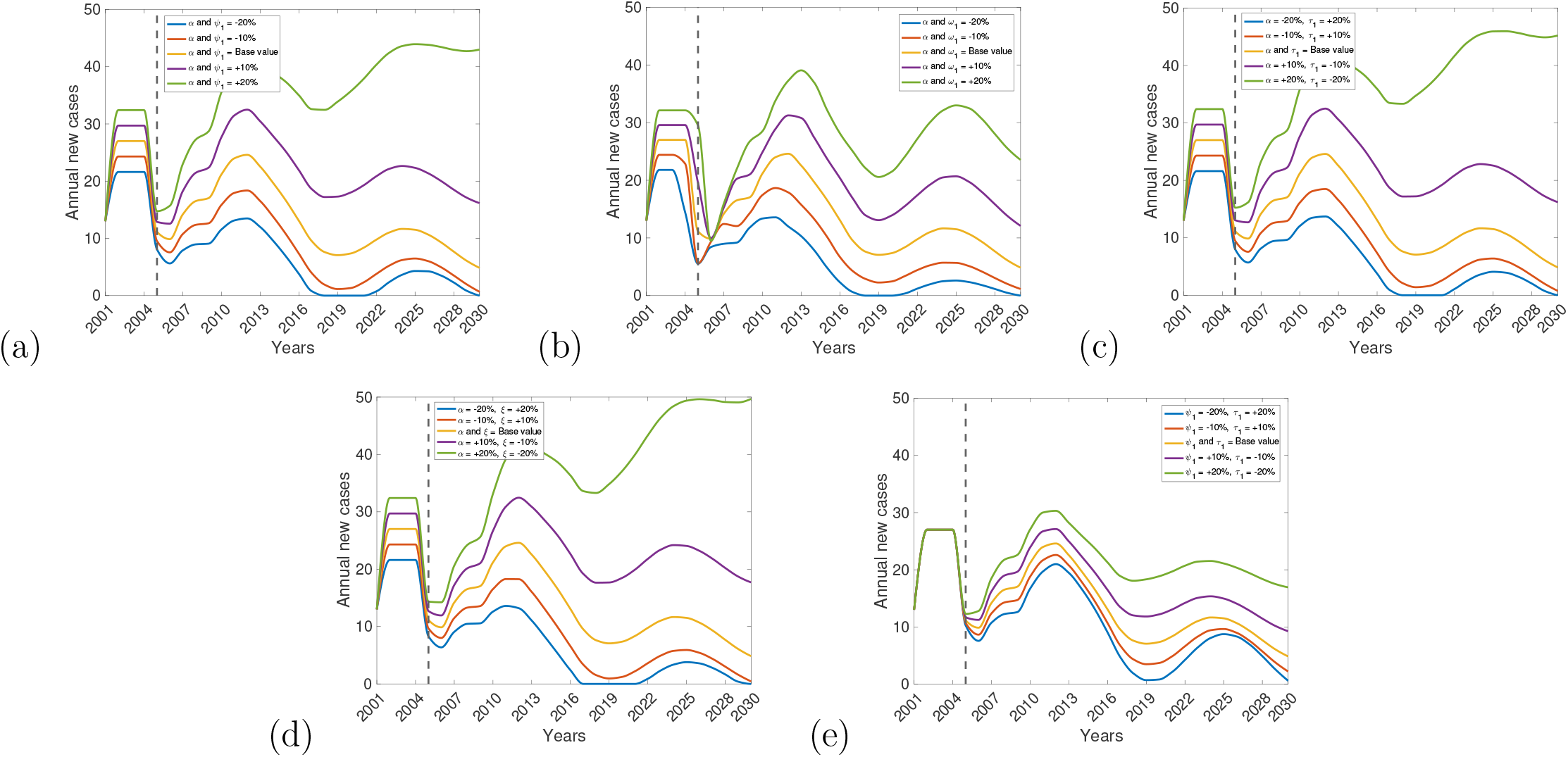
Time-series trajectories corresponding to the joint parameter perturbations underlying the sensitivity envelopes shown in Figure 9. Each panel displays model trajectories obtained by simultaneously varying two parameters within ±20% of their baseline values, while all other parameters are held fixed. Panels correspond to the following joint parameter combinations: (a) transmission rate (*α*) and immediate relapse rate (*ψ*_1_), (b) transmission rate (*α*) and effective population-level infection-to-transmission delay (*ω*_1_), (c) transmission rate (*α*) and treatment or clinical-management rate of early-stage infectious individuals (*τ*_1_), (d) transmission rate (*α*) and hospitalization rate of later-stage infectious individuals (*ξ*), and (e) immediate relapse rate (*ψ*_1_) and treatment or clinical-management rate of early-stage infectious individuals (*τ*_1_). Individual trajectories differ in magnitude and timing, with the largest separation occurring during the later phase of the outbreak. Together, these trajectories illustrate the range of simulated epidemic outcomes that define the envelope bounds presented in the main text.

## 7 Discussion

Emerging and re-emerging infectious diseases have consistently posed serious threats to both human and animal life. Disease control is frequently hampered not only by initial transmission but also by relapse, when persons who seem clinically recovered subsequently exhibit symptoms again and may contribute to future transmission. Moreover, intrinsic biological temporal delays, such as infection-to-transmission periods, the delayed onset of severe disease (relapse onset), or delayed therapeutic responses, can further complicate epidemic control by modifying the timing and dynamics of transmission. For NiV disease, a portion of recovered patients subsequently relapse or develop delayed-onset encephalitis [18,20], underscoring the necessity of explicitly considering relapse and temporal delays when evaluating long-term dynamics and formulating control strategies. Numerous mathematical models have analyzed NiV transmission in humans, predominantly concentrating on transmission mechanisms and intervention strategies, including isolation, vaccine, or therapy [26–31]. To our knowledge, however, no study has directly examined the synergistic effects of relapse and several time delays on NiV dynamics. This study addresses the gap by developing a unified DDE framework that rigorously characterizes how multiple delay mechanisms and relapse-mediated feedback interact to generate bifurcation phenomena, including transcritical bifurcation and delay-induced Hopf bifurcations.

Analytical findings for the proposed deterministic DDE model indicate that the disease-free equilibrium is locally asymptotically stable if the characteristic roots of the complete linearized delay system lie in the open left half-plane (see Section 4). We derive the human-to-human transmission threshold ℛ_*h*_ from the cumulative infectious residence time and establish its threshold property directly from the characteristic equation. We also establish a unique biologically feasible endemic equilibrium for the model and determine the corresponding algebraic feasibility conditions. The existence of a positive endemic equilibrium is governed by the condition ℛ_*h*_ *>* 1. Because relapse extends the expected time spent in the infectious classes, it enters ℛ_*h*_ directly and can therefore replenish the infectious compartments, increase the potential for onward transmission, and prolong disease persistence. However, relapse does not generate a biologically feasible endemic equilibrium when ℛ_*h*_ ≤ 1.

Our findings additionally indicate that time delays further influence system stability. Provided the associated transversality conditions hold, the endemic equilibrium is locally asymptotically stable while the delays remain below their critical thresholds. However, when delays surpass these values, stability is lost through Hopf bifurcation, resulting in persistent oscillations. Biologically, this corresponds to recurrent epidemic cycles rather than convergence to a stable endemic state, although in the present model such oscillations arise only under the illustrative parameter regimes explored in Subsection 6.4. We further characterize the direction and stability of these oscillations and estimate critical delay lengths using the Nyquist criterion, providing quantitative thresholds at which oscillatory dynamics emerge.

The transcritical bifurcation analysis further confirms the threshold nature of human-to-human NiV transmission in the proposed model. Near ℛ_*h*_ = 1, the disease-free and endemic equilibria exchange stability: for ℛ_*h*_ slightly below unity, *D*_0_ is locally stable and no biologically feasible endemic equilibrium exists, whereas for ℛ_*h*_ slightly above unity, *D*_0_ loses stability and a positive endemic equilibrium emerges. This local exchange of stability indicates that even small increases in transmission intensity, infectious residence time, or relapse-related feedback can shift the system from disease elimination toward endemic persistence near the threshold. Epidemiologically, this result highlights the importance of maintaining ℛ_*h*_ below unity through early detection, reduction of effective contacts, timely supportive care, and management of relapse or delayed encephalitic outcomes.

Using reported NiV cases from Bangladesh between 2001 and 2024, we calibrated the model and estimated key epidemiological quantities, including the human-to-human transmission rate *α*, disease-induced death rate *δ*, relapse rates *ψ*_1_ and *ψ*_2_, and time delays *ω*_1_ and *ω*_2_. We further generated forward projections through 2030 to explore potential epidemic trajectories under the calibrated baseline assumptions. The calibrated trajectory reproduces a secondary rise peaking around 2023-2024, and the projections show a subsequent decline through 2030. Within the model, this recurrence arises from the interplay of the relapse pathway and the delays; however, because the model omits repeated zoonotic introductions, these mechanisms may partly absorb recurrence that is driven by spillover in reality. The model-derived relapse contribution should therefore be interpreted as an effective replenishment mechanism rather than as evidence that relapse drives recurrent NiV outbreaks.

The practical identifiability analysis (Section 5.3) further clarifies which conclusions the annual incidence data support. The transmission rate *α*, the disease-induced death rate *δ*, the delays *ω*_1_ and *ω*_2_, and the initial infectious population *I*(0) are practically identifiable, whereas the relapse rates *ψ*_1_ and *ψ*_2_ are not: the data provide only a lower bound for *ψ*_1_ and an upper bound for *ψ*_2_. Consequently, the individual relapse rates, and quantities derived from them such as the relapse contribution fraction, should not be interpreted as epidemiological estimates. This non-identifiability does not change the conclusion that human-to-human transmission is subcritical: ℛ_*h*_ remains between 0.67 and 0.95 along all profile likelihoods, and 96% of bootstrap replicates yield ℛ_*h*_ *<* 1.

Reductions in the immediate relapse rate *ψ*_1_ suppress the late-period rise in incidence and reduce cumulative incidence, whereas increases in *ψ*_1_ amplify this late rise and increase cumulative incidence. Given the non-identifiability of *ψ*_1_, these scenarios describe the model-based role of relapse-driven replenishment rather than clinically estimated relapse effects. Changes in the effective population-level infection-to-transmission delay *ω*_1_ alter the entire simulated trajectory, with reductions in *ω*_1_ producing substantial decreases in cumulative incidence under the parameter ranges considered. Since *ω*_1_ enters ℛ_*h*_ only through the survival factor 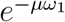, this effect operates mainly through the early dynamics, via the delayed-history period and the initial exposed population determined by (2.3), rather than through a change in the threshold itself. By comparison, the delayed relapse time *ω*_2_ has a smaller effect on cumulative incidence but shifts the timing of the late rise, reflecting the delayed return of recovered individuals to the infectious compartment *I*_2_. Collectively, these findings indicate that the infection-to-transmission delay primarily shapes the early outbreak phase and overall epidemic magnitude, whereas relapse-associated mechanisms, particularly the immediate relapse pathway, become increasingly influential during the post-peak phase.

These sensitivity patterns also have implications for surveillance and control. Measures that reduce effective transmission through early detection, isolation, and timely clinical management may suppress both the initial outbreak and the later recurrent rise in incidence. Although the time delays *ω*_1_ and *ω*_2_ may not be directly modifiable, their influence on epidemic timing highlights the need for timely diagnosis and extended follow-up. Interventions capable of reducing relapse frequency, were they available, may further lower post-peak and cumulative disease burden, underscoring the value of continued post-recovery monitoring. Improved empirical estimates of relapse frequency and timing would reduce uncertainty in long-term projections and strengthen surveillance planning.

Joint scenario analyses further show that the effects of relapse and time delays depend strongly on their interactions with transmission and clinical-management parameters. Concurrent variation in the transmission rate *α* with either the immediate relapse rate *ψ*_1_ or the effective population-level infectionto-transmission delay *ω*_1_ affects both the initial epidemic phase and the later recurrent rise, indicating that these mechanisms jointly shape the simulated outbreak patterns. Scenarios combining reduced transmission with increased early-stage clinical management *τ*_1_ or hospitalization *ξ* attenuate both the early and later humps, highlighting the potential benefit of coordinated interventions that reduce transmission while improving the clinical response. In contrast, joint variation in *ψ*_1_ and *τ*_1_ mainly affects the later phase, consistent with the interpretation that relapse-related mechanisms become more influential after a recovered population has accumulated. Overall, the joint perturbation results indicate that the later phase of the epidemic is sensitive to combined changes in relapse, delays, transmission, and clinical-management parameters.

The estimated human-to-human transmission threshold, ℛ_*h*_≈0.83, lies below unity at the calibrated point estimate, although parameter uncertainty admits values on both sides of the threshold. Taken at face value, this indicates that sustained NiV transmission is unlikely in the absence of additional zoonotic introductions, which is compatible with the observed epidemiology in Bangladesh, where a substantial proportion of cases arise through person-to-person transmission but the resulting chains are not self-sustaining [15]. The sporadic occurrence of reported outbreaks is also strongly influenced by repeated spillover events [15, 16], which are not represented explicitly in the present model. The sensitivity analysis further indicates that relapse-related processes influence ℛ_*h*_ and the timing of later incidence peaks. Consequently, strategies focused exclusively on reducing contact-based transmission may not fully address delayed post-recovery disease dynamics, whether these contribute to transmission or constitute clinical burden alone. Long-term management may therefore benefit from continued follow-up of recovered patients, timely identification of relapse or delayed-onset disease, and sustained surveillance for both secondary transmission and renewed zoonotic introductions.

The no-delay simulations further clarify the distinct roles of relapse and time-delay mechanisms in the model. Increasing *ψ*_1_ raises the endemic infection level by returning recovered individuals to the early infectious class, but it does not generate sustained recurrent oscillations in the absence of explicit delays (Figure 12). This shows that relapse strengthens disease persistence and increases the long-term burden without, by itself, producing oscillatory dynamics. The model exhibits sensitivity to the length of the delays *ω*_1_ and *ω*_2_, with several scenarios demonstrating that stability of the endemic equilibrium is lost once the delays surpass their critical thresholds. Numerical simulations, encompassing time-series plots and bifurcation diagrams, consistently support the theoretical findings. From a biological standpoint, these results indicate that the effective population-level infection-to-transmission period and the onset delay of encephalitis both influence epidemic trajectories, with *ω*_1_ having the stronger effect. Within the illustrative regimes considered, short delays permit convergence to a stable endemic state, whereas longer delays increase the likelihood of sustained oscillations.

Our study has several limitations. The model does not explicitly incorporate zoonotic or foodborne spillover, which are central to the initiation of NiV outbreaks. Therefore, it is not a complete explanation of the sporadic annual incidence pattern observed in Bangladesh. Instead, the model isolates the contributions of human-to-human transmission, relapse, and time delays after infection enters the human population. Consequently, it cannot reproduce abrupt reintroductions, multi-year gaps, or environmentally driven seasonal exposure. Future extensions could incorporate a seasonal or stochastic spillover force representing repeated animal-to-human or foodborne introductions. Furthermore, the delayed relapse term used here represents a phenomenological delayed-state feedback rather than a strict cohort-based fixed-duration transition. Accordingly, the delayed relapse outflow is linked to the recovered population at an earlier time rather than explicitly to the cohort entering the recovered compartment. A more mechanistic formulation could instead derive the relapse outflow from the cohort entering *R* at time *t*− *ω*_2_, together with the appropriate survival over the intervening interval. Such cohort-structured or age-since-recovery formulations provide an important direction for future work when longitudinal post-recovery data become available.

Moreover, the relapse rates *ψ*_1_ and *ψ*_2_ are not practically identifiable from annual incidence data (Section 5.3), and the effective delay *ω*_1_ ≈ 3.2 years reflects population-level infection-to-transmission timing. We emphasize that *ω*_1_ is itself practically identifiable, with 95% profile confidence interval (2.638, 4.153) years and negligible loading on the weak Fisher-information direction (Section 5.3). The multi-year value is therefore required by the annual incidence series rather than an artefact of weak parameter constraint, which supports its interpretation as an effective population-level lag encoding the observed spacing between outbreaks. Monthly or case-level surveillance data, together with post-recovery follow-up data, would help constrain these quantities. The model further assumes homogeneous mixing, deterministic dynamics, and fixed delays, and does not include spatial structure, behavioral responses, or distributed delays. Finally, sustained oscillations arise only under hypothetical parameter regimes outside currently available NiV estimates. These oscillations are illustrations of the model’s theoretical dynamical potential rather than forecasts of future epidemic behavior.

Future methodological extensions may also benefit from integrating mechanistic epidemic models with complementary data-driven approaches as richer NiV surveillance data become available. Recent studies have demonstrated such approaches for Ebola transmission using data from North Kivu, Democratic Republic of the Congo [55], and for SARS-CoV-2 dynamics using United Kingdom data [56], combining mechanistic modeling with data-informed inference, sensitivity analysis, and forecasting. Similar approaches could be explored for NiV when sufficiently detailed temporal and epidemiological data become available.

Despite these limitations, the analysis establishes that relapse and time delays can sustain recurrent incidence within a subcritical human-to-human system, a behaviour that models without post-recovery feedback cannot produce. Post-recovery viral persistence and survivor-associated resurgence have been documented for other emerging zoonotic pathogens, notably Ebola virus [57, 58], reinforcing the broader relevance of incorporating relapse-driven mechanisms into transmission models. At the same time, the calibrated model cannot separate relapse- and delay-mediated recurrence from recurrence driven by repeated zoonotic introduction; distinguishing the two requires both explicit spillover structure and finer-resolution surveillance data. A further general implication is that in delay models with post-recovery relapse, aggregated annual incidence may determine the transmission threshold while leaving the relapse rates unidentified, so the two classes of quantity warrant separate treatment when such models are reported.

## Data Availability

All data produced in the present work are contained in the manuscript.

## Data availability statement

The dataset supporting the findings of this study is included in this article.

## Acknowledgments

The research work of Sarita Bugalia and Liliana Salvador was supported by the National Science Foundation (Award # 2200269 “PIPP Phase I: BEHIVE - Behavioral Interaction and Viral Evolution for Pandemic Prevention and Prediction”). The research of Hao Wang was partially supported by the Natural Sciences and Engineering Research Council of Canada (Individual Discovery Grant RGPIN-2025-05734 and Discovery Accelerator Supplement Award PGPAS-2020-00090) and a Canada Research Chair.

## Appendix A

**Proof of Theorem 2.1:** We first establish local existence and uniqueness, then non-negativity, and finally boundedness and global continuation. Let

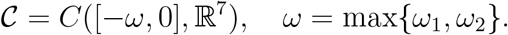

The right-hand side of system (2.1) is continuous in the history variable and locally Lipschitz on *C*, since it consists of linear terms and bilinear products of present and delayed state variables. Therefore, for every continuous initial history satisfying the compatibility condition (2.3), there exists a unique maximal solution

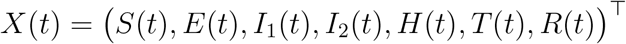

defined on an interval [0, *t*_max_), by the standard theory of retarded functional differential equations [43].

We next establish non-negativity of the state variables under the non-negative initial histories and the admissibility condition (2.4). At the boundary *S*(*t*) = 0, the first equation of system (2.1) gives

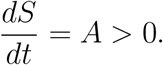

Hence *S*(*t*) cannot cross from the non-negative region into negative values.

For the remaining variables, suppose that *t*_1_ *>* 0 is the first time at which one of *E, I*_1_, *I*_2_, *H, T, R* reaches the boundary of the non-negative orthant, while all state variables remain non-negative on [0, *t*_1_).

Consider first *I*_1_(*t*_1_) = 0. From system (2.1),

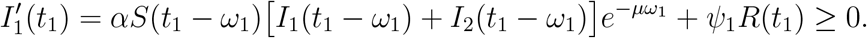

Thus *I*_1_(*t*) cannot cross below zero at *t*_1_. Similarly, if *I*_2_(*t*_1_) = 0, then

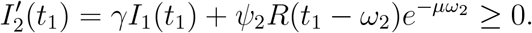

If *H*(*t*_1_) = 0, then

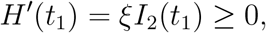

and if *T* (*t*_1_) = 0, then

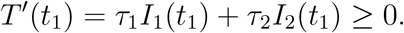

Hence *I*_1_, *I*_2_, *H*, and *T* remain non-negative.

For the exposed compartment, the compatibility condition (2.3) yields the representation

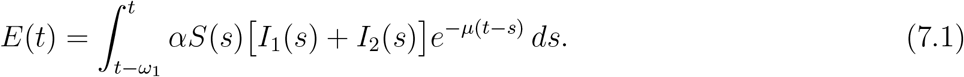

Since *S*(*s*), *I*_1_(*s*), and *I*_2_(*s*) are non-negative for all relevant times, the integrand is non-negative. Therefore,

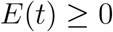

for all *t* ≥ 0.

Finally, consider the recovered compartment. If *R*(*t*_1_) = 0, then from the *R*-equation in system (2.1),

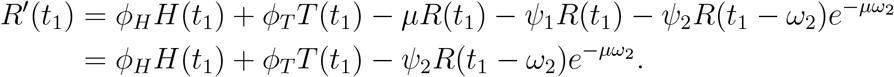

By condition (2.4),

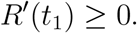

Thus *R*(*t*) cannot cross below zero. Therefore all state variables remain non-negative on their interval of existence.

We next establish boundedness. Define the total human population by

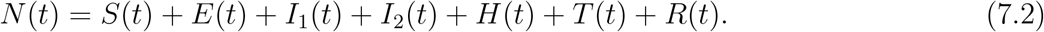

Adding the equations of system (2.1) gives

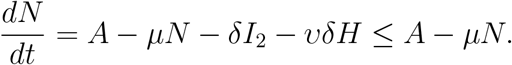

By the comparison principle,

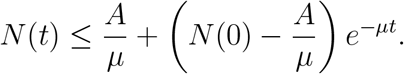

Consequently,

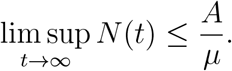

If

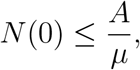

then

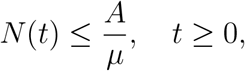

and hence the region

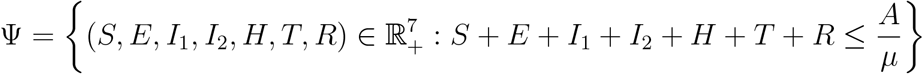

is positively invariant.

If *N* (0) *> A/µ*, then the preceding estimate shows that the solution remains bounded and satisfies

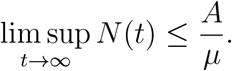

Thus every non-negative solution is bounded, while solutions whose initial data lie in Ψ remain in Ψ for all *t*≥0.

Since all state variables remain non-negative and *N* (*t*) is bounded on every finite interval, the solution remains in a bounded subset of the phase space. Therefore, by the continuation theorem for retarded functional differential equations, the maximal existence time satisfies

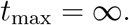

Hence system (2.1) admits a unique global non-negative solution under the stated assumptions, the region Ψ is positively invariant, and all non-negative solutions are bounded.□

## Appendix B

**Proof of Theorem 4.3:** To analyze the local asymptotic stability of *D*^*^, the characteristic equation of the linearized system at *D*^*^ is given by

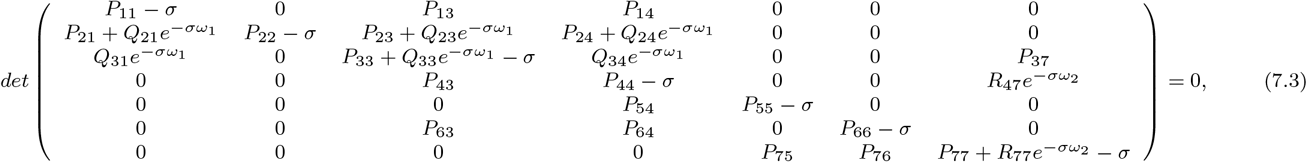

where

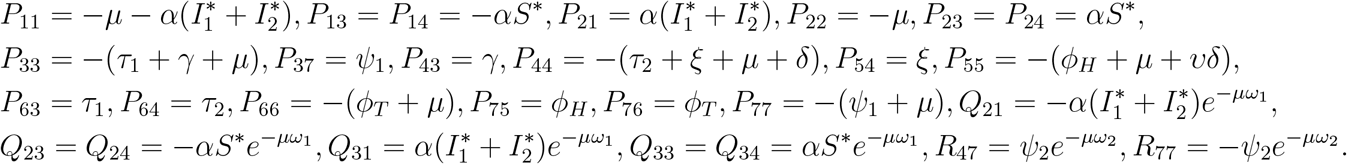

We can write the Eq. (7.3) as

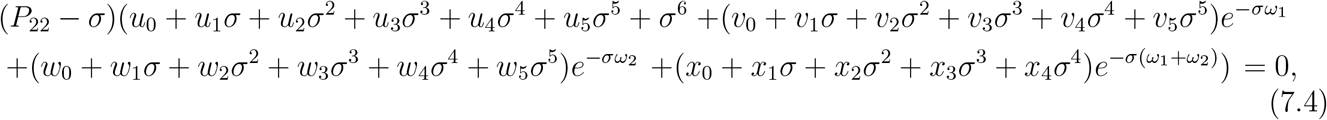

where *u*_*j*_, *v*_*j*_, *w*_*j*_, *j* = 0, 1, 2, 3, 4, 5, and *x*_*j*_, *j* = 0, 1, 2, 3, 4, are as follows:

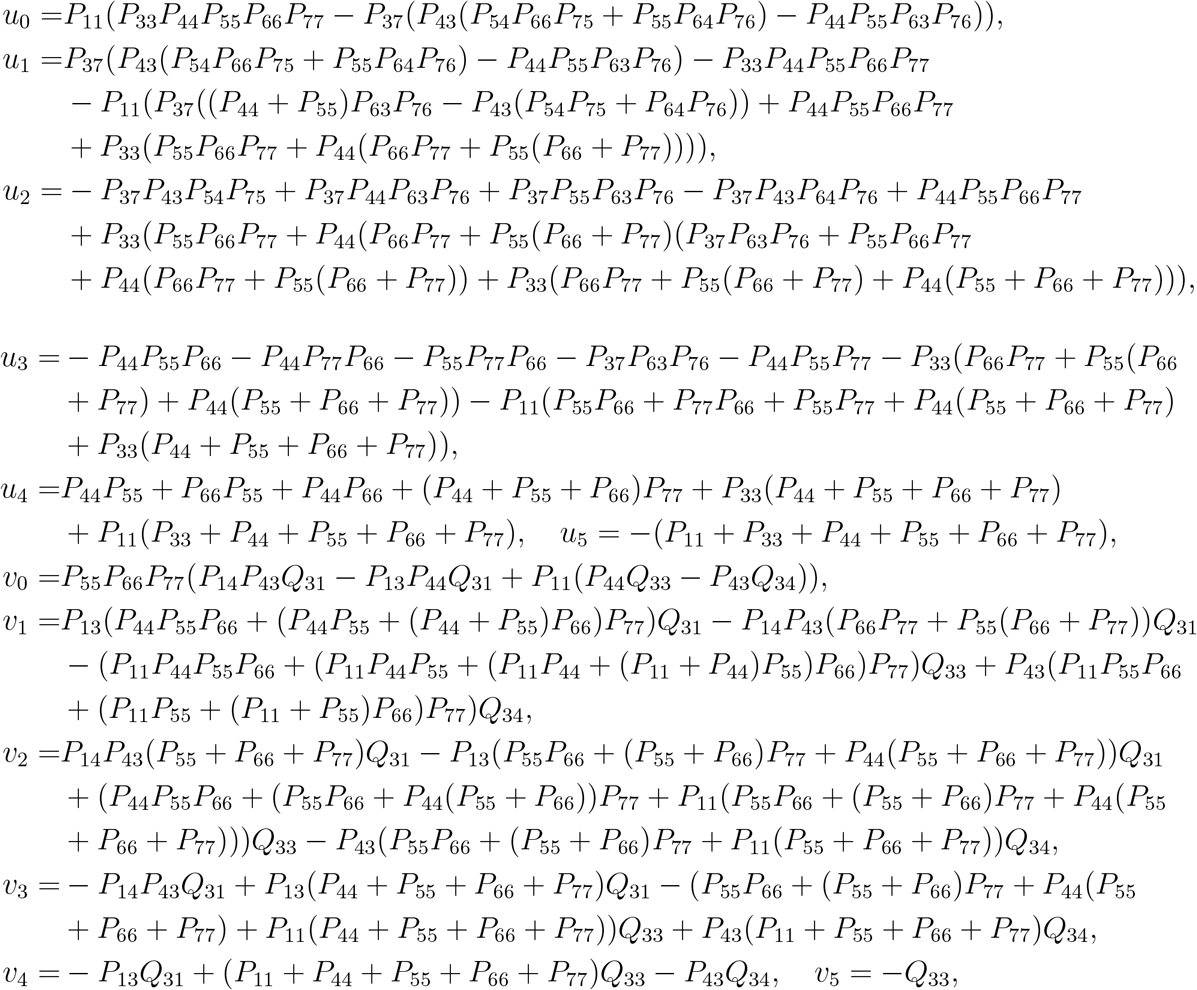

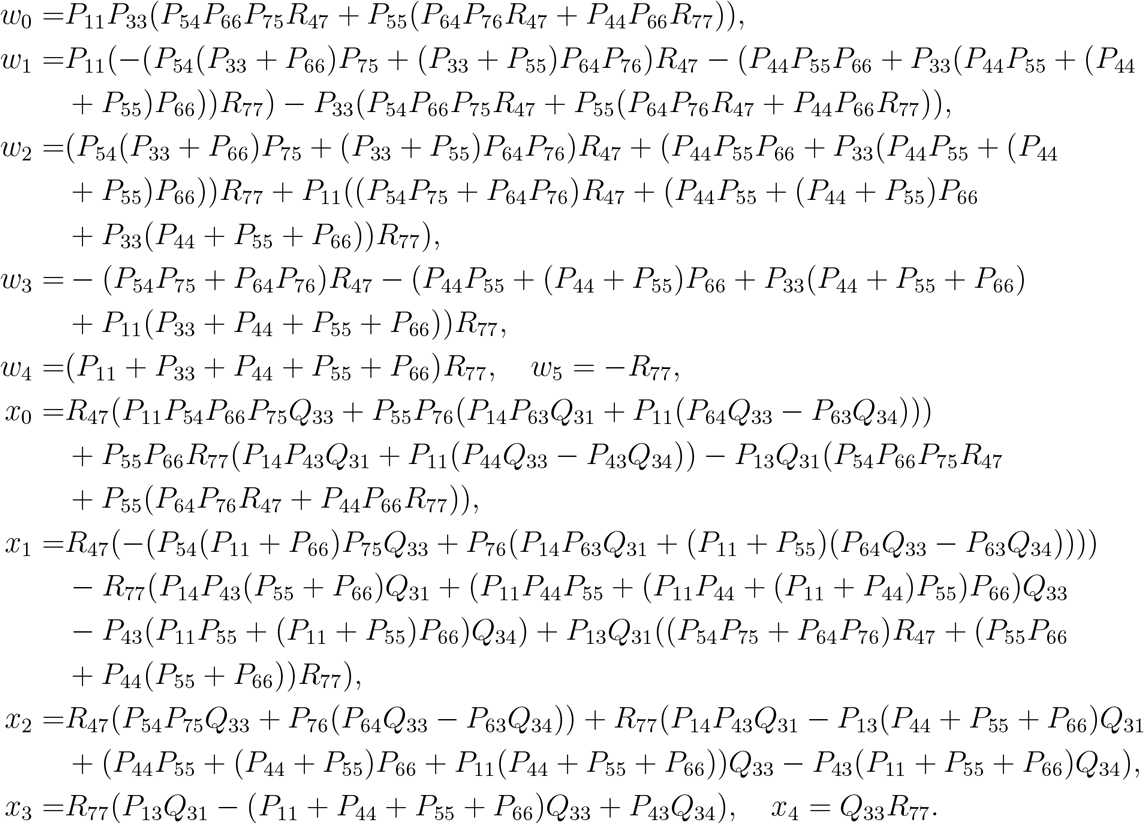

We first discuss the case when *ω*_1_ = *ω*_2_ = 0. Since *P*_22_ = −*µ <* 0, the root generated by the factor *P*_22_ −*σ* in Eq. (7.4) is always negative. Therefore, the stability of *D*^*^ is determined by the remaining sixth-degree polynomial. In this case, the characteristic Eq. (7.4) becomes

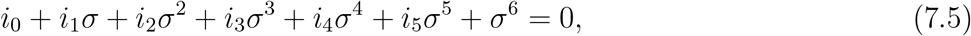

where

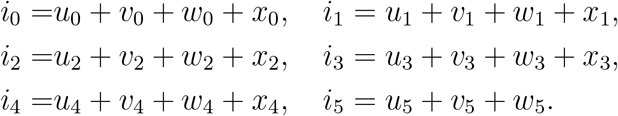

By the Routh-Hurwitz criterion, all roots of Eq. (7.5) have negative real parts if and only if

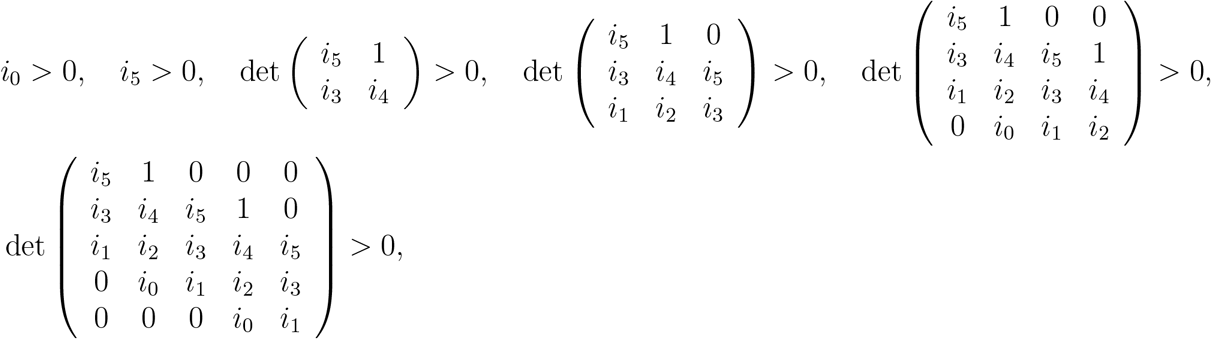

Thus, under these Routh-Hurwitz conditions, all roots of the characteristic equation have negative real parts. Hence, *D*^*^ is locally asymptotically stable when *ω*_1_ = *ω*_2_ = 0.

**Proof of Theorem 4.4:** When *ω*_1_ *>* 0, and *ω*_2_ = 0, then the characteristic Eq. (7.4) becomes

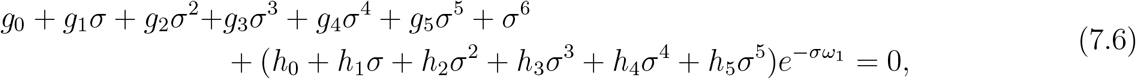

where

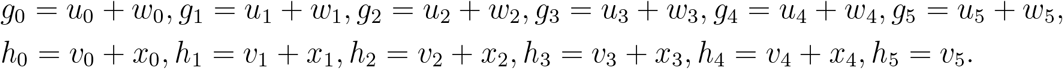

Let *σ* = *ip*_3_ (*p*_3_ *>* 0) be a root of Eq. (7.6). We have

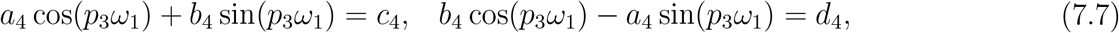

where

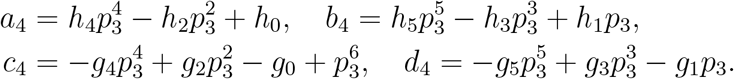

From Eq. (7.7), we have

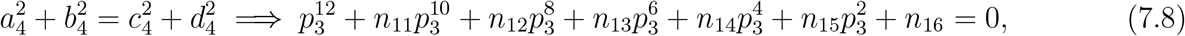

Where

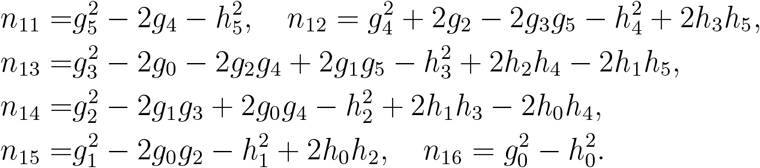

Let 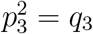, then Eq. (7.8) becomes in the following form:

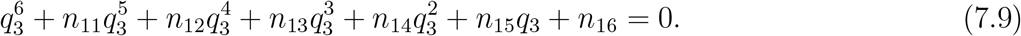

Assume that **H1:** Eq. (7.9) has a positive root *q*_30_, then Eq. (7.8) also has a positive root 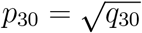. Furthermore, from Eq. (7.7), we obtain

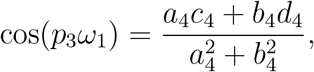

And

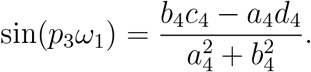

Replacing *p*_3_ by *p*_30_, the corresponding family of critical values of *ω*_1_ is given by

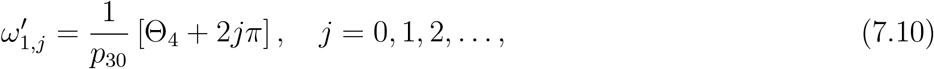

Where

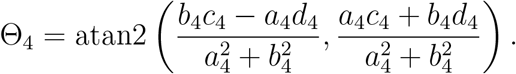

If Θ_4_ *<* 0, it is replaced by Θ_4_ + 2*π*. The first positive critical value is denoted by

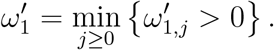

Now, differentiating Eq. (7.6) implicitly with respect to *ω*_1_, and then evaluating at *σ* = *ip*_30_ and 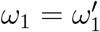, we obtain

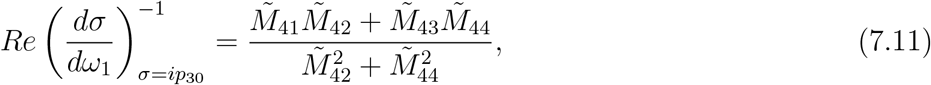

Where

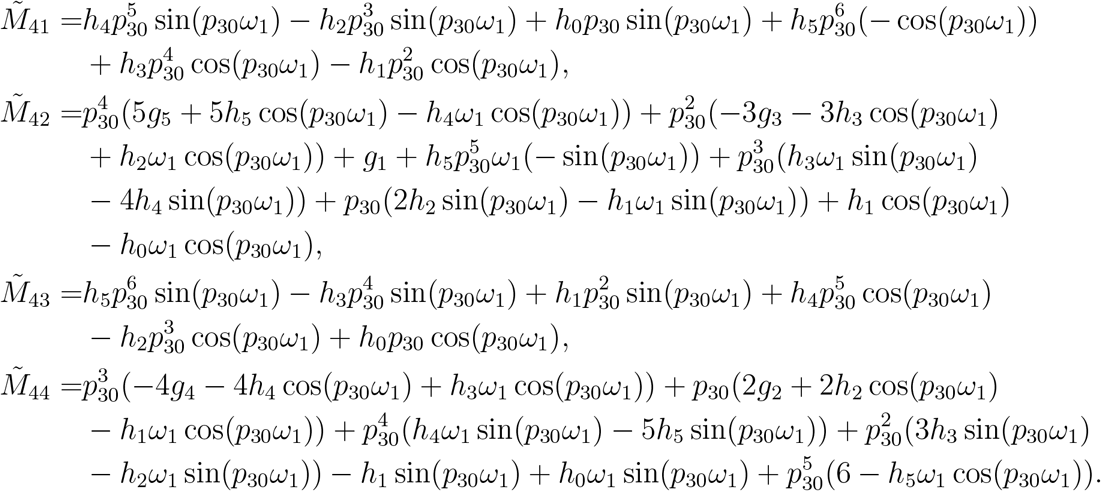

Hence, 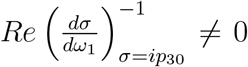 if the condition **H2:** 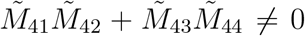 holds. Consequently, the Hopf bifurcation theorem [45] implies the Hopf bifurcation in system (2.1).

**Proof of Theorem 4.5:** When *ω*_1_ = 0, and *ω*_2_ *>* 0, then the characteristic Eq. (7.4) becomes

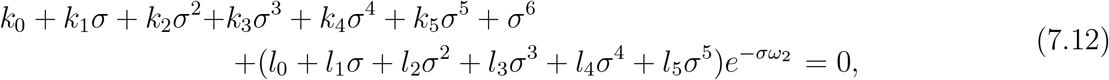

where

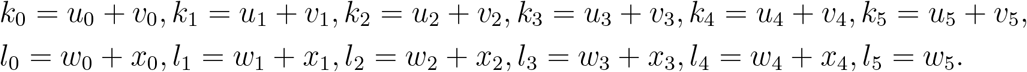

Let *σ* = *ip*_4_ (*p*_4_ *>* 0) be a root of Eq. (7.12). We have

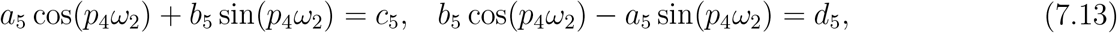

Where

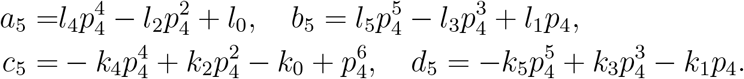

From Eq. (7.13), we obtain

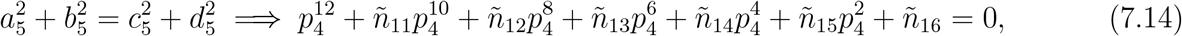

where

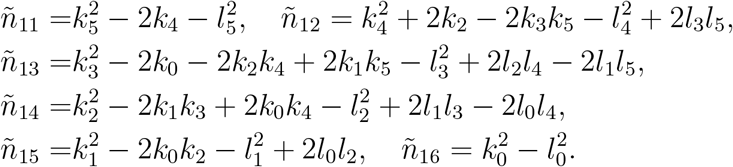

Let 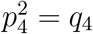. Eq. (7.14) becomes in the following form:

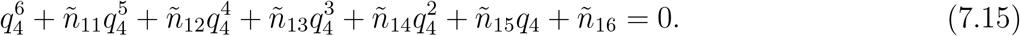

Assume that **H3:** Eq. (7.15) has a positive root *q*_40_, then Eq. (7.14) also has a positive root 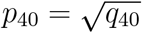. Furthermore, from Eq. (7.13), we obtain

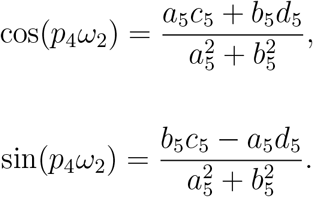

and

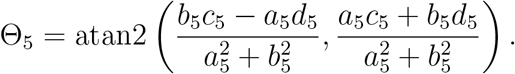

Replacing *p*_4_ by *p*_40_, the corresponding family of critical values of *ω*_2_ is given by

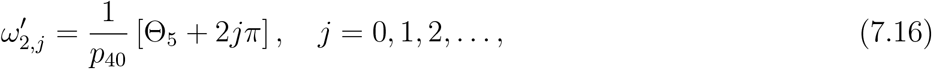

where

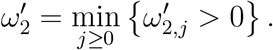

If Θ_5_ *<* 0, it is replaced by Θ_5_ + 2*π*. The first positive critical value is denoted by

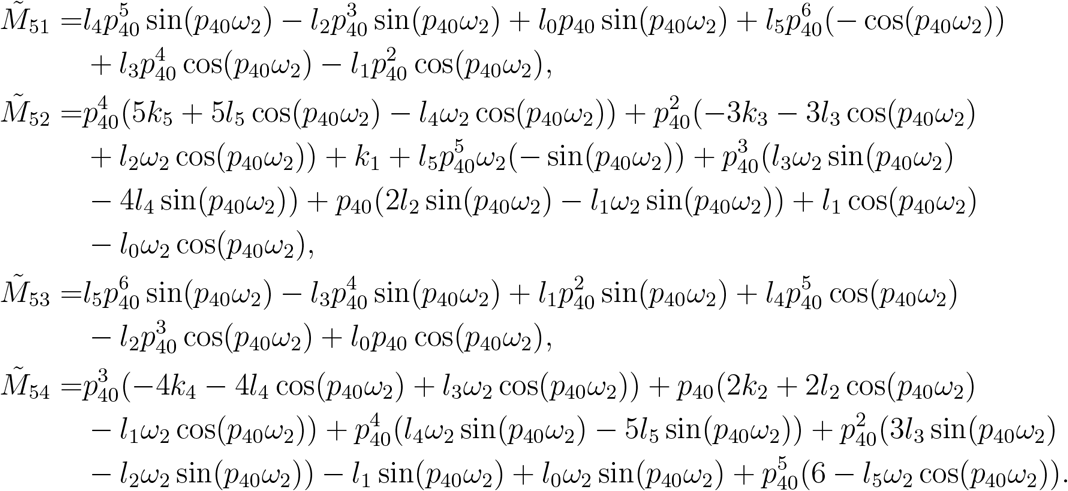

Now, differentiating Eq. (7.12) implicitly with respect to *ω*_2_, and then evaluating at *σ* = *ip*_40_ and 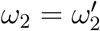, we obtain

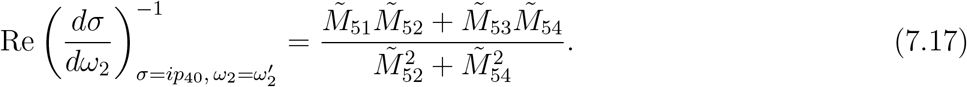

where

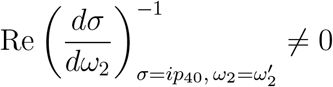

Hence,

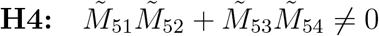

if the condition

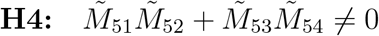

holds. Therefore, the transversality condition is satisfied. Consequently, by the Hopf bifurcation theorem [45], system (2.1) undergoes a Hopf bifurcation at *D*^*^ when

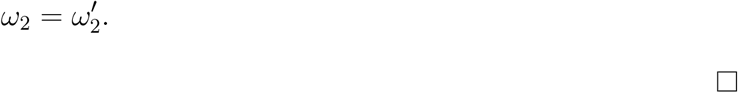

**Proof of Theorem 4.6:** When *ω*_1_ *>* 0 and 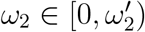. In this case, we take *ω*_1_ as bifurcation parameter and *ω*_2_ in stable interval. We can write Eq. (7.4) in the following form:

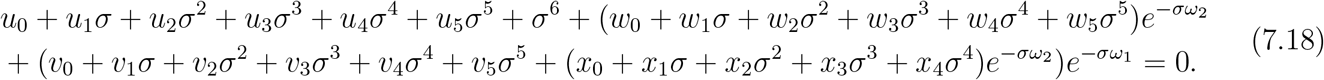

Let *σ* = *ip*_5_ (*p*_5_ *>* 0) be a root of Eq. (7.18). We have

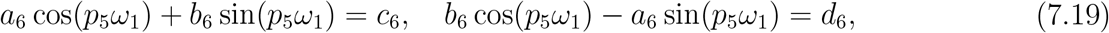

Where

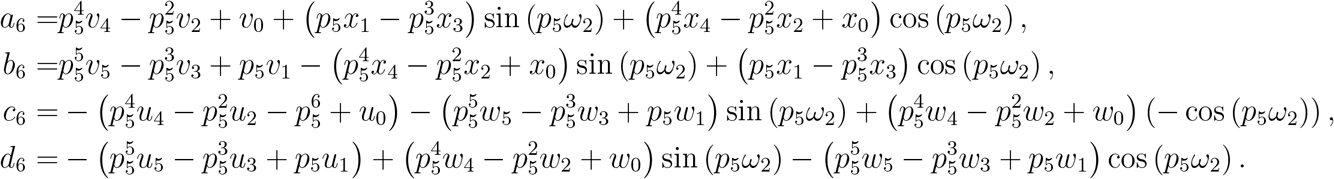

From Eq. (7.19), we have

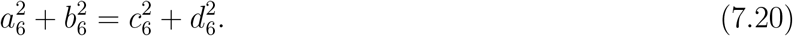

Assume that **H5:** Eq. (7.20) has a positive root *p*_50_. Furthermore, from Eq. (7.19), we obtain

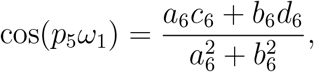

and

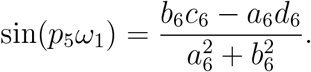

Replacing *p*_5_ by *p*_50_, the corresponding family of critical values of *ω*_1_ is given by

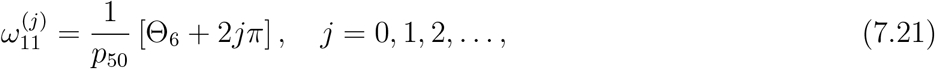

Where

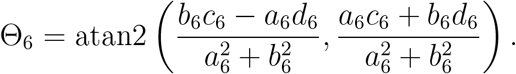

If Θ_6_ *<* 0, it is replaced by Θ_6_ + 2*π*. The first positive critical value is denoted by

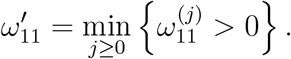

Further, differentiating Eq. (7.18) implicitly with respect to *ω*_1_, and then evaluating at *σ* = *ip*_50_ and 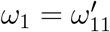, we obtain

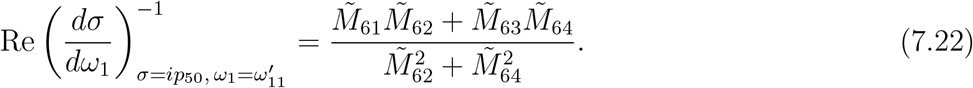

Where

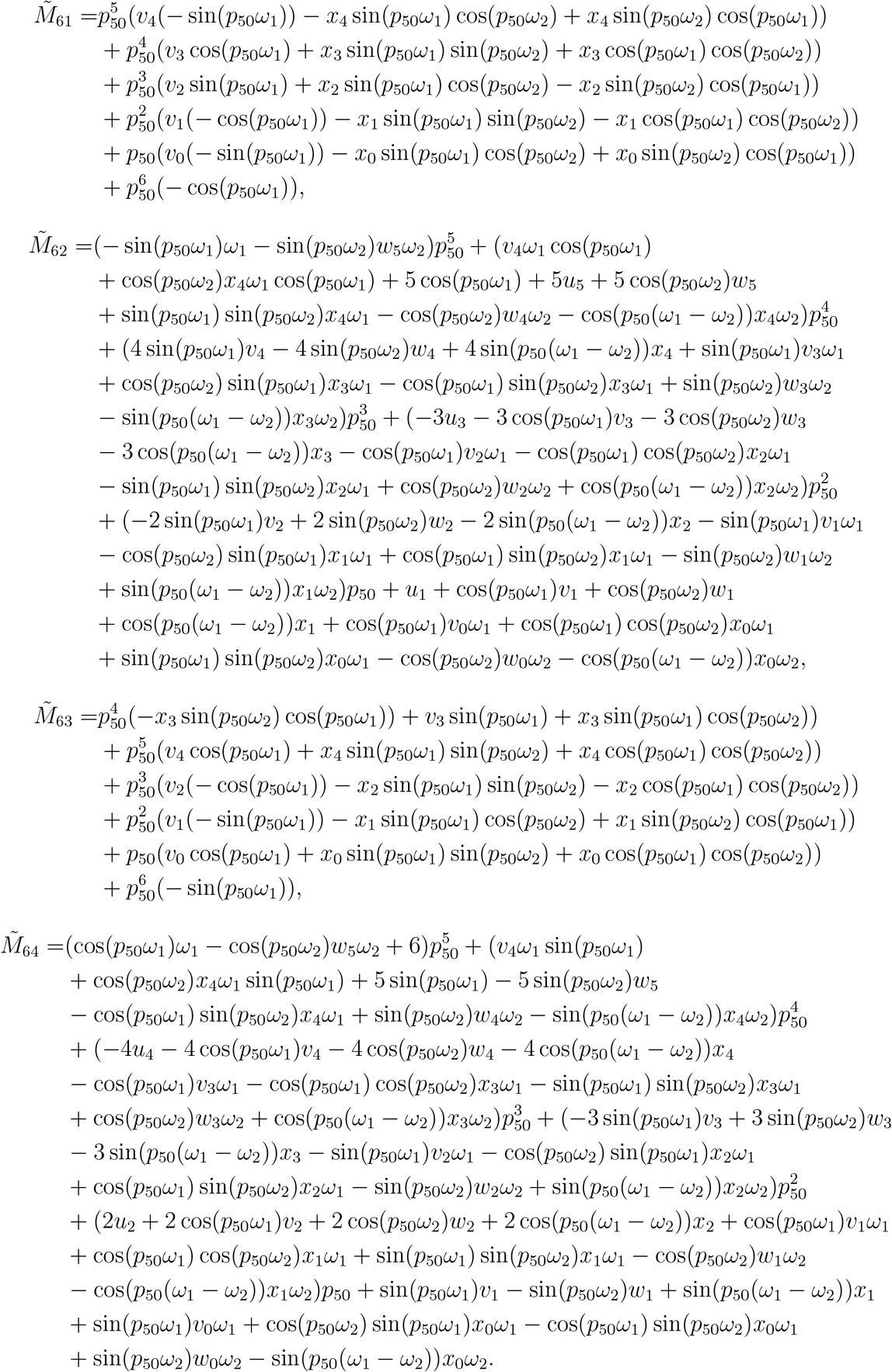

Hence,

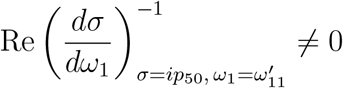

if the condition

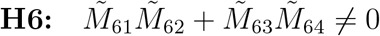

holds. Therefore, the transversality condition is satisfied. Consequently, by the Hopf bifurcation theorem [45], system (2.1) undergoes a Hopf bifurcation at *D*^*^ when

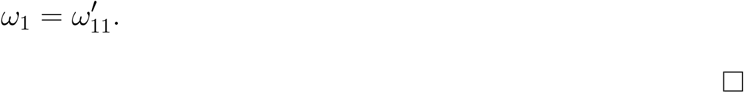

**Properties of Hopf bifurcation**

Here, we present the detailed derivation of the corresponding linear operator formulation, adjoint system, bilinear inner form, eigenvectors, center manifold reduction, and normal form coefficients used in the main text for the analysis of the properties of Hopf bifurcation at the threshold value of the time delay 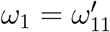 .

Assume

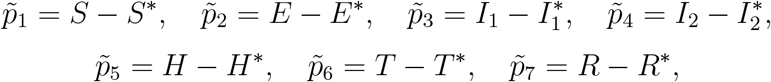

and

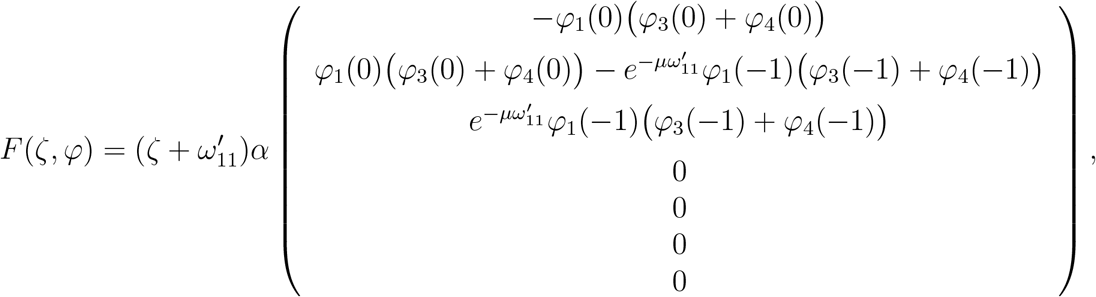

Further, normalize the time delay *ω*_1_ by the scaling of time *t*→ *t/ω*_1_. Thus, system (2.1) attains the functional differential form

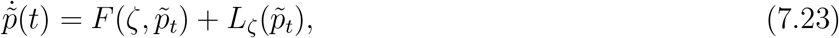

where *F* : ℝ × *C* → ℝ^7^ and *L*_*ζ*_ : *C* → ℝ^7^ are expressed as

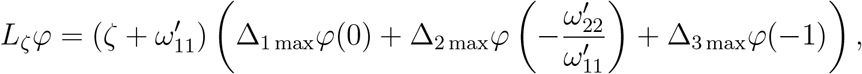

And

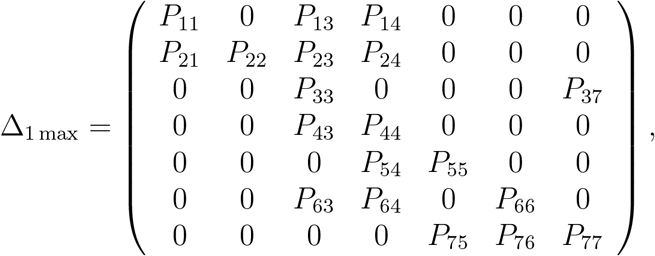

With

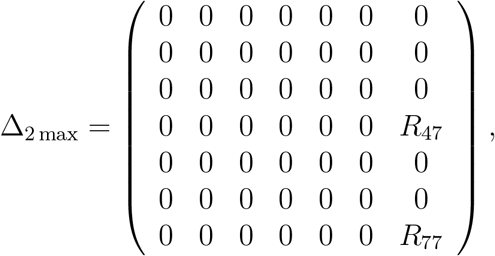

and

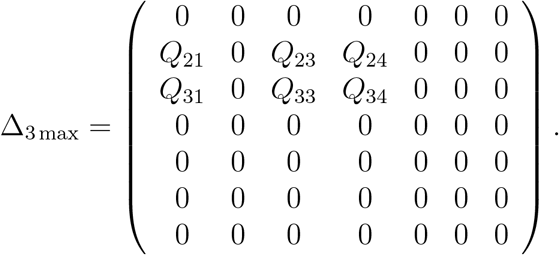

For compactness, define the symmetric bilinear operator B by

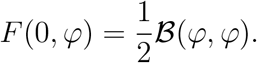

Thus,

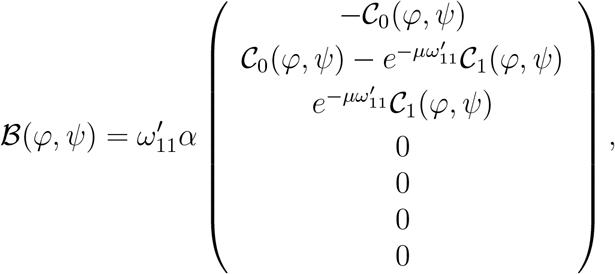

Where

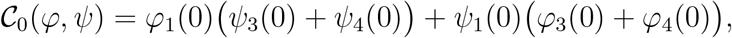

and

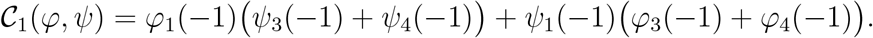

By the Riesz representation theorem [46], there exists a function *ϑ*(*θ, ζ*), whose components are of bounded variation for *θ* ∈ [−1, 0], such that

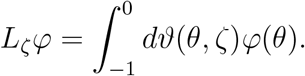

We can choose

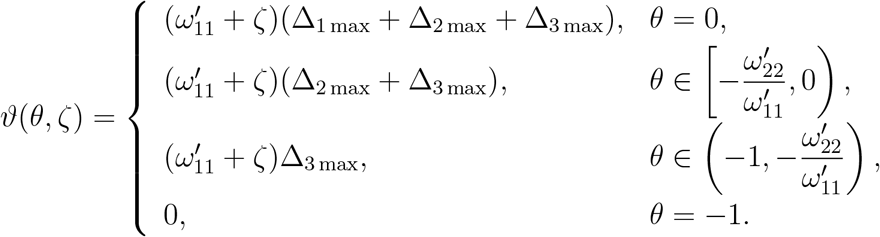

For *φ* ∈ *C*([−1, 0], ℝ^7^), define

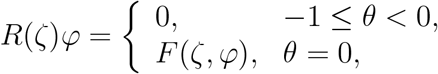

and

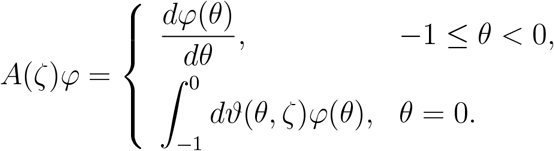

Thus, system (7.23) can be rewritten in the equivalent form

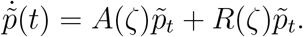

For *κ* ∈ *C*^′^([0, 1], (ℝ^7^)^*^) and *φ* ∈ *C*([−1, 0], ℝ^7^), the bilinear inner form is defined as

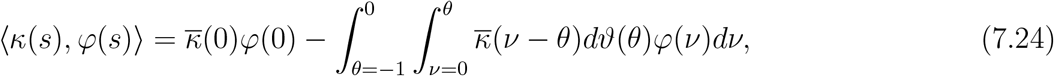

where *ϑ*(*θ*) = *ϑ*(*θ*, 0). For *κ* ∈ *C*^′^([0, 1], (ℝ^7^)^*^), the adjoint operator of *A*(0) is expressed as

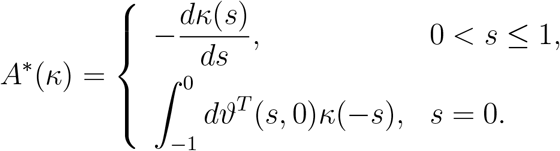

From the previous section, it can be observed that 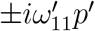 are eigenvalues of *A*(0), and therefore 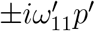 are also eigenvalues of *A* (0). Let

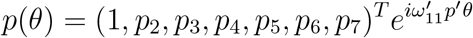

and

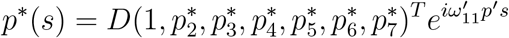

be the eigenvectors of *A*(0) and *A*^*^(0) corresponding to the eigenvalues 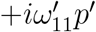 and 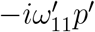, respectively. Hence, we obtain

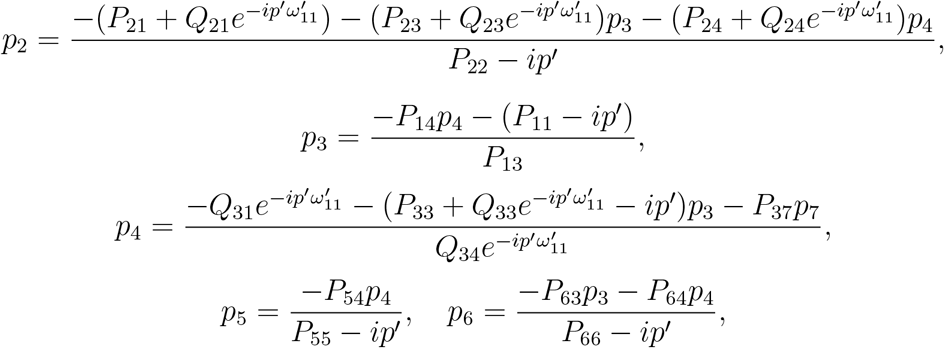

and

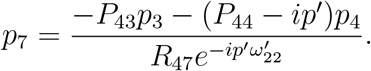

For the adjoint eigenvector, the components satisfy

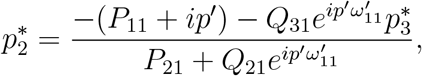

and

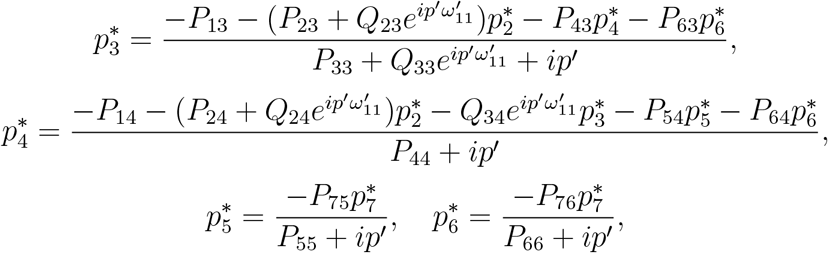

Here, the normalization constant *D* is chosen such that

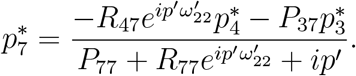

Considering Eq. (7.24), we choose

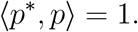

Here, we use similar notation as in Hassard et al. [45] and calculate the coordinates to describe the center manifold *C*_0_ at *ζ* = 0. Let 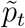be the solution of Eq. (7.23) when *ζ* = 0. Define

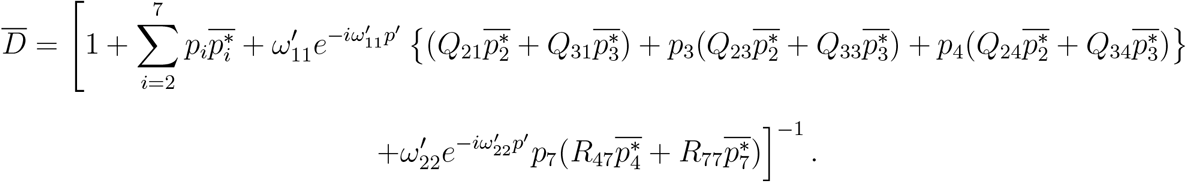

On the center manifold *C*_0_, we have

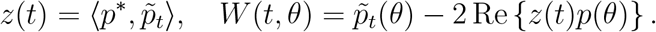

where

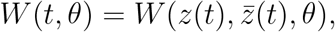

For solutions 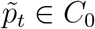, since *ζ* = 0,

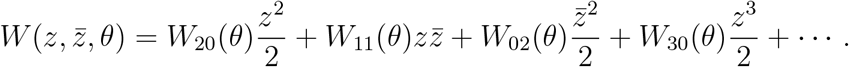

and hence

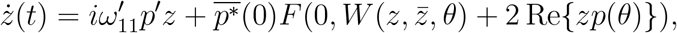

Where

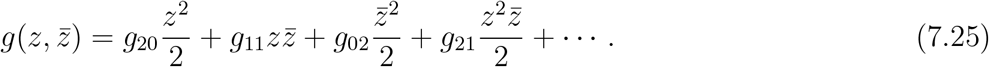

Using the bilinear operator B, the coefficients in (7.25) are obtained as

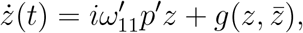

and

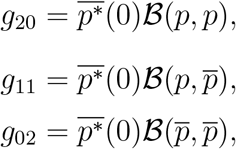

Let

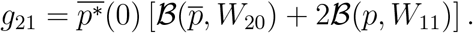

Then

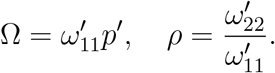

and

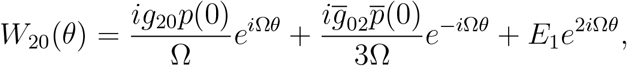

The constant vectors *E*_1_ and *E*_2_ are determined by the following linear systems:

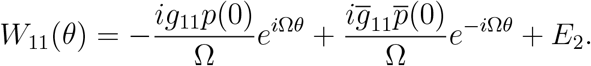

and

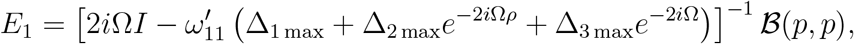

Hence, the first Lyapunov coefficient and the associated normal-form quantities are

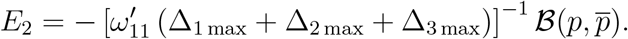

And

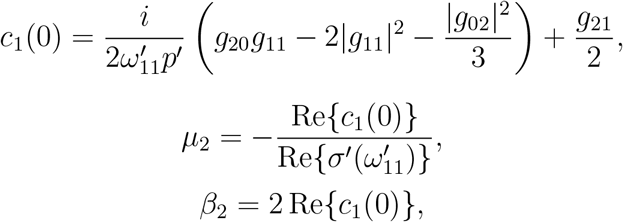

The sign of these coefficients determines the local properties of the Hopf bifurcation. If *µ*_2_ *>* 0, then the Hopf bifurcation is supercritical, whereas if *µ*_2_ *<* 0, then the Hopf bifurcation is subcritical. The bifurcating periodic solutions are orbitally asymptotically stable if *β*_2_ *<* 0, and unstable if *β*_2_ *>* 0. Finally, *T*_2_ *>* 0 indicates that the period of the bifurcating periodic solutions increases with the bifurcation parameter, while *T*_2_ *<* 0 indicates that the period decreases.

**Estimation of the length of time delays to sustain stability**

Here, we present the detailed derivation of the linearized system, Laplace transform formulation, characteristic equation, and inequalities used to estimate the length of time delays to sustain stability by utilizing the Nyquist criterion [47].

Using the ensuing transformations 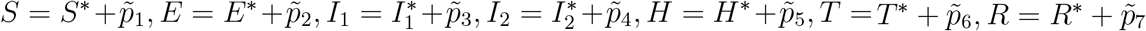, we obtain the linearized system as follows:

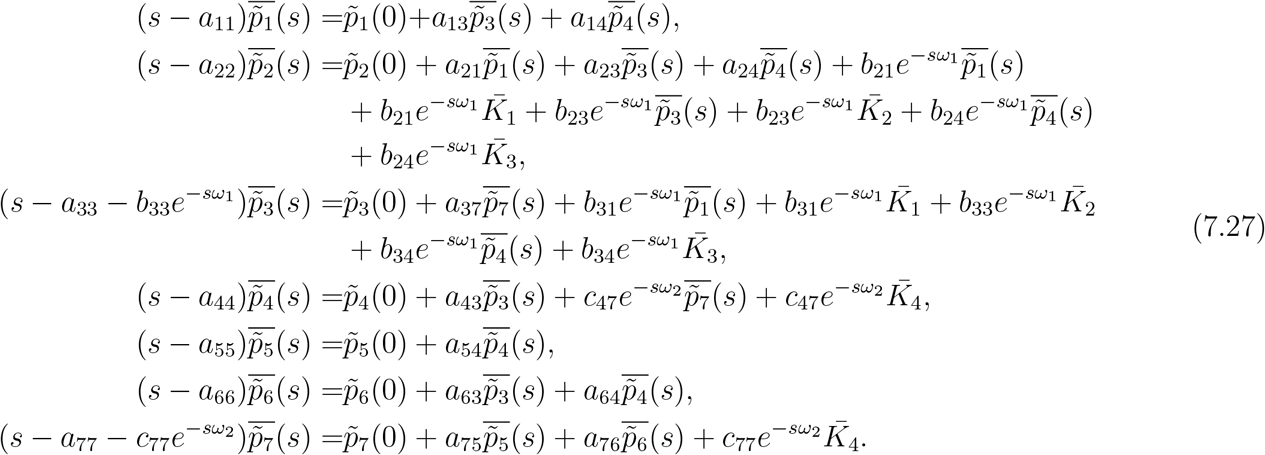

where

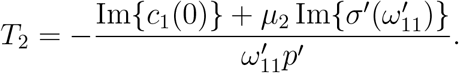

We further obtain the following system by applying Laplace transform on system (7.26):

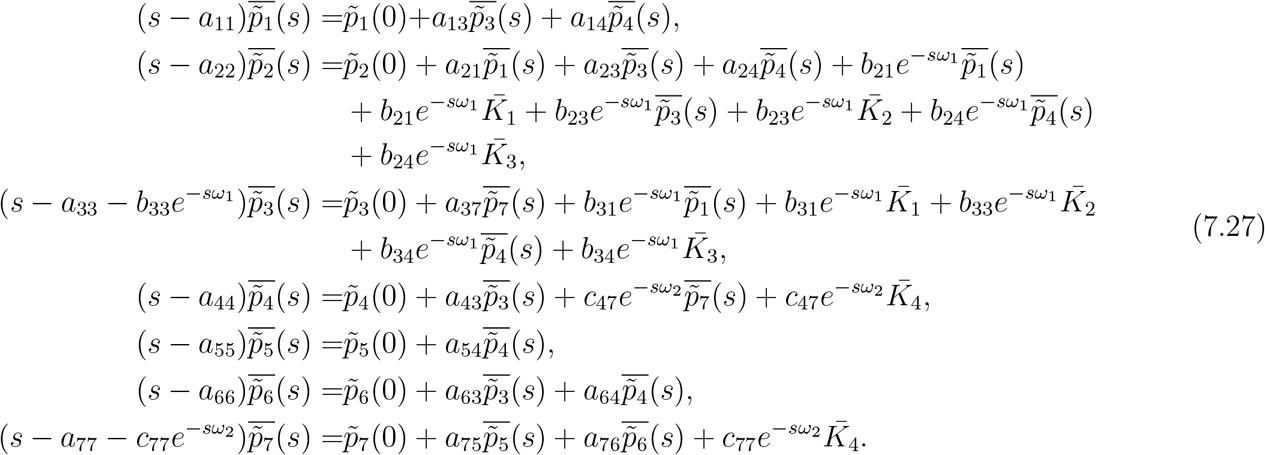

where 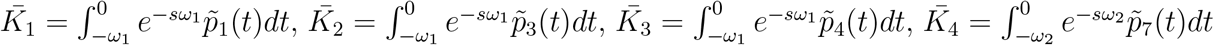, and 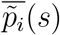 are the Laplace transform of 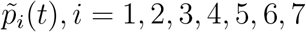 respectively. The characteristic equation of Eq. (7.27) is

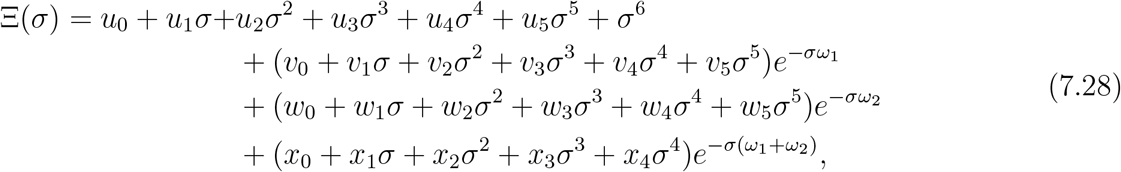

where *u*_*i*_, *v*_*i*_, *w*_*i*_, *i* = 0, 1, 2, 3, 4, 5, and *x*_*i*_, *i* = 0, 1, 2, 3, 4, are provided in the previous section. Local stability of *D*^*^ follows if

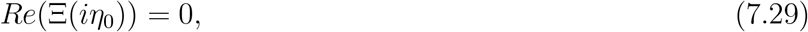

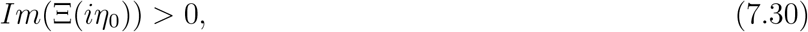

where *η*_0_ is the smallest positive root of Eq. (7.29).

We further ponder the conditions: *ω*_1_ *>* 0 and *ω*_2_ is in its stable interval, as of in Theorem 4.6. Eqs. (7.29) and (7.30) gives

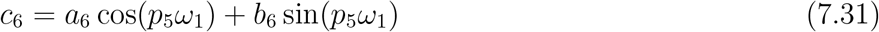

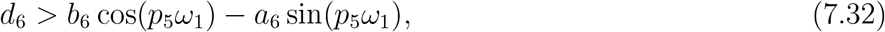

where *a*_6_, *b*_6_, *c*_6_, and *d*_6_ are given in the previous section. From (7.31), we have

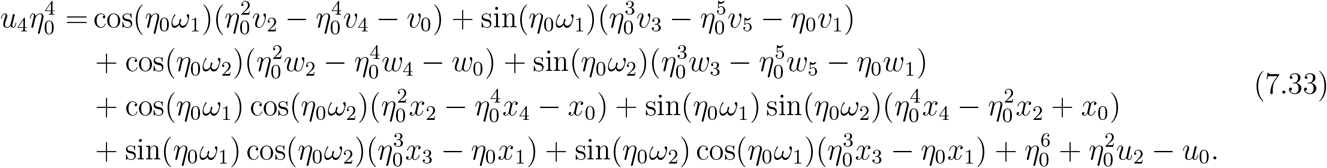

Maximize the right hand side of Eq. (7.33) subject to | sin(*η*_0_*ω*_1_)| ≤ 1, | sin(*η*_0_*ω*_2_)| ≤ 1, | cos(*η*_0_*ω*_1_)| ≤ 1, | cos(*η*_0_*ω*_2_)| ≤ 1, we obtain

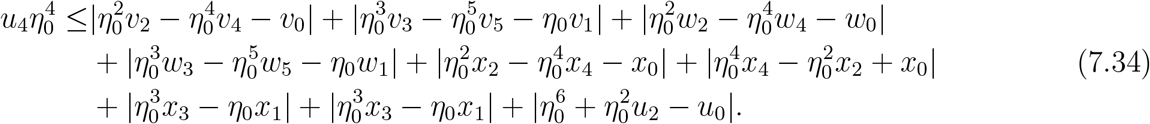

Let Eq. (7.34) has a positive root such that *η*_0_ ≤ *η*^+^. Now, rearranging the inequality (7.32), we obtain

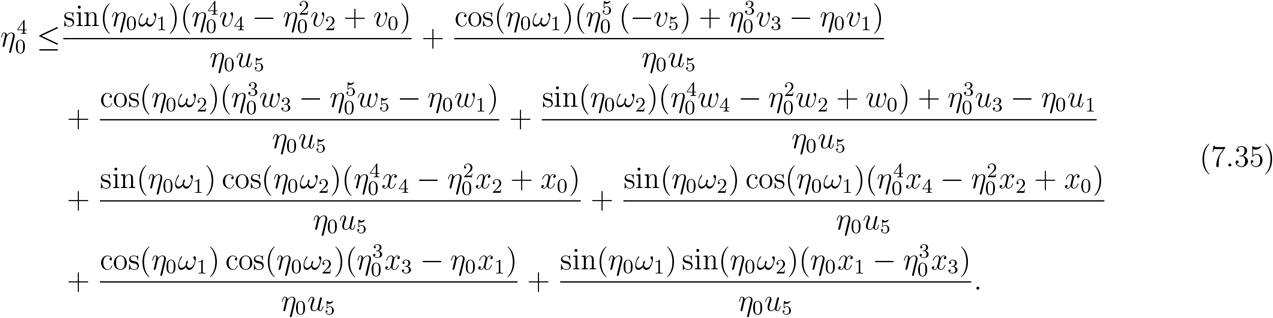

Substituting Eq. (7.33) in Eq. (7.35), assuming |sin(*η*_0_*ω*_2_)| ≤ 1, |cos(*η*_0_*ω*_2_)| ≤ 1, and rearranging, we obtain

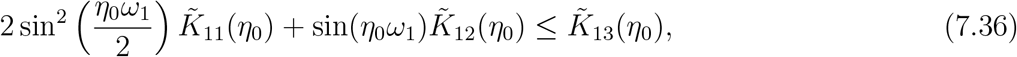

Where

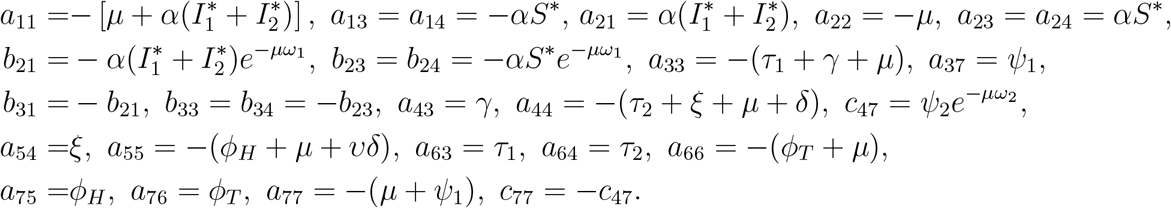

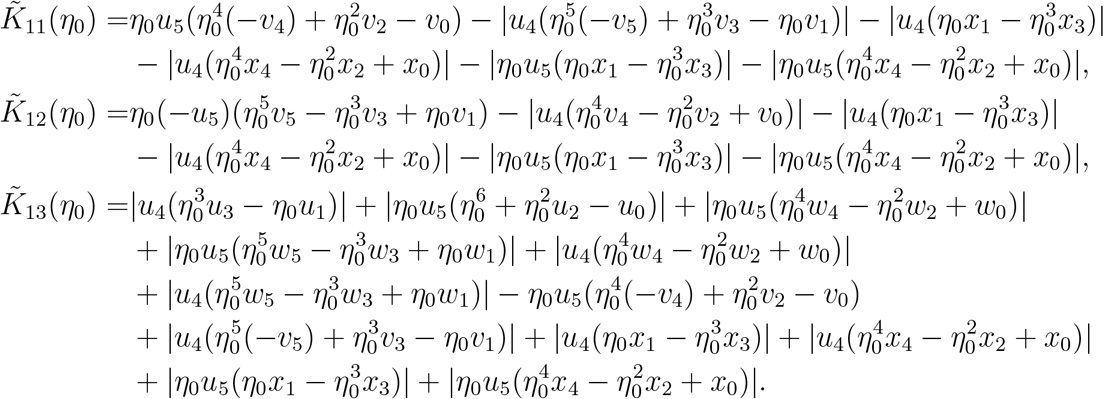

Now, from inequality (7.36), we have

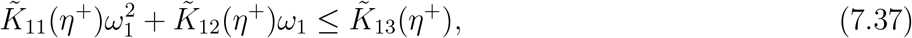

where 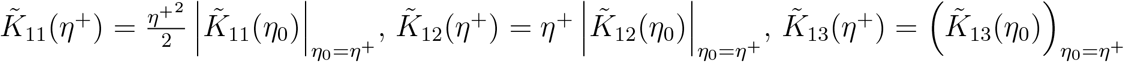. Hence, if

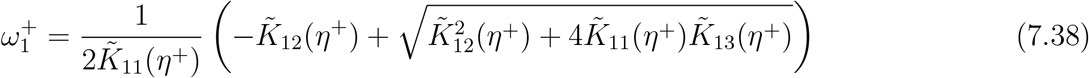

then the stability is preserved for 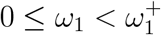.

A similar procedure yields the length of time delay *ω*_2_ where the stability is sustained, considering *ω*_1_ in its stable interval.

## Appendix C

